# Cholera outbreaks in sub-Saharan Africa during 2010-2019: A Descriptive Analysis

**DOI:** 10.1101/2021.10.25.21265347

**Authors:** Qulu Zheng, Francisco J Luquero, Iza Ciglenecki, Joseph F. Wamala, Abdinasir Abubakar, Placide Welo, Mukemil Hussen, Mesfin Wossen, Sebastian Yennan, Alama Keita, Justin Lessler, Andrew S. Azman, Elizabeth C. Lee

## Abstract

**Background:** Cholera remains a public health threat, but is inequitably distributed, especially affecting areas without universal access to safe water and sanitation, including much of sub-Saharan Africa. Lack of standardized reporting and inconsistent outbreak definitions limit our understanding of cholera outbreak epidemiology.

**Methods:** We curated a database of cholera incidence and mortality from sub-Saharan Africa from 2010 to 2020 and developed methods to reconstruct epidemic curves. We then described the distribution of key outbreak metrics, including outbreak size and duration.

**Results:** We identified 999 suspected cholera outbreaks in 744 unique regions across 25 sub-Saharan Africa countries, and outbreak periods accounted for 1.8 billion person-months (2% of the total during this period) from January 2010 through January 2020. Among the 692 outbreaks reported from second-level administrative units (e.g., districts), the median attack rate was 0.8 per 1,000 people (IQR, 0.3-2.4 per 1,000), the median epidemic duration was 13 weeks (IQR, 8-19), and the median early outbreak reproductive number was 1.8 (range, 1.1-3.5). Rural outbreaks had more than twice the case fatality risk than urban ones (median of 1.8% versus 0.8%). Larger attack rates were associated with longer times to outbreak peak, longer epidemic durations, and lower case fatality risks.

**Conclusions:** Despite reporting gaps and the limitations of analyzing outbreaks by administrative units, this work provides a baseline from which to monitor progress towards cholera control and essential statistics to inform outbreak management and emergency response in sub-Saharan Africa.

**Research in context:** *Evidence before this study:* We used PubMed to search for relevant studies published between 2010 and 2019, using the term “Cholera AND (Outbreak OR epidemic) AND (Africa OR Algeria OR Angola OR Benin OR Botswana OR Burkina Faso OR Burundi OR Cabo Verde OR Cameroon OR Central African Republic OR Chad OR Comoros OR Congo Democratic Republic of the OR Congo, Republic of the OR Cote d’Ivoire OR Djibouti OR Egypt OR Equatorial Guinea OR Eritrea OR Eswatini OR Ethiopia OR Gabon OR Gambia OR Ghana OR Guinea OR Guinea-Bissau OR Kenya OR Lesotho OR Liberia OR Libya OR Madagascar OR Malawi OR Mali OR Mauritania OR Mauritius OR Morocco OR Mozambique OR Namibia OR Niger OR Nigeria OR Rwanda OR Sao Tome and Principe OR Senegal OR Seychelles OR Sierra Leone OR Somalia OR South Africa OR South Sudan OR Sudan OR Tanzania OR Togo OR Tunisia OR Uganda OR Zambia OR Zimbabwe)”. Of 544 results, 137 were either not about cholera outbreaks or were about cholera outbreaks in a particular country or context and 407 were not about cholera outbreaks. The remaining three were a review of cholera outbreaks in Africa by Martin et al, who focused on general epidemiology and biology of cholera outbreaks before 2011; a review of epidemiology of cholera by Jacqueline et al, who explored the duration, case fatality rate, genomics, risk factors and surveillance of outbreaks across Africa before 2017; and a study of recurrent outbreaks in Africa by Abraham et al, who examined the historical trends, risk factors, burden, severity and control strategies of outbreaks by geographic regions based on available reports from January, 1970 through August, 2017. In addition, a summary table developed by Médecins Sans Frontières (MSF) for cholera outbreak management and response in the field provided major outbreak characteristics (e.g., attack rate, duration, time to outbreak peak) in rural settings, urban settings and slums, and closed situations (e.g., refugee camps) based on a review of cholera epidemics between 1990 and 1997.

*Added value of this study:* Previous reviews based on available outbreak reports reflect restricted summaries of outbreak characteristics. To present a comprehensive and up-to-date summary for outbreaks in sub-Saharan Africa, we examined key outbreak features by applying a systematic outbreak definition to time series from a large cholera incidence database. To our knowledge, this is the largest centralized source of global cholera incidence and mortality data. We identified 999 suspected cholera outbreaks in 744 sub-national regions across 25 countries in sub-Saharan Africa, where 2% or 1.8 billion person-months of the total population were living in regions with ongoing outbreaks in the period from 2010 through 2019. In addition, our results suggest that population density may not be universally associated with more severe outbreak outcomes. Compared to historical summaries from MSF, our estimates of attack rates and CFRs are much lower in both rural and urban settings, while the estimate of proportion of cases reported during the peak week is slightly higher, leading to only one-third to one-fourth of the previous estimation of peak bed capacity.

*Implications of all the available evidence:* Cholera remains a public health threat in sub-Saharan Africa. This summary of the characteristics and transmission dynamics of outbreaks occurring in sub-Saharan Africa in the period from 2010 through 2019 increases our understanding of cholera outbreak epidemiology and serves as a practical source for future outbreak management and response. As several sub-Saharan African countries have started to developed country plans to reduce cholera incidence in the coming years, our study emphasizes the importance of improving cholera monitoring and surveillance (e.g., laboratory confirmation and finer geographic scale of reporting) in order to identify finer-scale outbreaks, estimate the true burden of cholera and target interventions with limited resources.

## Background

Cholera remains a public health threat worldwide, predominantly in countries with inadequate access to safe water and improved sanitation facilities. Sub-Saharan Africa is one of the regions with the highest cholera burden, where more than 140 million suspected cases are estimated to occur each year in both endemic and epidemic settings.^1^ From 2010 to 2019, 1,080,778 of the 4,426,844 (24%) cholera cases reported to the World Health Organization (WHO) came from sub-Saharan Africa.^2^

Large cholera outbreaks in refugee camps in the late 20th century have shaped public perception about the nature of epidemic cholera in sub-Saharan Africa (e.g., 1994 Goma outbreak among Rwandan refugees),^3^ and data from these outbreaks continue to inform contemporary cholera outbreak responses.^4^ Ministries of health and medical humanitarian organizations, typical actors in emergency cholera outbreak response, use summary statistics on historical cholera outbreaks to set expectations for the duration and magnitude of potential emergency responses.^5–7^ As urban and rural infrastructure has changed over the years and new cholera lineages have proliferated in sub-Saharan Africa, outbreak characteristics may have changed. In addition, while many recent cholera outbreaks are associated with humanitarian crises,^8,9^ outbreaks in refugee and internally displaced people (IDP) camps appear to be less prevalent and less explosive than in times past, likely due to improved camp coordination and preparedness, timely response to outbreaks, and increased preventive cholera vaccination in these settings.^10^

Over the past decade, reported cholera attack rates, case fatality risks (CFR), and estimates of the basic reproductive number have varied widely.^11–16^ This heterogeneity in outbreak characteristics makes it hard to set clear expectations for how outbreaks may unfold. Previous studies of cholera outbreaks have focused on a narrow range of temporal and spatial scales, typically a single outbreak, so it is not often possible to examine factors associated with this heterogeneity.^11–16^ Despite the existence of WHO outbreak definitions, the geographic range and time bounds of cholera outbreaks (e.g., the beginning and end) have been inconsistently defined in the past, further complicating the comparison of such statistics.^17^

Taking advantage of a large, global cholera incidence database, we perform a systematic examination of the characteristics and transmission dynamics of cholera outbreaks occurring in sub-Saharan Africa from January 2010 through January 2020. These results are meant to provide a contemporary picture of cholera outbreaks in the region, while serving as a practical resource in the control and management of cholera outbreaks in the years to come.

## Methods

### Cholera data

We extracted daily and weekly suspected cholera incidence data from the Global Task Force for Cholera Control’s (GTFCC) Global Cholera Database, which contains public and confidential surveillance reports from sources including WHO, MSF, the Program for Monitoring Emerging Diseases (ProMED), ReliefWeb, UNICEF, ministries of health, and the scientific literature.^1,18^ Typical reports were of suspected cholera cases using variants of the WHO-recommended suspected case definition,^19^ though confirmed cases and deaths were also reported in select reports. While this database is not comprehensive, it is, to our knowledge, the largest centralized source of global cholera incidence and mortality data. After aggregating daily incidence data to the weekly level, and averaging reports from different sources that overlapped in space and time, we assembled weekly unified cholera incidence for each sub-national administrative unit (hence, “regions”). Further information on data coverage is described in Tables S1 and S2.

### Population data

Gridded, yearly population data (100m resolution) were obtained from WorldPop for each country that was associated with an outbreak.^20^ Where shapefiles could be found from Humanitarian Data Exchange (HDX), Ministries of Health, GADM, and GAUL,^21–23^ the outbreak region population was estimated using the gridded population data (Figure S1). We defined regions as “urban” when population density was equal to or higher than 1,000 habitants per km^2^, and “rural” when below 1,000 habitants per km^2^ following previous convention.^24,25^

### Outbreak definition

We applied a consistent operational outbreak definition across all regions in our analysis, which assumes that cholera outbreaks are defined by time periods where cholera incidence equals or exceeds the baseline incidence (also called the “outbreak threshold”) in a given region (Figure S2). An outbreak start was defined as a week when weekly cholera incidence reaches the outbreak threshold and is followed by increasing incidence for at least two consecutive weeks. For each unique region, the outbreak threshold was the average weekly cholera incidence during the period between the first and last reported daily or weekly suspected case; for this purpose, weeks without case reports were assumed to have no suspected cases. An outbreak was considered over after two consecutive weeks where weekly cholera incidence remained below the outbreak threshold, provided that this outbreak end was followed by a four-week “wash-out” period where weekly cholera incidence also remained below the outbreak threshold. Reported cases and mortality during the “wash-out” period were not considered as part of the outbreak.

Outbreaks at sub-national administrative unit levels with uninterrupted reporting during the epidemic period were included in our main analysis (Figure S1). We removed outbreaks in the same region with overlapping time periods as duplicates, manually selecting those outbreaks with less censoring and fewer missing fields and observations. Outbreaks reported at different administrative reporting levels with overlapping time periods (e.g., province-level and district-level reporting) were counted as separated outbreaks, and assessed independently in our study.

### Epidemic metrics

We calculated a standard set of metrics for each outbreak. *Attack rate* was defined as suspected cholera cases per 1,000 population living in the outbreak region. *Case-fatality risk* (CFR) was defined as the ratio of cholera-associated deaths to suspected cholera cases in an outbreak. *Time to outbreak peak* was defined as the number of weeks between the start of an outbreak and the week with the greatest number of reported cases. We estimated instantaneous reproductive numbers using the EpiEstim package in R, assuming that cholera’s serial interval follows a Gamma distribution (mean = 4 days, sd=3 days), and that the smoothing window during which transmission was assumed constant was one week.^26^ The *early outbreak reproductive number* was the average of the instantaneous reproductive estimates over the first week of an outbreak (see details in supplement).^26,27^ We used the Wilcoxon rank sum test to test the null hypothesis that urban and rural settings had no differences in epidemic metrics. To further measure the associations between different epidemic metrics, Pearson bivariate correlation tests were performed for outbreaks reported across different administrative units.^28,29^ For those epidemic metrics with a skewed distribution (i.e., outbreak attack rates, outbreak thresholds and CFRs), a log transformation (i.e., log10-transformed scale) was used to normalize the covariate in correlation tests.

### Sensitivity analyses

Although we assumed zero suspected cases for weeks without case reports, many regions only report cases during officially declared outbreaks, leaving the possibility that more sporadic cases were not captured in the surveillance systems.^30–33^ Hence, setting the average of the reported weekly incidence as the fixed outbreak threshold may not reflect the true burden of cholera for a given region. To assess the sensitivity of the threshold and assumption, we repeated the analysis using a different definition for the outbreak threshold, which was defined as the average number of reported suspected cases per week during the first three epidemic weeks with an increasing number of reported cases (see supplements).

Reported cases were aggregated to administrative units (e.g., district, and province levels) in the surveillance systems, while the population of administrative units differed greatly within and between countries.^34^ Therefore, outbreaks reported at the same spatial level may not be comparable to each other in terms of the epidemic characteristics. To test other grouping methods, we further summarized the outbreak statistics by four total population size groups, including regions with the population of <10,000, regions with population between 10,000-100,000, regions with population between 100,000-1,000,000, and regions with population of >1,000,000 (see supplements).

### Code and data availability

All analyses were conducted in the R statistical programming language, version 4.0.2 (R Foundation for Statistical Computing). Outbreak extraction code and aggregated data unlinked from specific geographic locations are available on Github (https://github.com/QLLZ/Cholera-outbreaks).

## Results

From January 2010 through January 2020, we captured 999 cholera outbreaks with a total of 484,450 suspected cholera cases from 744 unique sub-national regions across 25 sub-Saharan African countries in our database. This included 62 outbreaks in 50 unique first-level administrative units, 692 outbreaks in 492 unique second-level administrative units, and 245 outbreaks in 202 unique third-level administrative units (Figures 1 and S1, Table S1). Among them, only 128 sub-national regions reported at least one confirmed cholera case and 876 sub-national regions reported cholera-associated deaths.

**Figure 1.**
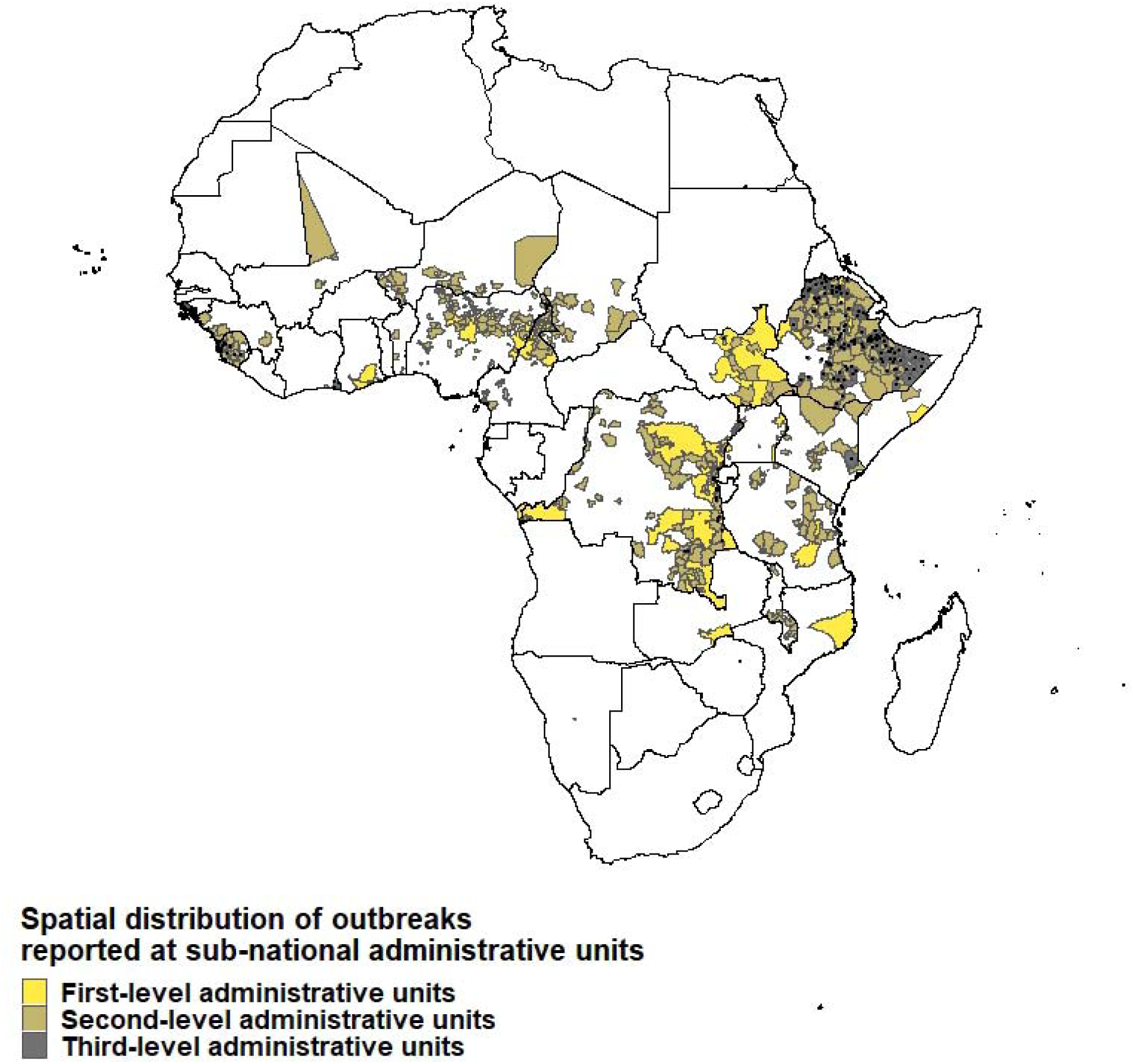
Spatial distribution of outbreaks reported at sub-national administrative units, 2010-2020. This map shows the regions that are associated with reported cholera outbreaks. Different colors represent different sub-national administrative units at which cholera outbreaks were reported. Outbreaks in third-level administrative units are additionally marked with black dots to increase visibility on the map.

Over this 10-year period, 1.8 billion person-months (2% of the total during this period) were spent at risk of a cholera outbreak in sub-Saharan Africa (Figure 2 and Table S2). The collective outbreaks in four countries covered 65% of this total person-month burden -- Democratic Republic of the Congo (27.8%), Ethiopia (27.1%), Cameroon (6.3%), and South Sudan (3.2%). These values corresponded to 5% each of Democratic Republic of the Congo and Ethiopia person-months, and 4% each of Cameroon and South Sudan person-months.

**Figure 2.**
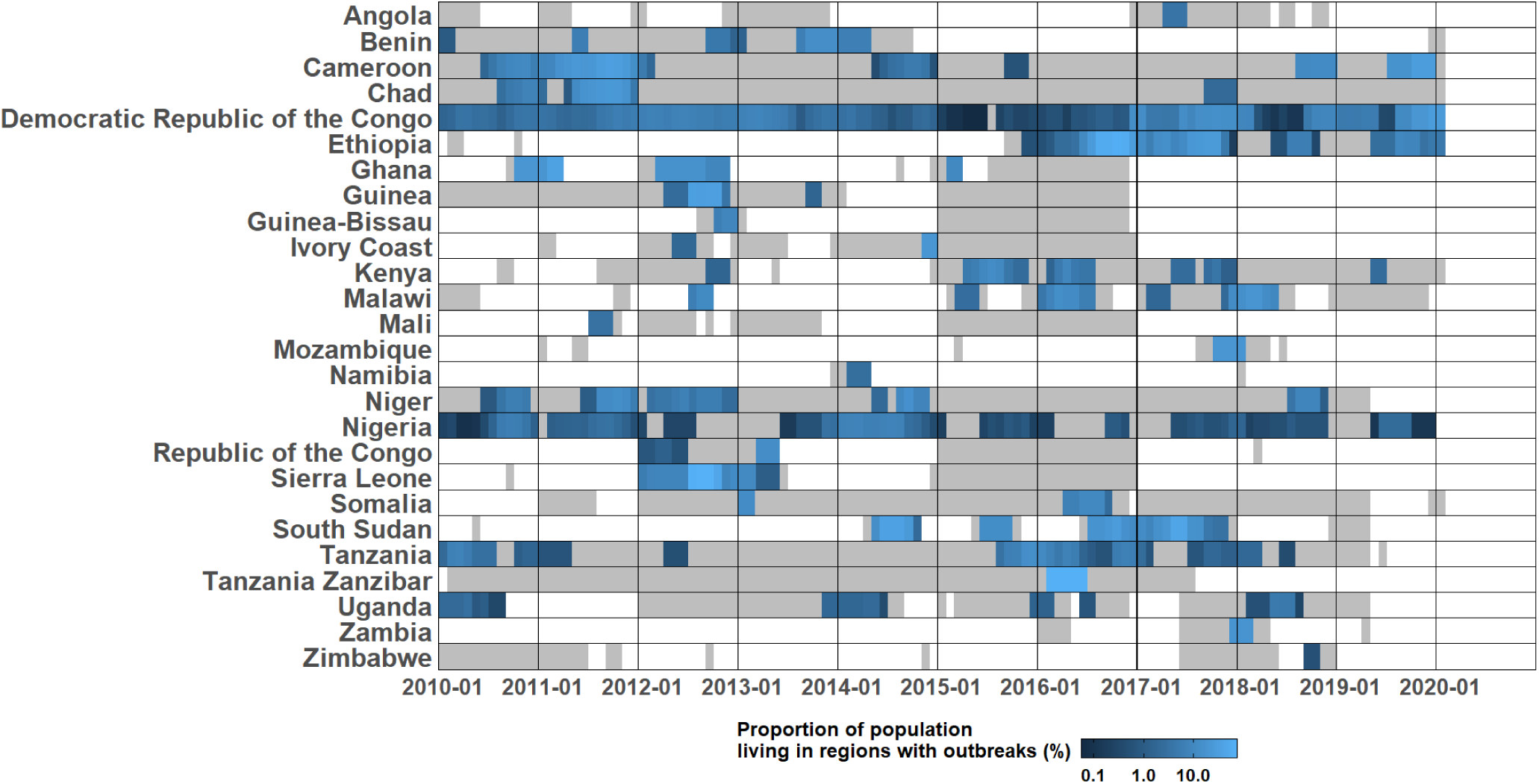
Proportion of population living in regions with outbreaks reported at the sub-national administrative units (%) The proportion of population living in regions with outbreaks reported at sub-national administrative units for each month between 2010-01-01 and 2020-01-31. The areas in grey represent time periods covered by daily and weekly cholera reports. To combine the population at different spatial levels, we added the population of different regions together, and when an outbreak was reported at multiple spatial units, the population of the highest spatial unit was used to represent the population affected by that outbreak.

Outbreaks were extracted and examined by administrative levels, as reported by the original data source (Table 1). Across the 692 suspected case outbreaks reported in second-level administrative units, the median outbreak threshold was 0.7 per 100,000 people per week (interquartile range (IQR), 0.7-2.8 per 100,000 people per week), the median epidemic duration was 13 weeks (IQR, 8-19), the median time to outbreak peak was 4 weeks (IQR, 3-6), the median early outbreak reproductive number was 1.8 (range, 1.1-3.5), and the median attack rate was 0.8 per 1,000 people (IQR, 0.3-2.4 per 1,000 people) (Table 1 and Figures S3-7). One in four cases (median 23.8; IQR 16.4-34.5%) during each outbreak were reported in the week following the epidemic peak, with a median peak week incidence of 0.2 per 1,000 people (IQR, 0.1-0.5 per 1,000 people) (Table 1 and Figures S8-9).

**Table 1.** Summary of cholera outbreaks reported in sub-Saharan Africa, 2010-2020. This table presents the key epidemic metrics of outbreaks by different administrative reporting units, including outbreak size, duration, time to outbreak peak, initial reproductive numbers during the first epidemic week, attack rate, and CFRs.

|  | Outbreaks reported at first-level administrative units | Outbreaks reported at second-level administrative units | Outbreaks reported at third-level administrative units |
| --- | --- | --- | --- |
| Metrics based on suspected cholera cases |  |  |  |
| Number of outbreaks | 62 | 692 | 245 |
| Outbreak threshold, weekly incidence per 100,000 people (IQR) | 0.7<br>(0.7-2.0) | 0.7<br>(0.7-2.8) | 1.7<br>(1.7-12.8) |
| Median outbreak size, cases (IQR) | 620<br>(251-2,191) | 182<br>(70-449) | 100<br>(42-234) |
| Median epidemic durations, weeks (IQR) | 12<br>(8-19) | 13<br>(8-19) | 12<br>(8-16) |
| Median time to epidemic peak, weeks (IQR) | 4<br>(3-5.5) | 4<br>(3-6) | 3<br>(3-5) |
| Median proportion of suspected cases reported during the peak week (%) (IQR) | 19.1<br>(14.8-29.7) | 23.5<br>(16.4-34.2) | 28.8<br>(20.5-38.1) |
| Median weekly incidence during the peak week per 1,000 people (IQR) | 0.1<br>(0.03-0.2) | 0.2<br>(0.1-0.5) | 0.5<br>(0.2-1.5) |
| Median early outbreak reproductive number (range) | 1.8<br>(1.2-2.9) | 1.8<br>(1.1-3.5) | 1.9<br>(1.02-3.2) |
| Median attack rate per 1,000 people (IQR)<sup>b</sup> | 0.5<br>(0.2-1.2) | 0.8<br>(0.3-2.4) | 2.0<br>(0.6-6.4) |
| Metrics based on cholera-associated deaths<sup>a</sup> |  |  |  |
| Number of outbreaks with reports of deaths | 37 | 646 | 193 |
| Median case fatality risk (%)(IQR) | 1.2<br>(0.4-2.1) | 1.6<br>(0.3-4) | 0<br>(0-0.9) |
| Population-weighted case fatality risk (%)<sup>e</sup> | 0.7 | 1.9 | 0.8 |

<sup>a</sup>N.B.: Outbreaks with reports of deaths may not have documented this information systematically, so these results are highly sensitive to reporting biases.
<sup>b</sup>Only outbreaks with valid population estimates are included. There were 62 outbreaks at the first-level administrative units, 657 outbreaks at the second-level units, and 281 outbreaks at the third-level units.
<sup>c</sup>Only outbreaks with reports of deaths and valid population estimates are included. There were 36 outbreaks at the first-level administrative units, 610 outbreaks at the second-level units, and 229 outbreaks at the third-level units.

Outbreaks reported at a higher spatial-scale (e.g., province is higher than village) tended to have more suspected cases (e.g., median suspected cases of 620 versus 100 for first-level and third-level administrative units, respectively) but lower attack rates (e.g., median attack rate of 0.5 per 1,000 versus 2 per 1,000 for first-level and third-level, respectively) (Table 1 and Figures S7 and S10). Epidemic durations (e.g., median of 12-13 weeks across levels), times to peak (e.g., median of 4 weeks across levels), and early outbreak reproductive numbers were similar across spatial reporting units (e.g., median of 1.9-2 across levels) (Table 1 and Figures S4-6).

Only a subset of outbreaks also reported confirmed cases and deaths, and these data were not systematically reported (Table 1, Table S3, and Figures S11-14). For example, only 92 (13%) of second-level administrative unit outbreaks reported any confirmed case data (including zeros) with only 54 (8%) reported at least one confirmed cholera case; a median of 2.2% of suspected cases were confirmed (IQR, 0.5-9.8%) among outbreaks with at least one confirmed case (Table S3 and Figures S11-12). There were 646 (93%) outbreaks at second-level administrative units that reported cholera-associated deaths, with a median CFR of 1.6% (IQR, 0.3-4%) (Table 1 and Figures S13-14).

Outbreaks in rural and urban areas reported at a fine spatial scale (3rd level administrative unit), had different characteristics. Urban outbreaks had higher peak weekly incidence (e.g., median peak weekly incidence of 0.4 versus 0.8 per 1,000 people in rural and urban settings, respectively, p=0.03) (Table 2 and Figure S22), greater attack rates (e.g., median attack rate per 1,000 population of 1.5 versus 2.9 in rural and urban settings, respectively, p=0.002) (Table 2 and Figure S23), longer epidemic durations (e.g., median duration of 12 weeks versus 14 weeks in rural and urban settings, respectively, p=0.03) (Table 2 and Figure S24), and a higher rate of confirmed cases per population (e.g., median confirmed cases per 1,000 population of 0.04 versus 0.7 in rural and urban settings, respectively, p<0.001) (Table S5 and Figure S25). We found different results when comparing outbreaks reported at coarser spatial scales (Table S4). However, it is likely that many of the larger administrative units classified as rural were actually a mix of urban and rural areas, thus challenging the interpretation of these results.

**Table 2.** Outbreaks reported at the third administrative level in rural and urban settings. This table presents the comparisons of epidemic metrics between rural and urban settings for outbreaks at third-level administrative units.

|  | Rural | Urban |
| --- | --- | --- |
| Metrics based on suspected cholera cases |  |  |
| Number of outbreaks | 188 | 57 |
| Outbreak threshold (weekly incidence per 100,000 people) (IQR) | 1.5*<br>(0.5-11.4) | 3.0*<br>(1-28.2) |
| Median outbreak size (IQR) | 100<br>(43-215) | 96<br>(40-469) |
| Median epidemic durations, weeks (IQR) | 12*<br>(8-15) | 14*<br>(10-17) |
| Median time to epidemic peak, weeks (IQR) | 3<br>(3-5) | 4<br>(3-6) |
| Median proportion of suspected cases reported during the peak week (%) (IQR) | 31.4*<br>(22.2-39.2) | 22.7*<br>(16.7-30) |
| Median weekly incidence during the peak week per 1,000 people (IQR) | 0.4*<br>(0.1-1.4) | 0.8*<br>(0.3-3) |
| Median early outbreak reproductive number (range) | 1.9<br>(1.02-3) | 2.0<br>(1.3-3.2) |
| Median attack rate per 1,000 people (IQR) | 1.5*<br>(0.6-5.7) | 2.9*<br>(1.9-10.9) |
| Metrics based on cholera-associated deaths<sup>a</sup> |  |  |
| Number of outbreaks with reports of deaths | 138 | 55 |
| Median case fatality risk (%) (IQR) | 0<br>(0-1) | 0<br>(0-0.2) |

\* P<0.05 Wilcoxon rank sum test was used to test if the medians of outbreak characteristics are the same between rural and urban settings.
<sup>a</sup>N.B.: Outbreaks with reports of deaths may not have documented this information systematically, so these results are highly sensitive to reporting biases.

We explored the relationships between outbreak metrics, focusing on outbreaks at the second administrative level (Figure 3, see Figures S26-27 for bivariate relationships at different spatial scales). Two comparisons, outbreak threshold versus attack rate (correlation = 0.75, p<0.0001), and time to outbreak peak versus epidemic duration had strong positive relationships (correlation= 0.62, p<0.0001). Larger attack rates were also associated with longer times to outbreak peak (correlation=0.2, p<0.0001) and longer epidemics (correlation=0.4, p<0.0001). Larger attack rates also appeared to have a negative relationship with CFRs (correlation=-0.3, p<0.0001). The early reproductive number did not appear to have an association with other outbreak metrics. These relationships were consistent across different spatial reporting units.

**Figure 3.**
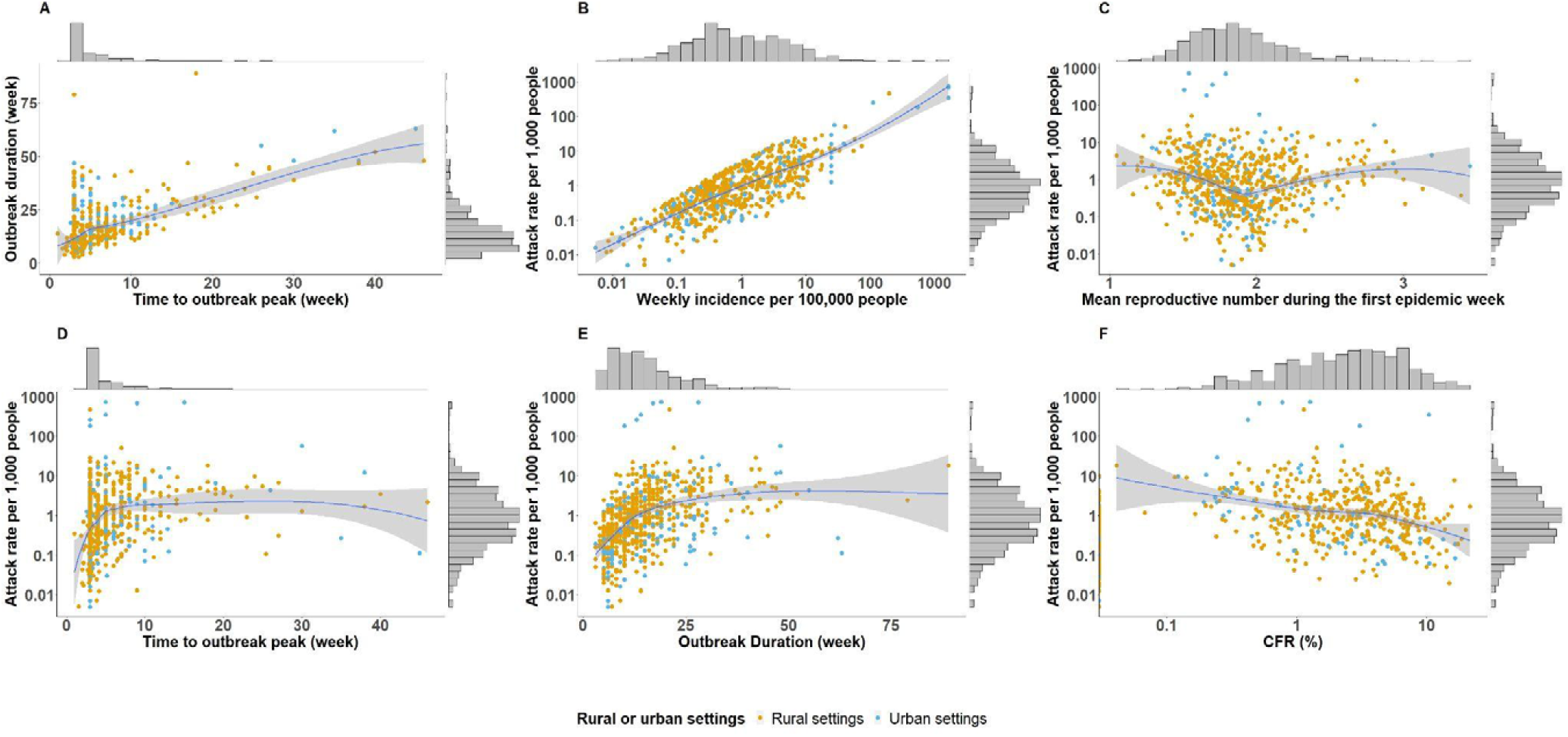
Bivariate relationships between epidemic metrics among outbreaks reported at the second-level administrative units. This figure shows the correlations between different epidemic metrics for second-level administrative unit outbreaks, including outbreak threshold, mean reproductive number during the first epidemic week, attack rate, duration, time to outbreak peak and CFR. The marginal histograms show the distributions of univariates. Panel A shows the correlation between time to outbreak peak (week) and outbreak duration (week). Panel B shows the correlation between outbreak threshold (i.e., weekly incidence per 100,000 people) and attack rate per 1,000 people. Panel C shows the correlation between mean reproductive number during the first epidemic week and attack rate per 1,000 people. Panel D shows the correlation between time to outbreak peak (week) and attack rate per 1,000 people. Panel E shows the correlation between outbreak duration (week) and attack rate per 1,000 people. Panel F shows the correlation between CFR (%) and attack rate per 1,000 people.

As a sensitivity analysis, we extracted outbreaks using an alternate outbreak threshold definition (see Methods) and found only minor changes to summary outbreak metrics (see supplement and Tables S6-7 and Figures S39-74). We also examined outbreak metrics after grouping outbreaks by population size instead of administrative units (Table S8 and Figures S28-38). Case fatality risk increased monotonically for the three smaller population sized groups and declined again for outbreaks in regions with populations greater than 1 million (Table S8 and Figure S28). Early reproductive numbers, outbreak duration and time to outbreak peak were similar across regions with different population sizes (Table S8 and Figures S29-31).

## Discussion

We shed new light on cholera outbreaks in Sub-Saharan Africa over the past decade by applying a systematic outbreak definition to time series from a large database of cholera incidence. We found that 2% or 1.8 billion person-months of the total sub-Saharan Africa population were at risk in regions with ongoing cholera outbreaks in the period from 2010 through 2019; this impact was spread across 999 suspected cholera outbreaks in 744 sub-national regions across 25 sub-Saharan African countries.

Timing (time to outbreak peak and epidemic duration) and incidence rate metrics (outbreak threshold and attack rate) had strong positive correlations with each other, and as may be expected, outbreaks with larger attack rates tended to be longer. Outbreaks with larger attack rates tended to have lower CFRs; the low specificity of suspected cholera case definitions and limited laboratory testing for cholera, may mean that attack rates are overestimated, CFRs are underestimated, and that these metrics are negatively correlated.^8^

While a few studies report greater relative cholera burden and mortality in rural areas, high population density and urban settings have long been thought to be drivers of *V. cholerae* in outbreaks.^35–39^ Our results add nuance to this general wisdom, in that population density may not be universally associated with more severe cholera outbreak outcomes; attack rates were higher and epidemics were longer in urban settings for third-level administrative unit outbreaks, but the peak outbreak week captured a larger percentage of total outbreak cases in rural settings.

Summaries of historical outbreaks have served as the basis of planning for emergency cholera responses for decades, enabling organizations like Médecins Sans Frontières (MSF) and UNICEF to perform data-driven allocation of resources like beds and rehydration fluids.^40^ Compared to existing planning resource tables, our results estimate that contemporary outbreaks in sub-Saharan Africa have much lower attack rates and CFRs in both urban and rural settings, a slightly higher proportion of cases reported during the peak week, and that peak bed capacity needs may be only one-third to one-fifth of existing estimates.^41^ Unlike the outbreak-specific surveillance used to develop these planning resource tables, our analyses used heterogeneous surveillance sources aggregated to administrative units, so these metrics may not be directly comparable.

This work may also support decision making related to reactive oral cholera vaccine (OCV) campaigns, as our descriptive analysis may be used to inform prospective vaccine impact estimates for a given campaign. In addition, future policy guidance for emergency OCV requests may wish to leverage the positive association between outbreak incidence thresholds and overall outbreak attack rates (Figure 3 and Figures S26-27) to triage ongoing outbreak locations at greatest risk for a high attack rate.

Our analysis suggests that cholera outbreaks at larger geographic scales may be a compilation of outbreaks at smaller geographic scales, as duration, peak timing, and early reproductive number metrics remained relatively stable across geographic scales (and for an alternate outbreak threshold definition, Table S4). The cholera incidence and mortality data were reported at the scale of administrative units (as opposed to “outbreak” scale), which means that a single outbreak spanning multiple administrative units appears as multiple outbreaks, and outbreaks below the level of an administrative reporting unit would be aggregated to a higher scale. Our metrics are likely sensitive to reporting units, which vary greatly in size by country, and may not directly translate to outbreak metrics reported elsewhere, particularly for regions with smaller populations (see Table S8 for summary statistics by population size). However, third-level administrative units observed the highest outbreak thresholds, attack rates, and proportions of cases reported in the peak week (compared to larger geographic scales), and an assessment of outbreaks overlapping in space and time found that it was common for multiple third-level administrative units to report concurrent outbreaks at the same time as their surrounding second-level administrative unit (Figures S75-77). This suggests that multiple, geographically-proximate administrative level 3 units may often represent an epidemiologically-relevant geographic region for cholera outbreaks.

This work represents one of the most comprehensive analyses of cholera outbreaks, but, reporting gaps between outbreaks posed challenges to outbreak extraction and may have biased our outbreak metrics; an average of 54% (range 5-100%) of the study period had some surveillance data coverage across all countries. Cholera surveillance data is not collected systematically across sub-Saharan Africa and fewer than 13% of our reported outbreaks had even one confirmed case, making it hard to understand the true cholera burden and progress in control. Our analysis leverages but is also limited by its disparate data sources, case definitions, and testing and reporting protocols. Further, our outbreak threshold definition does not account for cholera seasonality (which has not been systematically characterized), although sensitivity analysis suggests that our results are robust to alternate non-seasonal baseline definitions (Table S7-8).

This study serves as an important baseline from which to monitor progress towards cholera control in sub-Saharan Africa and provides essential information to inform cholera outbreak management and emergency response. As several countries have started to announce ambitious plans for large-scale reductions in cholera incidence and outbreak frequency, improvements in surveillance and reporting, including laboratory confirmation, will be key not only to describing the true burden but also targeting interventions with scarce resources.

## Data Availability

All data produced in the present study are available upon reasonable request to the authors.

## Acknowledgments

We thank all of the ministries of health, WHO offices, and the UNICEF WCAR and ESAR cholera platforms for contributing data to the database. We thank Sean Moore, Heather McKay, Joshua Kaminsky, Maya Demby, Rachel DePencier, Hridika Shah, Van To, Lauren Norris, Ellen Ferriss, Isadora Salles, Keya Joshi, Hojoon Lee, Ananya Kumar, Qifang Bi, and Forrest Jones for assistance with data management and data entry over the past years. We thank Dominique Legros for initial discussions that led to this manuscript and David Olson, Kathryn Alberti, and Morgane Dominguez for their feedback on an early draft of the manuscript.

## Funding

QZ, JL, ASA, and ECL were supported by the Bill and Melinda Gates Foundation (INV-002667).

## Declaration of interests

We have no competing interests to declare.

## Notes

### Competing Interest Statement

The authors have declared no competing interest.

### Author Declarations

The IRB of Johns Hopkins Bloomberg School of Public Health deemed this work as non-human subjects research.

## Reference

1. Lessler J, Moore SM, Luquero FJ, McKay HS, Grais R, Henkens M, et al. Mapping the burden of cholera in sub-Saharan Africa and implications for control: an analysis of data across geographical scales. Lancet. 2018 May 12;391(10133):1908–15.

2. WHO | WER 2020 Index. 2020 Dec 18 [cited 2021 Apr 12]; Available from: http://www.who.int/wer/2020/en/

3. Public health impact of Rwandan refugee crisis: what happened in Goma, Zaire, in July, 1994ã Goma Epidemiology Group. Lancet. 1995 Feb 11;345(8946):339–44.

4. Cholera Outbreak Response field manual [Internet]. [cited 2021 Sep 25]. Available from: https://choleraoutbreak.org/home

5. Management of A CHOLERA EPIDEMIC - Management of a CHOLERA EPIDEMIC [Internet]. [cited 2021 Apr 14]. Available from: https://medicalguidelines.msf.org/viewport/CHOL/english/management-of-a-cholera-epidemic-23444438.html

6. Khambira M. UNICEF Cholera Toolkit [Internet]. [cited 2021 Apr 14]. Available from: https://plateformecholera.info/index.php/resources/tools/technical-books/692-unicef-cholera-toolkit

7. Cholera Outbreak Response field manual [Internet]. [cited 2021 Sep 25]. Available from: https://choleraoutbreak.org/home

8. Camacho A, Bouhenia M, Alyusfi R, Alkohlani A, Naji MAM, de Radiguès X, et al. Cholera epidemic in Yemen, 2016-18: an analysis of surveillance data. Lancet Glob Health. 2018 Jun;6(6):e680–90.

9. Jones FK, Wamala JF, Rumunu J, Mawien PN, Kol MT, Wohl S, et al. Successive epidemic waves of cholera in South Sudan between 2014 and 2017: a descriptive epidemiological study. Lancet Planet Health. 2020 Dec;4(12):e577–87.

10. Shannon K, Hast M, Azman AS, Legros D, McKay H, Lessler J. Cholera prevention and control in refugee settings: Successes and continued challenges. PLoS Negl Trop Dis. 2019 Jun;13(6):e0007347.

11. Msyamboza KP, Kagoli M, M’bang’ombe M, Chipeta S, Masuku HD. Cholera outbreaks in Malawi in 1998-2012: social and cultural challenges in prevention and control. J Infect Dev Ctries. 2014 Jun 11;8(6):720–6.

12. Troeger C, Gaudart J, Truillet R, Sallah K, Chao DL, Piarroux R. Cholera Outbreak in Grande Comore: 1998-1999. Am J Trop Med Hyg. 2016 Jan;94(1):76–81.

13. Denue BA, Akawu CB, Kwayabura SA, Kida I. Low case fatality during 2017 cholera outbreak in Borno State, North Eastern Nigeria. Ann Afr Med. 2018 Oct;17(4):203–9.

14. Gbary AR, Dossou JP, Sossou RA, Mongbo V, Massougbodji A. [Epidemiologic and medico-clinical aspects of the cholera outbreak in the Littoral department of Benin in 2008]. Med Trop. 2011 Apr;71(2):157–61.

15. Mukandavire Z, Liao S, Wang J, Gaff H, Smith DL, Morris JG Jr. Estimating the reproductive numbers for the 2008-2009 cholera outbreaks in Zimbabwe. Proc Natl Acad Sci U S A. 2011 May 24;108(21):8767–72.

16. Eisenberg MC, Robertson SL, Tien JH. Identifiability and estimation of multiple transmission pathways in cholera and waterborne disease. J Theor Biol. 2013 May 7;324:84–102.

17. Cholera [Internet]. [cited 2021 Apr 14]. Available from: https://www.who.int/news-room/fact-sheets/detail/cholera

18. Moore SM, Azman AS, Zaitchik BF, Mintz ED, Brunkard J, Legros D, et al. El Niño and the shifting geography of cholera in Africa. Proc Natl Acad Sci U S A. 2017 Apr 25;114(17):4436–41.

19. Cholera [Internet]. [cited 2021 Jun 10]. Available from: https://www.who.int/news-room/fact-sheets/detail/cholera

20. Worldpop. WorldPop [Internet]. [cited 2021 Apr 13]. Available from: https://www.worldpop.org/

21. Welcome - Humanitarian Data Exchange [Internet]. [cited 2021 Apr 13]. Available from: https://data.humdata.org/

22. GADM [Internet]. [cited 2021 Apr 13]. Available from: https://gadm.org/

23. GeoNetwork Team. GeoNetwork opensource portal to spatial data and information. 2007 Feb 7 [cited 2021 Apr 13]; Available from: http://www.fao.org/geonetwork/srv/en/main.home

24. Chen Y-C, Yu S-H, Chen W-J, Huang L-C, Chen C-Y, Shih H-M. Dispatcher-Assisted Cardiopulmonary Resuscitation: Disparity between Urban and Rural Areas [Internet]. Vol. 2020, Emergency Medicine International. 2020. p. 1–7. Available from: http://dx.doi.org/10.1155/2020/9060472

25. Hay SI, Guerra CA, Tatem AJ, Atkinson PM, Snow RW. Urbanization, malaria transmission and disease burden in Africa. Nat Rev Microbiol. 2005 Jan;3(1):81–90.

26. Bi Q, Abdalla FM, Masauni S, Reyburn R, Msambazi M, Deglise C, et al. The Epidemiology of Cholera in Zanzibar: Implications for the Zanzibar Comprehensive Cholera Elimination Plan. J Infect Dis. 2018 Oct 15;218(suppl_3):S173–80.

27. Cori A, Ferguson NM, Fraser C, Cauchemez S. A new framework and software to estimate time-varying reproduction numbers during epidemics. Am J Epidemiol. 2013 Nov 1;178(9):1505–12.

28. Best DJ, Roberts DE. Algorithm AS 89: The Upper Tail Probabilities of Spearman’s Rho [Internet]. Vol. 24, Applied Statistics. 1975. p. 377. Available from: http://dx.doi.org/10.2307/2347111

29. Hollander M, Wolfe DA, Chicken E. Nonparametric Statistical Methods. John Wiley & Sons; 2013. 848 p.

30. Ganesan D, Gupta SS, Legros D. Cholera surveillance and estimation of burden of cholera. Vaccine. 2020 Feb 29;38 Suppl 1:A13–7.

31. Deen J, Mengel MA, Clemens JD. Epidemiology of cholera. Vaccine. 2020 Feb 29;38 Suppl 1:A31–40.

32. Balakrish Nair G, Takeda Y. Cholera Outbreaks. Springer; 2014. 263 p.

33. Ajayi A, Smith SI. Recurrent cholera epidemics in Africa: which way forwardã A literature review [Internet]. Vol. 47, Infection. 2019. p. 341–9. Available from: http://dx.doi.org/10.1007/s15010-018-1186-5

34. Fontanelli O, Miramontes P, Cocho G, Li W. Population patterns in World’s administrative units [Internet]. Vol. 4, Royal Society Open Science. 2017. p. 170281. Available from: http://dx.doi.org/10.1098/rsos.170281

35. An Ecological Study of the Cholera Outbreak in Rural and Urban Areas of Haiti. [Internet]. [cited 2021 Apr 12]. Available from: https://scholarworks.gsu.edu/cgi/viewcontent.cgi?article=1009&context=iph_capstone

36. Page A-L, Ciglenecki I, Jasmin ER, Desvignes L, Grandesso F, Polonsky J, et al. Geographic distribution and mortality risk factors during the cholera outbreak in a rural region of Haiti, 2010-2011. PLoS Negl Trop Dis. 2015 Mar;9(3):e0003605.

37. Mengel MA, Delrieu I, Heyerdahl L, Gessner BD. Cholera outbreaks in Africa. Curr Top Microbiol Immunol. 2014;379:117–44.

38. Cowman G, Otipo S, Njeru I, Achia T, Thirumurthy H, Bartram J, et al. Factors associated with cholera in Kenya, 2008-2013. Pan Afr Med J. 2017 Oct 3;28:101.

39. Penrose K, de Castro MC, Werema J, Ryan ET. Informal urban settlements and cholera risk in Dar es Salaam, Tanzania. PLoS Negl Trop Dis. 2010 Mar 16;4(3):e631.

40. Management of A CHOLERA EPIDEMIC - Management of a CHOLERA EPIDEMIC [Internet]. [cited 2021 Aug 31]. Available from: https://medicalguidelines.msf.org/viewport/CHOL/english/management-of-a-cholera-epidemic-23444438.html

41. 2.7 Estimation of treatment resource needs - Management of a CHOLERA EPIDEMIC [Internet]. [cited 2021 Sep 9]. Available from: https://medicalguidelines.msf.org/viewport/CHOL/english/2-7-estimation-of-treatment-resource-needs-23448748.html

42. Azman AS, Parker LA, Rumunu J, Tadesse F, Grandesso F, Deng LL, et al. Effectiveness of one dose of oral cholera vaccine in response to an outbreak: a case-cohort study [Internet]. Vol. 4, The Lancet Global Health. 2016. p. e856–63. Available from: http://dx.doi.org/10.1016/s2214-109x(16)30211-x

43. Ferreras E, Chizema-Kawesha E, Blake A, Chewe O, Mwaba J, Zulu G, et al. Single-Dose Cholera Vaccine in Response to an Outbreak in Zambia [Internet]. Vol. 378, New England Journal of Medicine. 2018. p. 577–9. Available from: http://dx.doi.org/10.1056/nejmc1711583

44. M’bangombe M, Pezzoli L, Reeder B, Kabuluzi S, Msyamboza K, Masuku H, et al. Oral cholera vaccine in cholera prevention and control, Malawi. Bull World Health Organ. 2018 Jun 1;96(6):428–35.

45. Parker LA, Rumunu J, Jamet C, Kenyi Y, Lino RL, Wamala JF, et al. Adapting to the global shortage of cholera vaccines: targeted single dose cholera vaccine in response to an outbreak in South Sudan. Lancet Infect Dis. 2017 Apr;17(4):e123–7.

