## Supplementary Appendix for "Cholera outbreaks in sub-Saharan Africa during 2010-2019: A Descriptive Analysis"

#### Cholera data

Daily and weekly cholera incidence data through January 2010 to January 2020 were obtained from the Global Taskforce for Cholera Control's (GTFCC) Global Cholera Database. All data were in a standard format that documented the spatial and temporal information for reported cholera cases and cholera-associated deaths. Nationwide and region-wide linelist data were aggregated into daily counts at the reporting administrative unit, and each higher-level administrative unit, assuming that they included all the suspected cases and deaths throughout the region over the reporting period. All daily cholera incidence data were aggregated to the weekly level and merged into weekly time series. Weekly data from the same region and reporting period were averaged. To get non-overlapping time series, we grouped weekly data into different time series based on the first start day of the reporting week for each observation. Outbreak extractions were conducted separately in these time series.

#### Location audit

To clean the spatial data in our dataset, we used the official government documentations, cholera situation reports, google maps, etc, to verify the location names and boundaries over time. With the help of open shapefiles sources and ministries of health, including GAUL, GADM, OCHA, etc, we obtained validated shapefiles over time for each region. [1–3] For regions at the fourth-level administrative units or lower, we did not find validated shapefiles for them. Thus, they were excluded from our main analysis.

For outbreaks reported in Moba health district, Katuba health district, Kenya health district, Mufunga Sampwe health district, and Baraka at fizi health district in the Democratic Republic of Congo, and outbreaks reported in Degehabur City of Jarar and Kebridehar City of Korahe in Somali region of Ethiopia, they had outbreak attack rates exceeding 120 per 1,000 people possibly due to the underestimated yearly population. We used the population data from the Ministry of Health and MSF as the yearly population estimates for these cases.

#### Epidemic Metrics

Instantaneous reproductive numbers ($R_t$), the average number of secondary cases infected by an infectious case at time t, were estimated using a previously described Bayesian framework. [4] We assumed the serial interval of cholera followed a shifted Gamma distribution with a mean of 4, a standard deviation of 3, and a one-week smoothing window to stabilize the instantaneous estimates, during which the transmission was assumed constant. [4–6] We also assumed that $R_t$ followed a Gamma distribution with a mean of 2 and a standard deviation of 0.7, based on previous studies. [4–6] $R_t$ was estimated based on daily cholera incidence data. For cases where only weekly data were available, we disaggregated the weekly data into the daily level and redistributed cases to each day assuming that cholera cases increased at a constant rate throughout the time interval. [4] The daily cases were predicted using cumsum and cm.spline functions. [7-8] We took the mean of $R_t$ during the first epidemic week to represent the initial reproductive number during the early stage of an outbreak.

**Sensitivity analysis**

We performed a sensitivity analysis with a different outbreak threshold definition (see supplement, and Tables S6-7 for details), and identified 113 outbreaks reported at 79 unique first-level administrative units, 1,087 outbreaks reported at 663 unique second-level administrative units, 399 outbreaks reported at 283 unique third-level administrative units and 8 outbreaks reported at 6 unique fourth-level administrative units or lower (Table S6). For outbreaks across different spatial reporting units, the distributions of outbreak metrics remained similar across different outbreak threshold definitions (Figures S34-63), while outbreaks identified in the sensitivity analyses had higher median early reproductive number during the first epidemic week (e.g., median of 1.9 in sensitivity analyses compared to 1.8 in main analyses for outbreaks reported at the second-level administrative units) and lower proportion of confirmed cases (e.g., median of 0 versus 0.4% in sensitivity and main analyses for outbreaks reported at the second-level administrative units), compared to outbreaks in our main analyses (Table 1, Table S6 and Figures S34-35, S44-45 and S54-55).

**Table S1. Cholera outbreaks with complete data by country and spatial reporting units**

This table presents the key epidemic metrics by country and spatial reporting units, including number of outbreaks, outbreak size, outbreak duration, time to outbreak peak, attack rate per 1,000 people, confirmed cases, and cholera-associated deaths.

| Country | Spatial reporting units | Number.of.outbreaks | Median outbreak size (range) | Median epidemic durations weeks (range) | Median time to outbreak peak (range) | Median attack rate per 1,000 people (range) | Number of outbreaks with confirmed cases | Median number of confirmed cases (range) | Percent of cases that were confirmed (range) | Median confirmed cases per 1,000 people per outbreak (range) | Number of outbreaks with deaths | Median CFR (range) | Population-weighted. CFR (%) |
|---|---|---|---|---|---|---|---|---|---|---|---|---|---|
| Angola | First administrative level | 1 | 61 (61-61) | 7 (7-7) | 5 (5-5) | 0.1 (0.1-0.1) | 0 | | | | 0 | | 0 |
| Benin | Second administrative level | 7 | 62 (16-312) | 10 (6-32) | 3 (3-5) | 0.8 (0-1.5) | 0 | | | | 5 | 0.5 (0-3.4) | 0.2 |
| Cameroon | First administrative level | 3 | 328 (136-448) | 15 (5-22) | 8 (3-9) | 0.1 (0-0.2) | 0 | | | | 1 | 3.4 (3.4-3.4) | 1.5 |
| Cameroon | Second administrative level | 79 | 197 (7-1883) | 17 (6-48) | 4 (3-38) | 1.4 (0-15.7) | 0 | | | | 76 | 4.2 (0-18.5) | 5.2 |
| Chad | Second administrative level | 28 | 488 (20-1986) | 15 (5-33) | 4 (3-19) | 3.05 (0.3-23.6) | 0 | | | | 27 | 2.9 (0-8.5) | 2.7 |
| Democratic Republic of the Congo | First administrative level | 13 | 1523 (74-15871) | 14 (3-37) | 3 (3-13) | 0.3 (0-5.6) | 0 | | | | 2 | 2.6 (1.2-4.1) | 0.2 |
| Democratic Republic of the Congo | Second administrative level | 229 | 188 (4-3529) | 15 (3-89) | 4 (3-45) | 1.3 (0-56.2) | 0 | | | | 227 | 1.4 (0-21.6) | 2.8 |

| | | | | | | | | | | | | | |
|---|---|---|---|---|---|---|---|---|---|---|---|---|---|
| Democratic Republic of the Congo | Third administrative level | 50 | 56<br>(8-662) | 14<br>(5-55) | 3<br>(3-49) | 5.4<br>(0.2-79.2) | 0 | | | | 50 | 0<br>(0-5.4) | 0.2 |
| Ethiopia | First administrative level | 9 | 1004<br>(48-7726) | 11<br>(6-21) | 5<br>(2-9) | 0.2<br>(0.1-2.3) | 9 | 12<br>(0-2751) | 0.00078<br>(0-0.0836) | 1<br>(0-0.83596) | 9 | 0.4<br>(0-6.6) | 0.5 |
| Ethiopia | Second administrative level | 66 | 293<br>(21-4255) | 12<br>(4-35) | 4<br>(2-25) | 0.2<br>(0-9.3) | 66 | 2<br>(0-1633) | 0.00012<br>(0-0.1258) | 1<br>(0-1.25802) | 66 | 0.5<br>(0-7.9) | 1 |
| Ethiopia | Third administrative level | 173 | 107<br>(5-2839) | 12<br>(3-35) | 4<br>(1-19) | 1<br>(0-117.1) | 140 | 0<br>(0-434) | 0<br>(0-0.83378) | 0<br>(0-8.33777) | 140 | 0<br>(0-11.4) | 0.7 |
| Ghana | First administrative level | 5 | 531<br>(178-3933) | 20<br>(4-24) | 5<br>(3-23) | 0.2<br>(0.1-0.9) | 1 | 79<br>(79-79) | 0.00189<br>(0.00189-0.00189) | 1<br>(0.01888-0.01888) | 5 | 0.8<br>(0-4.3) | 1.3 |
| Ghana | Second administrative level | 5 | 333<br>(148-1850) | 11<br>(7-19) | 3<br>(3-5) | 1<br>(0.5-1.2) | 5 | 1<br>(0-2) | 0.00011<br>(0-0.00037) | 1<br>(0-0.00374) | 5 | 0.7<br>(0.3-0.8) | 0.4 |
| Guinea | Second administrative level | 8 | 148<br>(21-4497) | 12<br>(5-15) | 4<br>(3-8) | 0.45<br>(0.1-2.9) | 0 | | | | 0 | | 0 |
| Guinea-Bissau | First administrative level | 1 | 633<br>(633-633) | 10<br>(10-10) | 6<br>(6-6) | 6.3<br>(6.3-6.3) | 0 | | | | 1 | 0.3<br>(0.3-0.3) | 0.3 |
| Guinea-Bissau | Second administrative level | 1 | 17<br>(17-17) | 5<br>(5-5) | 3<br>(3-3) | 0.5<br>(0.5-0.5) | 0 | | | | 1 | 0<br>(0-0) | 0 |

| | | | | | | | | | | | | | |
|---|---|---|---|---|---|---|---|---|---|---|---|---|---|
| Ivory Coast (Côte d'Ivoire) | Second administrative level | 1 | 22 (22-22) | 6 (6-6) | 3 (3-3) | 0 (0-0) | 0 | | | | 1 | 0 (0-0) | 0 |
| Ivory Coast (Côte d'Ivoire) | Third administrative level | 2 | 162 (128-197) | 9 (9-9) | 3 (3-3) | 1.15 (0.7-1.6) | 0 | | | | 2 | 5.9 (5.6-6.2) | 5.7 |
| Kenya | Second administrative level | 15 | 118 (20-1743) | 10 (5-18) | 3 (1-7) | 0.2 (0-0.9) | 1 | 14 (14-14) | 0.00112 (0.00112-0.00112) | 1 (0.01117-0.01117) | 15 | 0.7 (0-15) | 2.1 |
| Kenya | Third administrative level | 1 | 135 (135-135) | 10 (10-10) | 4 (4-4) | 0.9 (0.9-0.9) | 0 | | | | 1 | 10.4 (10.4-10.4) | 10.4 |
| Malawi | Second administrative level | 13 | 97 (25-388) | 10 (5-22) | 4 (3-8) | 0.2 (0-1) | 0 | | | | 13 | 3.3 (0-11.5) | 4 |
| Mali | Second administrative level | 3 | 59 (45-89) | 7 (7-8) | 3 (3-4) | 0.4 (0.4-0.4) | 0 | | | | 0 | | 0 |
| Mozambique | First administrative level | 1 | 1353 (1353-1353) | 13 (13-13) | 7 (7-7) | 0.2 (0.2-0.2) | 0 | | | | 0 | | 0 |
| Namibia | Second administrative level | 1 | 68 (68-68) | 12 (12-12) | 3 (3-3) | 1.2 (1.2-1.2) | 0 | | | | 0 | | 0 |
| Niger | Second administrative level | 24 | 160 (16-3314) | 9 (5-26) | 3 (3-21) | 0.45 (0-10.4) | 0 | | | | 20 | 2.5 (0-11.9) | 2.5 |

| | | | | | | | | | | | | | |
|---|---|---|---|---|---|---|---|---|---|---|---|---|---|
| Nigeria | First administrative level | 2 | 1020<br>(607-1433) | 20<br>(11-28) | 10<br>(4-17) | 0.15<br>(0.1-0.2) | 0 | | | | 1 | 3.3<br>(3.3-3.3) | 2.1 |
| Nigeria | Second administrative level | 138 | 148<br>(7-13642) | 10<br>(5-48) | 3<br>(3-46) | 0.55<br>(0-21.4) | 0 | | | | 123 | 1.8<br>(0-13.8) | 2.2 |
| Republic of the Congo | First administrative level | 1 | 439<br>(439-439) | 9<br>(9-9) | 3<br>(3-3) | 1<br>(1-1) | 0 | | | | 0 | | 0 |
| Republic of the Congo | Second administrative level | 2 | 142<br>(32-251) | 10<br>(10-11) | 4<br>(3-5) | 6.1<br>(1.7-10.5) | 2 | 142<br>(32-251) | 0.60878<br>(0.16823-1.04933) | 1<br>(1.68226-10.49331) | 2 | 9<br>(2.4-15.6) | 8.3 |
| Sierra Leone | First administrative level | 1 | 10052<br>(10052-10052) | 10<br>(10-10) | 4<br>(4-4) | 8.7<br>(8.7-8.7) | 0 | | | | 1 | 0.9<br>(0.9-0.9) | 0.9 |
| Sierra Leone | Second administrative level | 13 | 1025<br>(57-10650) | 9<br>(4-37) | 4<br>(3-8) | 2.5<br>(0.2-16.2) | 0 | | | | 10 | 2<br>(0-5.1) | 1.3 |
| Sierra Leone | Third administrative level | 16 | 202<br>(40-1006) | 13<br>(8-37) | 4<br>(2-10) | 4.05<br>(1.2-51.3) | 0 | | | | 0 | | 0 |
| Somalia | First administrative level | 2 | 519<br>(325-713) | 12<br>(3-21) | 4<br>(3-4) | 0.65<br>(0.3-1) | 0 | | | | 1 | 1<br>(1-1) | 0.4 |
| Somalia | Second administrative level | 2 | 721<br>(687-755) | 12<br>(9-16) | 4<br>(3-4) | 6.6<br>(3-10.2) | 0 | | | | 2 | 1.2<br>(0-2.3) | 1.8 |

| | | | | | | | | | | | | | |
|---|---|---|---|---|---|---|---|---|---|---|---|---|---|
| South Sudan | First administrative level | 10 | 1948 (560-4189) | 13 (6-23) | 4 (3-12) | 1.45 (0.4-3.9) | 8 | 6 (1-267) | 0.00056 (7e-05-0.03302) | 1 (0.00072-0.33018) | 8 | 1.9 (0.4-2.7) | 1.6 |
| South Sudan | Second administrative level | 20 | 596 (42-2112) | 13 (3-19) | 4 (3-10) | 2.5 (0.2-12) | 18 | 2 (0-185) | 0.00026 (0-0.13172) | 1 (0-1.31722) | 17 | 1.3 (0-20) | 2.7 |
| South Sudan | Third administrative level | 1 | 628 (628-628) | 6 (6-6) | 5 (5-5) | 45.2 (45.2-45.2) | 0 | | | | 0 | | 0 |
| Tanzania | First administrative level | 2 | 545 (491-599) | 14 (10-19) | 12 (10-14) | 0.35 (0.2-0.5) | 1 | 4 (4-4) | 0.00039 (0.00039-0.00039) | 1 (0.00389-0.00389) | 1 | 2.2 (2.2-2.2) | 0.7 |
| Tanzania | Second administrative level | 36 | 65 (8-550) | 11 (5-26) | 3 (2-13) | 0.2 (0-1.7) | 0 | | | | 36 | 0.5 (0-4.6) | 1 |
| Uganda | First administrative level | 10 | 92 (49-542) | 10 (6-27) | 3 (3-12) | 0.4 (0.1-2.3) | 0 | | | | 7 | 1.7 (0-5.9) | 0.6 |
| Zambia | First administrative level | 1 | 3314 (3314-3314) | 9 (9-9) | 5 (5-5) | 1.1 (1.1-1.1) | 0 | | | | 0 | | 0 |
| Zambia | Second administrative level | 1 | 3301 (3301-3301) | 9 (9-9) | 5 (5-5) | 1.5 (1.5-1.5) | 0 | | | | 0 | | 0 |
| Zimbabwe | Third administrative level | 2 | 716 (254-1178) | 6 (6-7) | 3 (3-3) | 40.95 (18.1-63.8) | 0 | | | | 0 | | 0 |

**Table S2 Total and proportion of person-months of population living in outbreak areas by country in sub-Saharan Africa**

This table presents the number and percent of person-months of the total population living in outbreak regions by country from January 2010 to January 2020.

| Country | Total person-months | Proportion of person-months of total population living in outbreak regions (%) |
|---|---|---|
| Angola | 3,137,465,312 | 0.09 |
| Benin | 1,288,925,415 | 0.88 |
| Ivory Coast (Côte d'Ivoire) | 2,790,240,062 | 0.36 |
| Cameroon | 2,852,428,474 | 4.13 |
| Democratic Republic of the Congo | 10,678,222,272 | 4.86 |
| Republic of the Congo | 437,763,585 | 0.33 |
| Ethiopia | 10,899,055,168 | 4.65 |
| Ghana | 3,407,049,340 | 2.2 |
| Guinea | 1,295,831,664 | 1.06 |
| Guinea-Bissau | 198,652,882 | 0.18 |
| Kenya | 5,592,561,388 | 0.88 |
| Mali | 2,225,476,318 | 0.07 |
| Mozambique | 3,123,348,768 | 0.7 |

| | | |
|---|---|---|
| Malawi | 1,905,777,660 | 1.63 |
| Namibia | 270,098,059 | 0.06 |
| Niger | 2,322,205,822 | 1.54 |
| Nigeria | 22,194,150,048 | 1.15 |
| Sierra Leone | 739,725,332 | 2.75 |
| Somalia | 1,159,854,920 | 0.58 |
| South Sudan | 1,425,871,411 | 4.16 |
| Chad | 1,642,708,449 | 1.25 |
| Tanzania | 4,013,605,452 | 2 |
| Uganda | 4,309,672,400 | 0.37 |
| Zambia | 1,862,447,024 | 0.49 |
| Zimbabwe | 1,629,500,824 | 0.01 |

**Table S3. Summary of confirmed cholera cases in outbreaks reported in sub-Saharan Africa, 2010-2020**
This table presents the summary of confirmed cholera cases in outbreaks by different administrative reporting units.

| | Outbreaks reported at the first-level administrative units | Outbreaks reported at the second-level administrative units | Outbreaks reported at the third-level administrative units |
|---|---|---|---|
| Number of outbreaks with non-missing confirmed case data;at least one confirmed case | 19;18 | 92;54 | 140;56 |
| Median number of confirmed cases^ | 10;11 | 2;6 | 0;6 |

| | | | |
|---|---|---|---|
| (IQR) | (3-28);(4-32) | (0-8);(2-18) | (0-4);(2-34) |
| Percentage of suspected cases that were confirmed^ (%)(IQR) | 0.8;0.9<br>(0.2-4.8)/(0.3-5.3) | 0.4;2.2<br>(0-4.4);(0.5-9.8) | 0;9.0<br>(0-4.3);(2.2-29.2) |
| Median confirmed cases per 1,000 people per outbreak (IQR)[a] | 0.01;0.01<br>(0.002-0.02)/(0.002-0.02) | 0.001;0.01<br>(0-0.01)/(0.002-0.04) | 0;0.07<br>(0-0.04)/(0.02-0.3) |

^ Among outbreaks with any confirmed case data ; Among outbreaks with at least one confirmed case
[a]Only outbreaks with at least one confirmed case and valid population estimates are included. There were 17 outbreaks at first-level administrative units, 54 outbreaks at second-level units and 56 outbreaks at third-level units.

**Table S4 Outbreaks reported at first, second, and third administrative levels in rural and urban settings.**
This table presents the comparisons of epidemic metrics between rural and urban settings across different administrative reporting units. Only outbreaks reported at the first-, and second-level administrative units are included here.

| | Outbreaks reported at the first-level administrative units | | Outbreaks reported at the second-level administrative units | |
|---|---|---|---|---|
| | Rural | Urban | Rural | Urban |
| Metrics based on suspected cholera cases | | | | |
| Number of outbreaks | 55 | 7 | 558 | 134 |
| Outbreak threshold (weekly incidence per 100,000 people) (IQR) | 0.7<br>(0.2-2.0) | 1.2<br>(0.6-4.6) | 0.7<br>(0.3-2.7) | 0.7<br>(0.2-4.0) |
| Median outbreak size (IQR) | 607<br>(211-1,948) | 3,715<br>(382-5,830) | 182<br>(74-448) | 190<br>(62-461) |
| Median epidemic durations, weeks (IQR) | 12<br>(8-20) | 10<br>(8-14) | 13*<br>(8-18) | 14*<br>(8-23) |

| | | | | |
|---|---|---|---|---|
| Median time to epidemic peak, weeks (IQR) | 4<br>(4-7) | 3<br>(3-5) | 3<br>(3-6) | 4<br>(3-7) |
| Median proportion of suspected cases reported during the peak week (%) (IQR) | 21.2<br>(15.2-30) | 18.5<br>(15.1-19.4) | 24.6*<br>(17-35.6) | 20.4*<br>(15.6-30.9) |
| Median weekly incidence during the peak week per 1,000 people (IQR) | 0.1<br>(0.03-0.2) | 0.2<br>(0.1-0.3) | 0.2*<br>(0.1-0.5) | 0.1*<br>(0.04-0.5) |
| Median early outbreak reproductive number (range) | 1.8<br>(1.2-2.9) | 2.0<br>(1.5-2.8) | 1.9<br>(1.1-3.4) | 1.9<br>(1.3-3.5) |
| Median attack rate per 1,000 people (IQR) | 0.4<br>(0.1-1.2) | 0.9<br>(0.6-1.7) | 0.9*<br>(0.3-2.5) | 0.6*<br>(0.2-2.2) |
| Metrics based on cholera-associated deaths[a] | | | | |
| Number of outbreaks with reports of deaths | 32 | 5 | 529 | 117 |
| Median case fatality risk (%)(IQR) | 1.4*<br>(0.4-2.3) | 0.8*<br>(0.2-0.8) | 1.8*<br>(0.5-4.3) | 0.8*<br>(0-2.7) |

**Table S5. Outbreaks reported at first, second, and third administrative levels in rural and urban settings.**
This table presents the comparisons of confirmed cholera cases between rural and urban settings across different administrative reporting units. Only outbreaks reported at the first-, second- and third-level administrative units are included here.

| | Outbreaks reported at the first-level administrative units | | Outbreaks reported at the second-level administrative units | | Outbreaks reported at the third-level administrative units | |
|---|---|---|---|---|---|---|
| | Rural | Urban | Rural | Urban | Rural | Urban |
| Number of outbreaks reporting at least one | 16 | 2 | 49 | 5 | 39 | 17 |

| | | | | | | |
|---|---|---|---|---|---|---|
| confirmed case | | | | | | |
| Median number of confirmed cases per outbreak (IQR) | 8* (4-17) | 1,415* (747-2,083) | 6 (4-23) | 2 (1-11) | 4* (2-9) | 216* (11-266) |
| Percentage of suspected cases that were confirmed (%) (IQR) | 0.6 (0.2-4.3) | 18.8 (10.4-27.2) | 2.5 (0.6-10.1) | 0.3 (0.3-2.5) | 5.1* (1.7-13.7) | 31.7* (14.8-41.6) |
| Median confirmed cases per 1,000 people per outbreak (IQR) | 0.01 (0.002-0.02) | 0.4 (0.2-0.6) | 0.01 (0.002-0.05) | 0.004 (0.003-0.01) | 0.04* (0.02-0.1) | 0.7* (0.3-0.9) |

* P<0.05 Wilcoxon rank sum test was used to test if the medians of outbreak characteristics are the same between rural and urban settings.

[a]N.B. Outbreaks with reports of confirmed cases may not have documented this information systematically, so these results are highly sensitive to reporting biases.

**Table S6. Summary of cholera outbreaks reported in sub-Saharan Africa, 2010-2020**

This table presents the key epidemic metrics of outbreaks by different administrative reporting units, including outbreak size, duration, time to outbreak peak, initial reproductive numbers during the first epidemic week, attack rate, proportion of confirmed cases, and CFRs. These outbreaks were extracted using the new definition in sensitivity analysis.

| | Outbreaks reported at the first-level administrative units | Outbreaks reported at the second-level administrative units | Outbreaks reported at the third-level administrative units | Outbreaks reported at the fourth-level administrative units or lower |
|---|---|---|---|---|
| Metrics based on suspected cholera cases | | | | |
| Number of outbreaks | 113 | 1,086 | 399 | 8 |
| Median outbreak size (IQR) | 526 (164-1,665) | 128 (47-317) | 57 (26-156) | 54 (30-188) |

| | | | | |
|---|---|---|---|---|
| Median epidemic duration in weeks (IQR) | 10<br>(6-18) | 8<br>(5-13) | 7<br>(5-11) | 5<br>(5-7) |
| Median weeks to outbreak peak (IQR) | 5<br>(3-10) | 4<br>(3-6) | 4<br>(3-5) | 3<br>(3-5) |
| Median proportion of suspected cases reported during the peak week (%) (IQR) | 24.5<br>(15.8-33.9) | 28.3<br>(18.7-40) | 34<br>(23.8-43.4) | 38.9<br>(16.6-42.5) |
| Median weekly incidence during the peak week per 1,000 people (IQR)[b] | 0.04<br>(0.01-0.1) | 0.2<br>(0.1-0.4) | 0.5<br>(0.2-1.4) | 0.6<br>(0.4-0.8) |
| Median early outbreak reproductive number (range) | 1.9<br>(1.4-5) | 1.9<br>(1.1-4.5) | 2<br>(1-3.6) | 1.9<br>(1.5-2.2) |
| Median attack rate per 1,000 people (IQR)[b] | 0.2<br>(0.06-1.0) | 0.6<br>(0.2-1.8) | 1.6<br>(0.5-4.8) | 1<br>(0.7-1.4) |
| Metrics based on confirmed cholera cases[a] | | | | |
| Number of outbreaks with non-missing confirmed case data;at least one confirmed case | 47;22 | 157;70 | 215;59 | 4;1 |
| Median number of confirmed cases^ (IQR) | 0;14<br>(0-14);(4-35) | 0;7<br>(0-6);(3-18) | 0;6<br>(0-1);(2-34) | 0;1 |
| Percentage of suspected cases that were confirmed^ (%) | 0;2.4<br>(0-2.3);(0.6-9.5) | 0;2.5<br>(0-1.7);(0.7-10.1) | 0;9.7<br>(0-0.9);(2.4-29.6) | 0;11.1<br>(0-2.8);(11.1-11.1) |
| Median confirmed cases per 1,000 people per outbreak (IQR)[cd] | 0;0.005<br>(0-0.005);(0.002-0.02) | 0;0.009<br>(0-0.008)/(0.003-0.04) | 0;0.1<br>(0-0.02)/(0.02-0.6) | - |
| Metrics based on cholera-associated deaths[a] | | | | |

| | | | | |
|---|---|---|---|---|
| Number of outbreaks with reports of deaths | 73 | 1,008 | 353 | 8 |
| Median case fatality risk (%)(IQR) | 1.4<br>(0.4-2.9) | 1.6<br>(0-4.1) | 0<br>(0-0.2) | 0.2<br>(0-2.6) |
| Population-Weighted case fatality risk (%)[e] | 1.1 | 1.9 | 0.7 | 9.5 |

- This value could not be reported due to missing information.
^ Among outbreaks with any confirmed case data / Among outbreaks with at least one confirmed case
[a]N.B. Outbreaks with reports of confirmed cases and deaths may not have documented this information systematically, so these results are highly sensitive to reporting biases.
[b]Only outbreaks with valid population estimates are included. There were 110 outbreaks at the first-level administrative units, 1036 outbreaks at the second-level units, 324 outbreaks at the third-level units and 2 outbreaks at the fourth-level units or lower.
[C]Only outbreaks with confirmed cases with valid population estimates are included. There were 47 outbreaks at the first-level administrative units, 152 outbreaks at the second-level units, and 193 outbreaks at the third-level units.
[d]Only outbreaks with at least one confirmed case and valid population estimates are included. There were 22 outbreaks at first-level administrative units, 70 outbreaks at second-level units and 59 outbreaks at third-level units.
[e]Only outbreaks with reports of deaths and valid population estimates are included. There were 73 outbreaks at the first-level administrative units, 972 outbreaks at the second-level units, 280 outbreaks at the third-level units and 2 outbreaks at the fourth-level units or lower.

**Table S7. Outbreaks reported at first, second, and third administrative levels in rural and urban settings.**
This table presents the comparisons of epidemic metrics between rural and urban settings across different administrative reporting units. Only outbreaks reported at the first-, second- and third-level administrative units are included here. These outbreaks were extracted using the definition in the sensitivity analysis.

| | Outbreaks reported at the first-level administrative units | | Outbreaks reported at the second-level administrative units | | Outbreaks reported at the third-level administrative units | |
|---|---|---|---|---|---|---|
| Setting | Rural | Urban | Rural | Urban | Rural | Urban |
| Metrics based on suspected cholera cases | | | | | | |
| Number of outbreaks | 103 | 7 | 824 | 212 | 242 | 82 |
| Median outbreak size (IQR) | 490*<br>(136-1,250) | 4,258*<br>(610-6,988) | 140*<br>(57-343) | 114*<br>(35-270) | 80<br>(32-158) | 49<br>(24-220) |

| | | | | | | |
|---|---|---|---|---|---|---|
| Median epidemic durations, weeks (IQR) | 10 (6-16) | 12 (10-20) | 8 (5-13) | 8 (5-14) | 7 (5-10) | 7 (5-12) |
| Median time to epidemic peak, weeks (IQR) | 5 (3-9) | 8 (4-12) | 4 (3-6) | 4 (3-7) | 4 (3-5) | 3 (3-6) |
| Median proportion of suspected cases reported during the peak week (%) (IQR) | 26.6* (16.6-34.4) | 15* (11.7-18.2) | 28.2 (18.8-40) | 27.6 (18-38.8) | 35.6* (25-43.4) | 30.8* (21.6-41.3) |
| Median weekly incidence during the peak week per 1,000 people (IQR) | 0.04* (0.01-0.1) | 0.2* (0.1-0.3) | 0.2* (0.1-0.4) | 0.1* (0.04-0.3) | 0.4* (0.1-1.3) | 0.6* (0.2-2.9) |
| Median early outbreak reproductive number (range) | 1.9 (1.4-5) | 1.9 (1.5-2.8) | 1.9 (1.1-4.5) | 1.9 (1.2-3.6) | 2 (1-3.6) | 2 (1.5-3.2) |
| Median attack rate per 1,000 people (IQR) | 0.2* (0.06-0.8) | 1.6* (0.7-2.1) | 0.7* (0.2-1.8) | 0.4* (0.1-1.6) | 1.4* (0.4-4.1) | 2.4* (0.9-9.0) |
| Metrics based on confirmed cholera cases[a] | | | | | | |
| Number of outbreaks reporting at least one confirmed case | 20 | 2 | 63 | 7 | 41 | 18 |
| Median number of confirmed cases per outbreak (IQR) | 14* (4-23) | 1,350* (728-1,972) | 6 (3-18) | 15 (6-43) | 4* (2-8) | 197* (12-251) |

| | | | | | | |
|---|---|---|---|---|---|---|
| Percentage of suspected cases that were confirmed (%) | 2.4 (0.5-8.6) | 18.4 (10-26.7) | 2.5 (0.7-10) | 3.6 (1.2-14.5) | 6.5* (1.7-14.9) | 30.9* (16.8-40.8) |
| Median confirmed cases per 1,000 people per outbreak (IQR) | 0.005 (0.001-0.02) | 0.4 (0.2-0.6) | 0.008 (0.003-0.04) | 0.03 (0.008-0.05) | 0.04* (0.02-0.09) | 0.7* (0.3-0.8) |
| Metrics based on cholera-associated deaths[a] | | | | | | |
| Number of outbreaks with reports of deaths | 68 | 5 | 786 | 186 | 203 | 77 |
| Median case fatality risk (%)(IQR) | 1.6 (0.5-3.2) | 0.7 (0.2-0.8) | 1.8* (0.1-4.4) | 0.9* (0-3.2) | 0 (0-0.07) | 0 (0-0) |

* P<0.05

[a]N.B. Outbreaks with reports of confirmed cases and deaths may not have documented this information systematically, so these results are highly sensitive to reporting biases.

**Table S8. Summary of cholera outbreaks reported in sub-Saharan Africa, 2010-2020**

This table presents the key epidemic metrics of outbreaks by different population sizes, including outbreak size, duration, time to outbreak peak, initial reproductive numbers during the first epidemic week, attack rate, proportion of confirmed cases, and CFRs.

| Population | <10,000 | 10,000-100,000 | 100,000-1,000,000 | >=1,000,000 |
|---|---|---|---|---|
| Metrics based on suspected cholera cases | | | | |
| Number of outbreaks | 33 | 246 | 624 | 96 |
| Median outbreak size (range) | 41 (33-92) | 153 (58-424) | 156 (65-421) | 437[a] (140-1493) |

| | | | | |
|---|---|---|---|---|
| Median epidemic durations (weeks)(IQR) | 15<br>(9-22) | 12<br>(9-18) | 12[ab]<br>(8-17) | 12[c]<br>(9-18) |
| Median time to epidemic peak (weeks)(IQR) | 3<br>(3-6) | 3<br>(3-5) | 4[b]<br>(3-6) | 4<br>(3-6) |
| Median proportion of suspected cases reported during the peak week (%) (IQR) | 22<br>(17.5-30) | 26<br>(17.9-35.9) | 24.8[ab]<br>(17.2-36.3) | 22[c]<br>(15.8-33.1) |
| Median weekly incidence during the peak week per 1,000 people (IQR) | 2.57<br>(1.25-5.47) | 0.7<br>(0.3-1.7) | 0.2[b]<br>(0.1-0.4) | 0.04[a]<br>(0.01-0.1) |
| Median early outbreak reproductive number (range) | 1.9<br>(1.5-2.3) | 1.9<br>(1.1-2.8) | 1.8<br>(1.02-3.5) | 1.8(1.2-3) |
| Median attack rate per 1,000 people (IQR) | 9.9<br>(5.2-30.9) | 2.95[a]<br>(1.2-9.3) | 0.7<br>(0.3-1.7) | 0.2[a]<br>(0.1-0.5) |
| Metrics based on confirmed cholera cases | | | | |
| Number of outbreaks with any confirmed case data / at least one confirmed case | 2;1 | 59;27 | 139;66 | 51;34 |
| Median number of confirmed cases^ (IQR) | 24;47<br>(12-35);47 | 0;6<br>(0-6);(4-14) | 0[ab];6[ab]<br>(0-5);(2-29) | 2[c];6<br>(0-13);(2-22) |
| Percentage of suspected cases that were confirmed^ (%) | 34.6;69.1<br>(17.3-51.8);(69.1-69.1) | 0;8.2272<br>(0-6.6);(2.1-17.8) | 0[ab];4.3[ab]<br>(0-3.8);(0.9-16.3) | 0.4[c];1.4<br>(0-4.3);(0.3-7.5) |
| Median confirmed cases per 1,000 people (%) per outbreak^ (IQR) | 4.2;8.3<br>(2.1-6.3);(8.3-8.3) | 0;0.1<br>(0-0.1);(0.05-0.3) | 0[ab];0.02[ab]<br>(0-0.02);(0.008-0.08) | 0.001[c];0.003[b]<br>(0-0.006);(0.001-0.01) |

| | | | | |
|---|---|---|---|---|
| | Metrics based on cholera-associated deaths | | | |
| Number of outbreaks with reports of deaths | 32 | 200 | 568 | 76 |
| Median case fatality risk (%)(IQR) | 0<br>(0-0) | 0.9[a]<br>(0-3.2) | 1.4[ab]<br>(0-3.5) | 0.8[abc]<br>(0.1-1.7) |
| Population-Weighted case fatality risk (%) | 0.5 | 2 | 2.2 | 0.8 |

[a] P<0.05 between regions with less than 10,000 people and groups with larger population sizes
[b] P<0.05 between regions with 10,000-100,000 people and groups with larger population sizes
[c] P<0.05 between regions with 100,000-1,000,000 people and groups with larger population sizes
^ Among outbreaks with any confirmed case data / Among outbreaks with at least one confirmed case

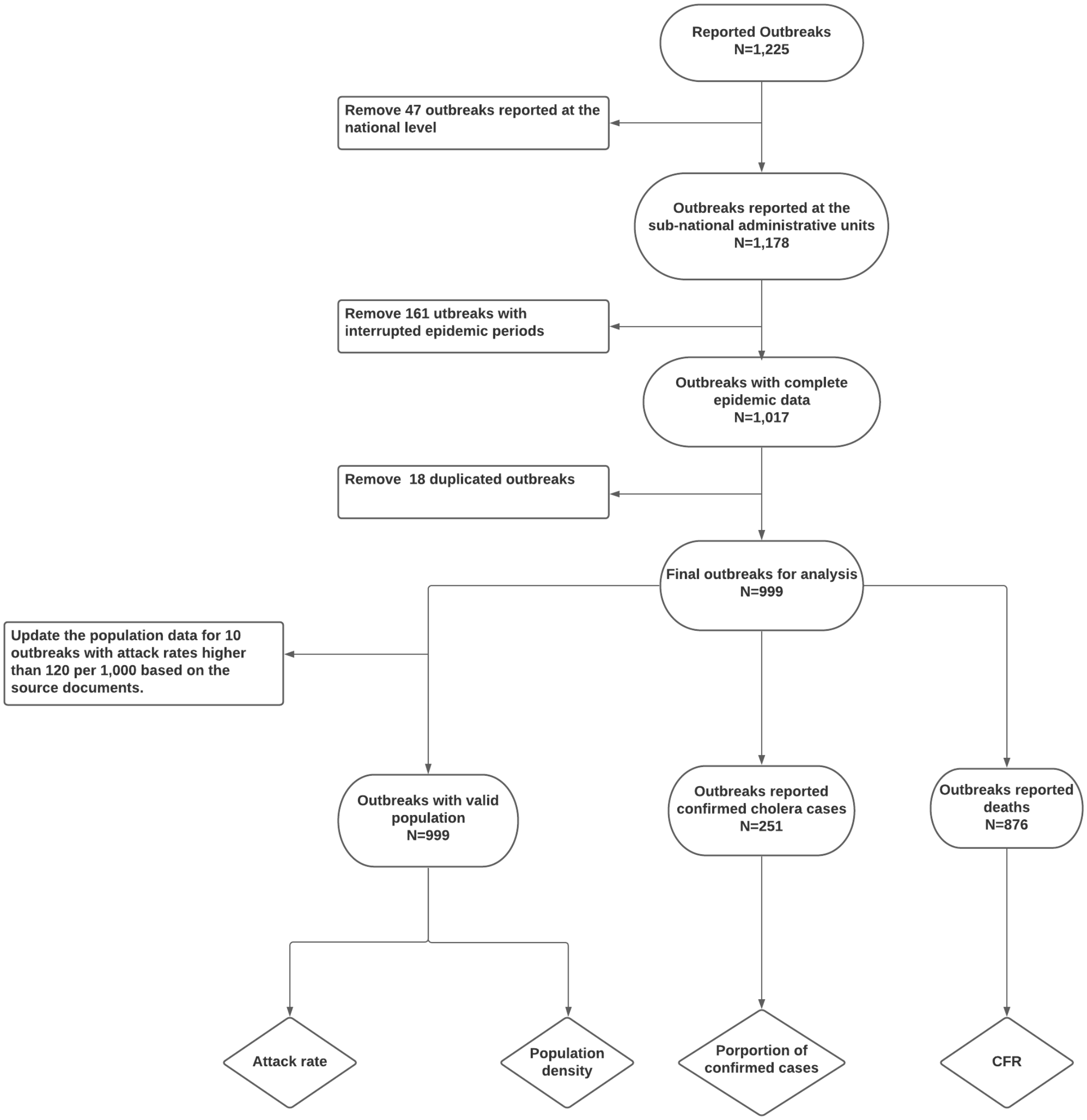


**Figure S1** Outbreak selection overview in the main analysis

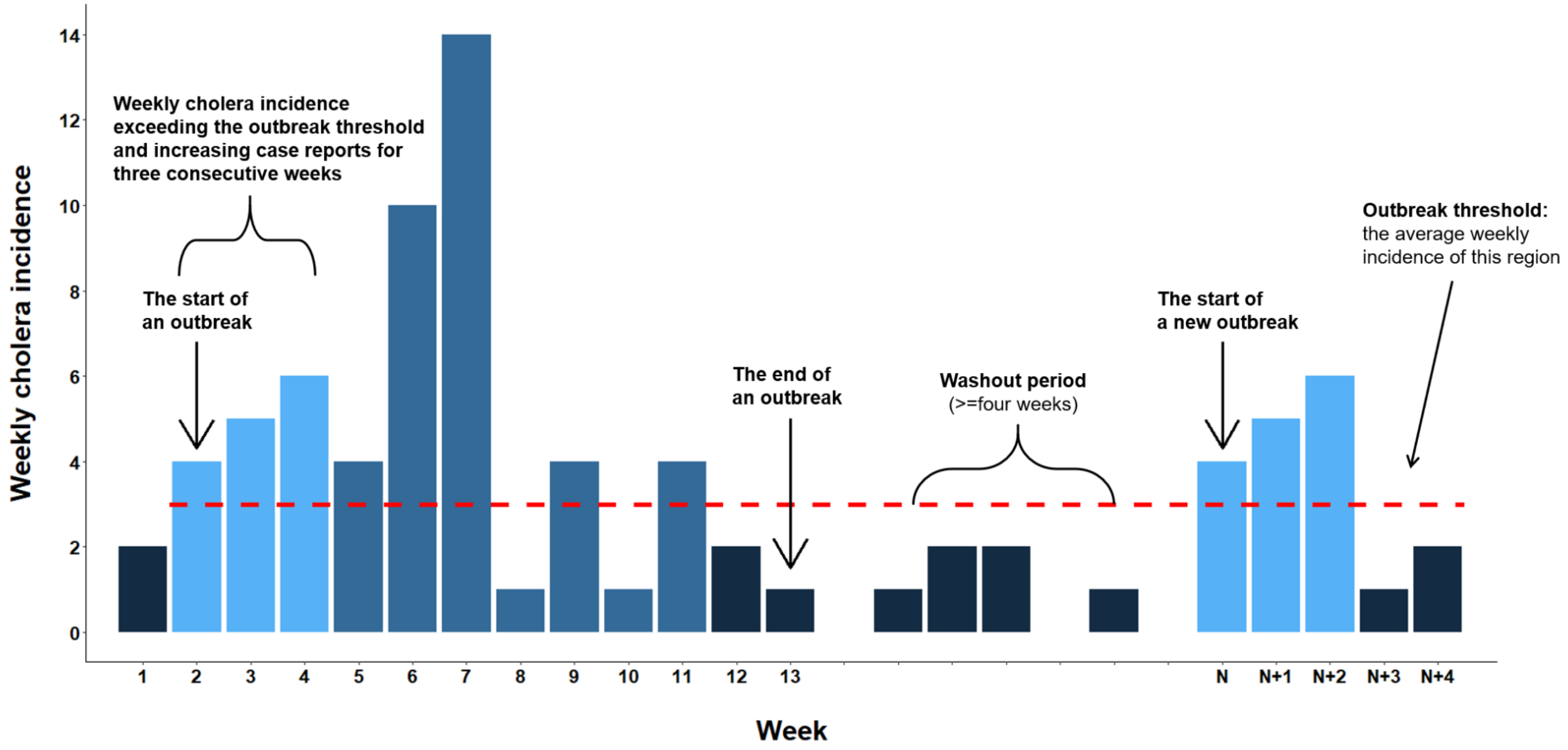


**Figure S2 Schematic of the cholera outbreak definition**

This conceptual diagram displays our systematic application of the cholera outbreak definition for cholera incidence time series in a given region. Light blue bars show the ascending phase of an outbreak which indicates an outbreak start. Dark blue bars indicate the *outbreak end* and *washout period*, all of which remain below the outbreak-specific outbreak threshold (red dashed line). An outbreak's *epidemic period* is defined as the period from the outbreak start to the last week with weekly incidence exceeding the outbreak threshold, displayed in steel blue. Reported cholera incidence and mortality from the first epidemic week through the last day of the outbreak end are included in the outbreak summaries.

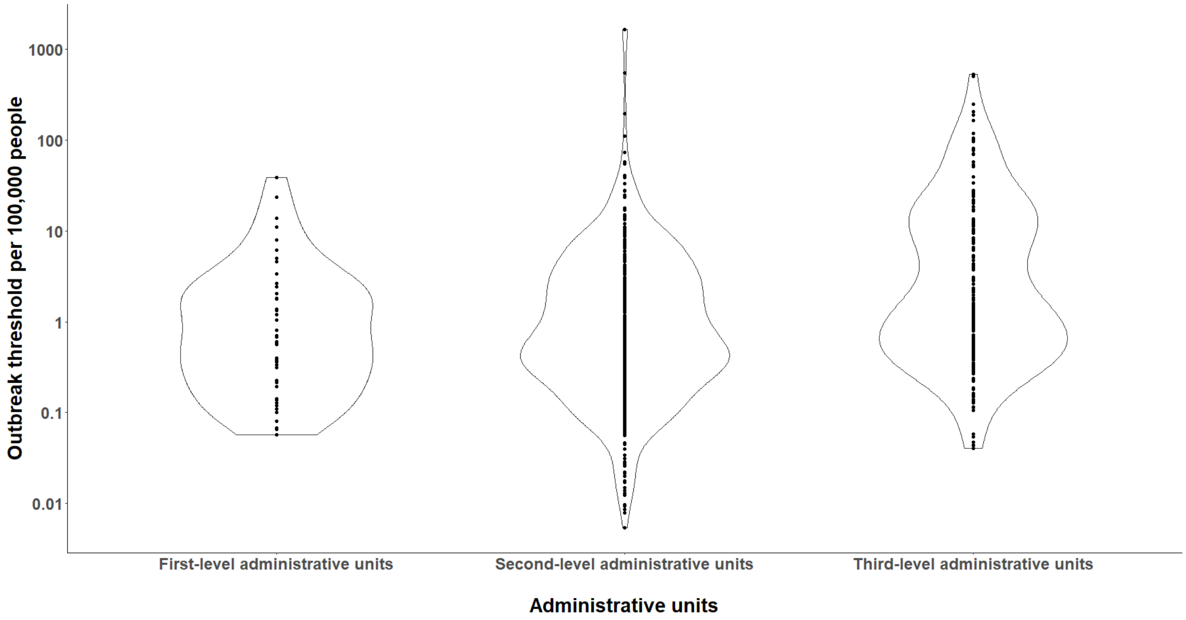


**Figure S3. Distributions of outbreak threshold per 100,000 people.** This figure shows the distributions of attack rates per 1,000 at different spatial reporting scales.

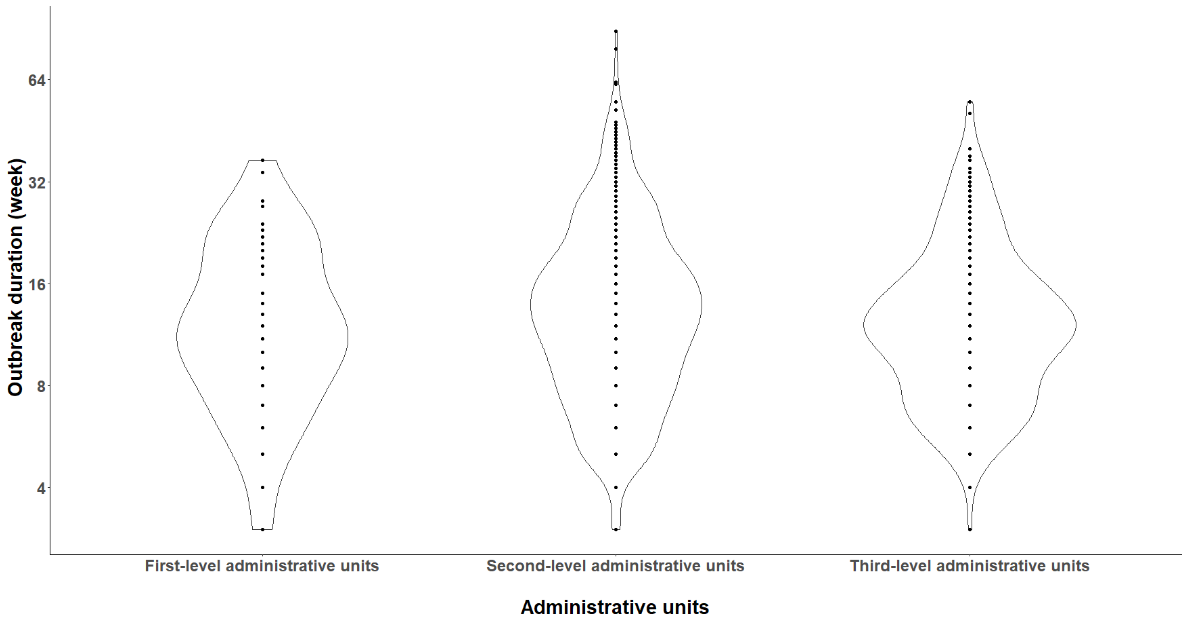


**Figure S4. Distributions of outbreak durations (weeks)**

This figure shows the distributions of outbreak duration in weeks at different spatial reporting units.

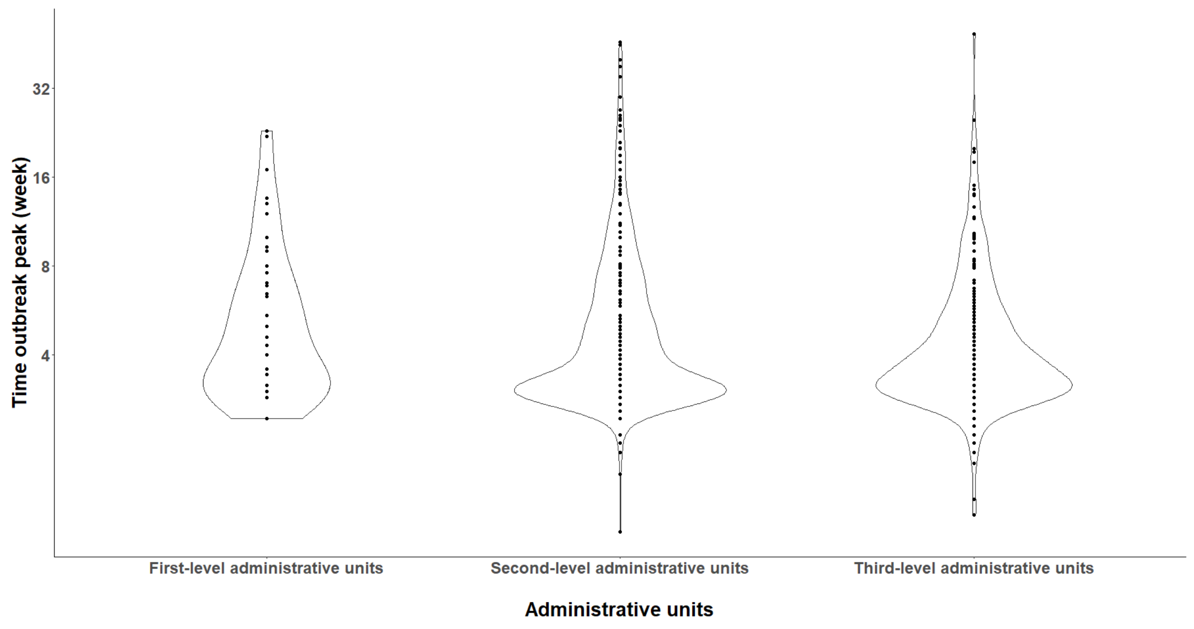


**Figure S5. Distributions of time to outbreak peak (weeks)**
This figure shows the distributions of time to outbreak peak in weeks at different spatial reporting units.

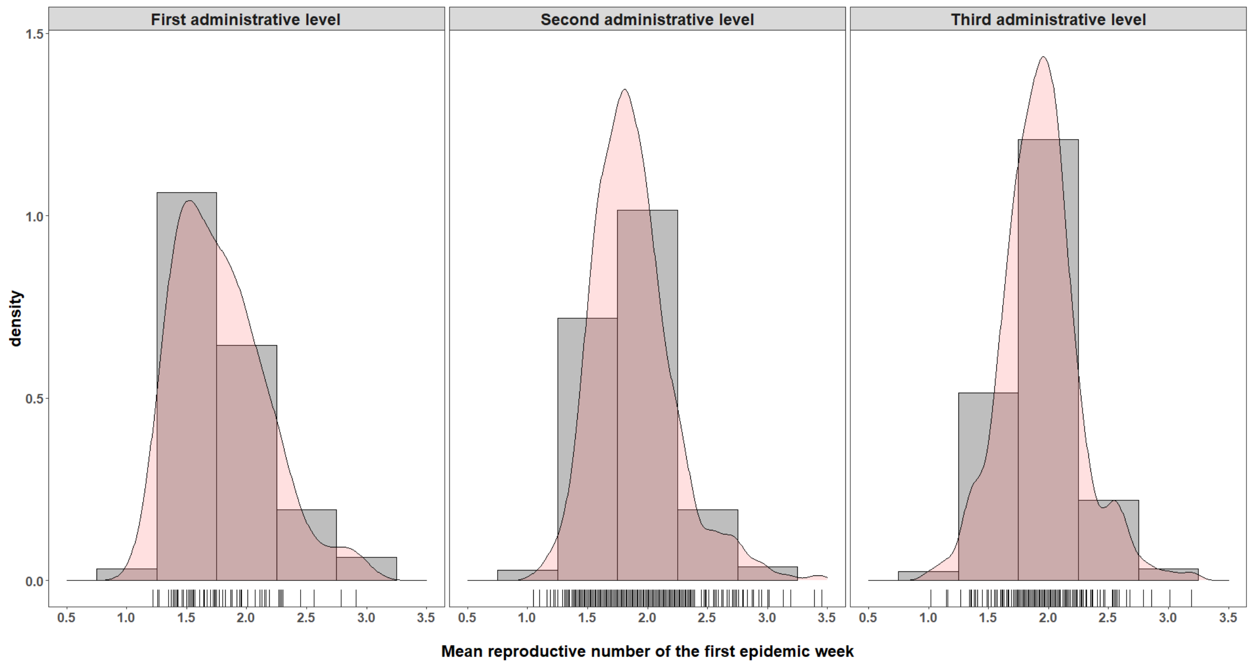


**Figure S6. Reproductive numbers of the first epidemic week distribution**
This figure shows the distributions of average reproductive numbers during the first epidemic week at different spatial reporting units.

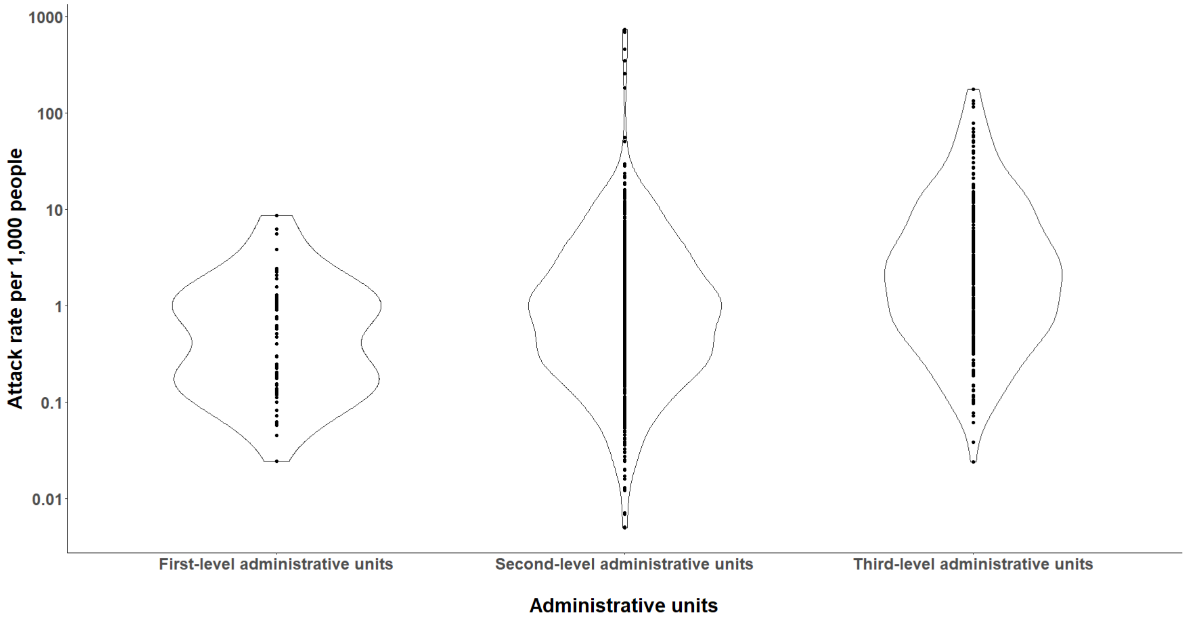


**Figure S7. Distributions of attack rates per 1,000 people**
This figure shows the distributions of attack rates per 1,000 across different spatial reporting units.

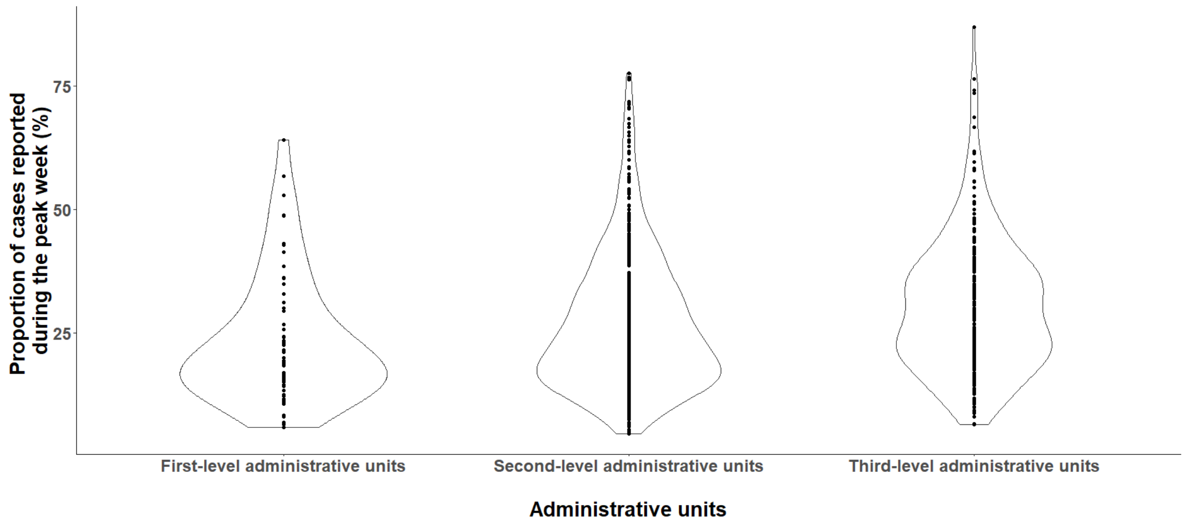


**Figure S8 Distribution of proportion of cases reported during the peak week**
This figure shows the distribution of the proportion of cases reported during the peak week (%).

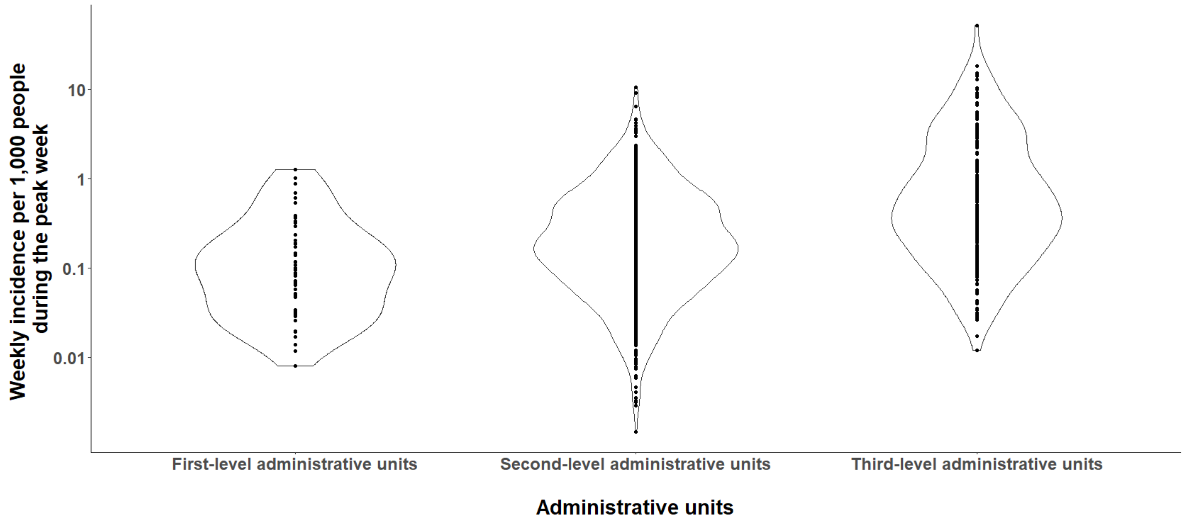


**Figure S9 Distribution of weekly incidence per 1,000 people during the peak week**
This figures shows the weekly incidence of the peak week (per 1,000 people)

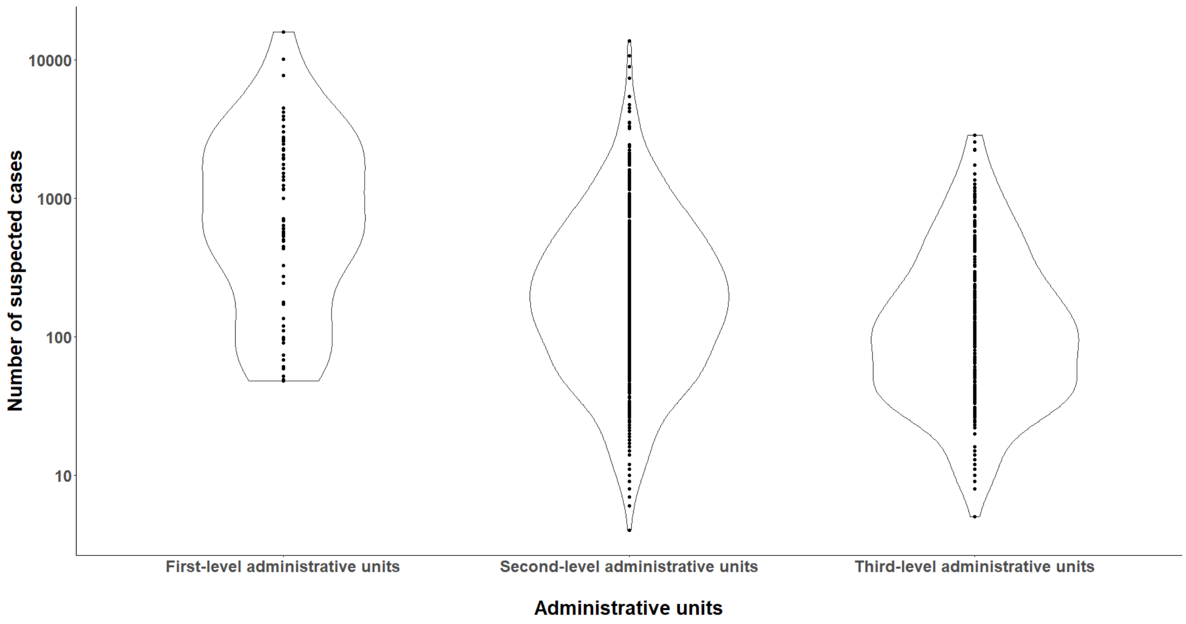


**Figure S10. Distributions of number of suspected cases per outbreak**
This figure shows the distributions of the number of suspected cases per outbreak across different spatial reporting units.

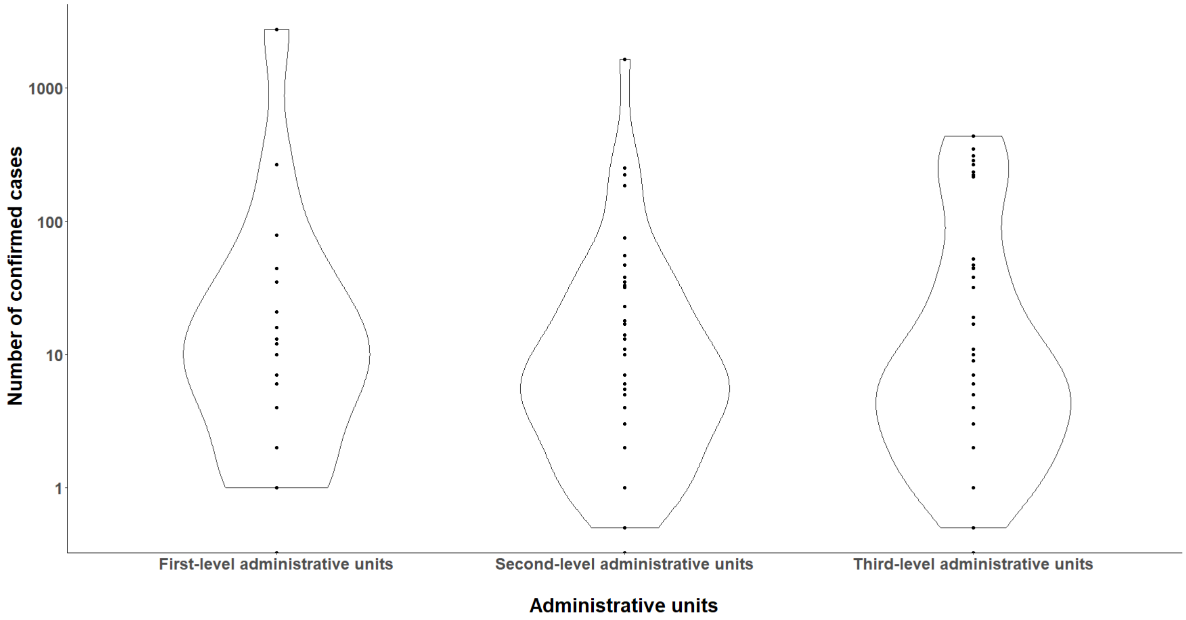


**Figure S11. Distributions of the number of confirmed cases stratified by administrative units**

This figure shows the distributions of the number of confirmed cases across different spatial reporting units.

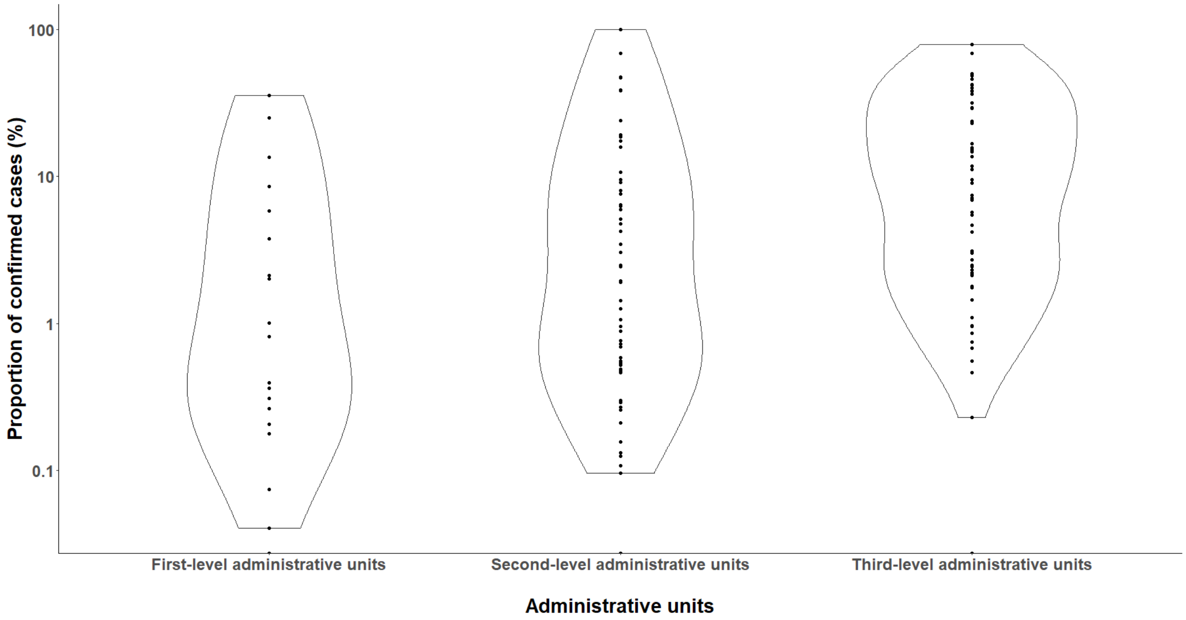


**Figure S12. Distributions of the proportion of confirmed cases stratified by administrative units**

This figure shows the distributions of the proportion of confirmed cases across rural and urban settings at different spatial reporting units.

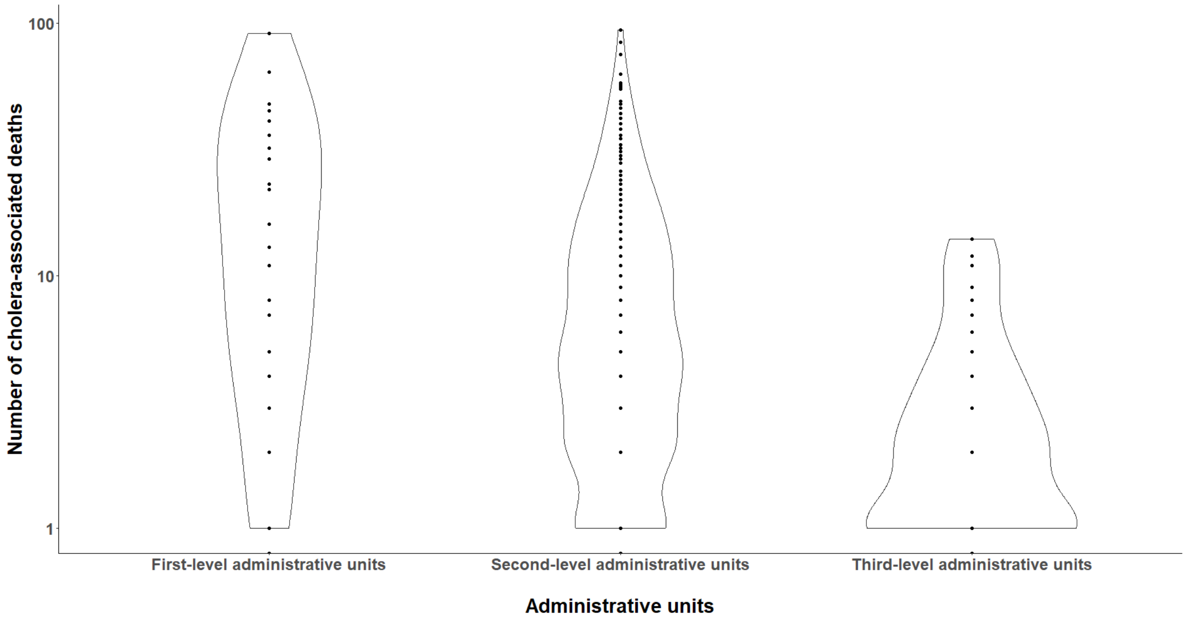


**Figure S13. Distributions of the number of cholera-associated deaths stratified by administrative units**

This figure shows the distributions of the number of cholera-associated deaths across different spatial reporting units.

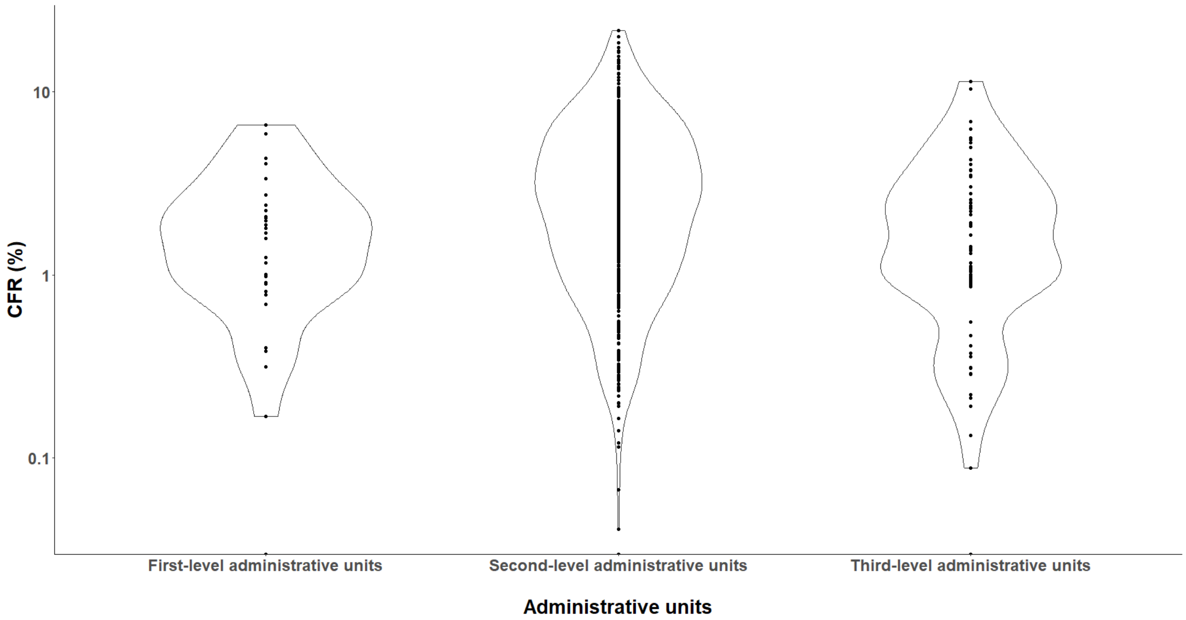

### Figure S14. Distributions of CFR (%) stratified by administrative units

This figure shows the distributions of CFR (%) across rural and urban settings at different spatial reporting units.

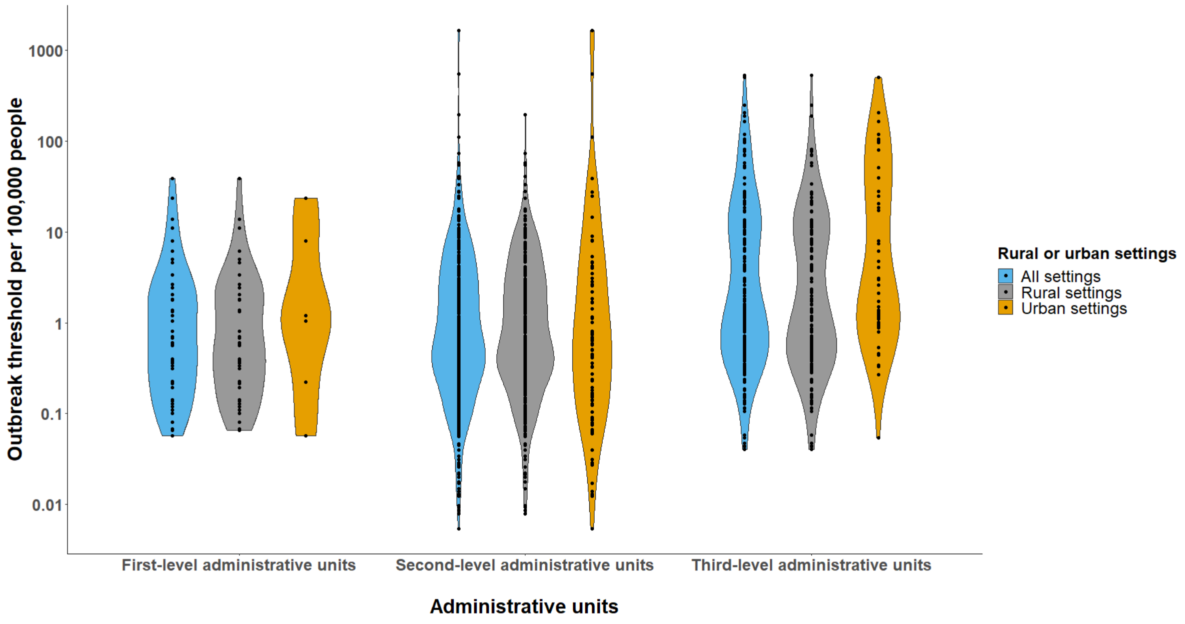


### Figure S15. Distributions of outbreak threshold stratified by rural and urban settings

This figure shows the distributions of outbreak threshold (weekly incidence per 100,000 people) across rural and urban settings at different spatial reporting units. Only outbreaks reported at the first-, second- and third-level administrative units are included.

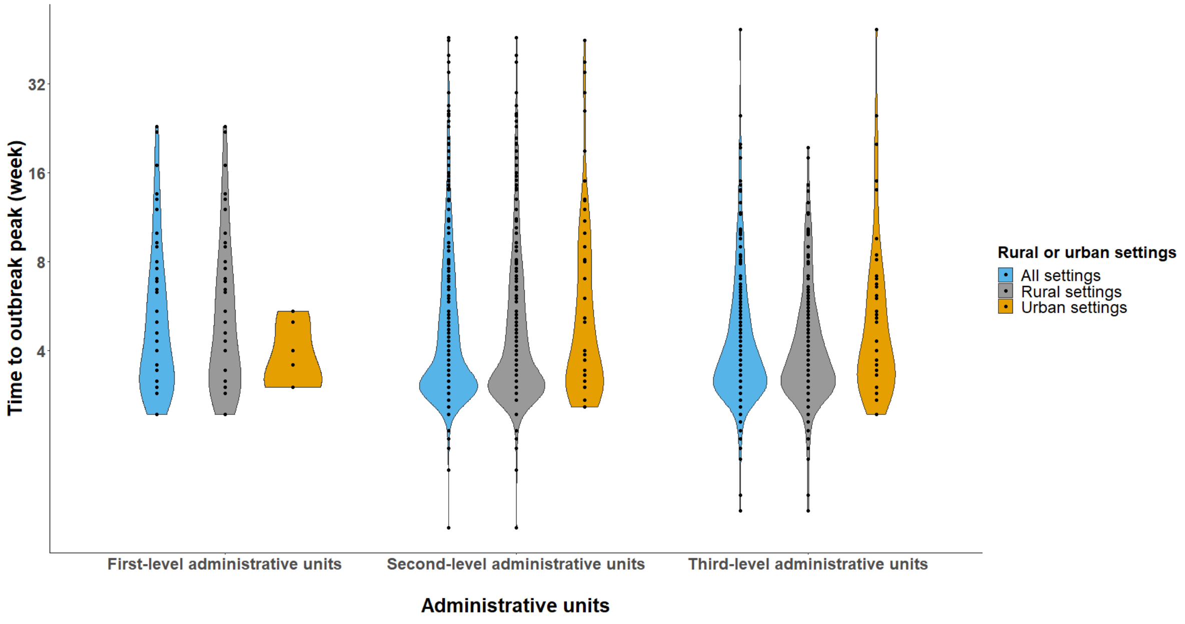

**Figure S16. Distributions of time to outbreak peak (weeks) stratified by rural and urban settings**

This figure shows the distributions of time to outbreak across rural and urban settings at different spatial reporting units. Only outbreaks reported at the first-, second- and third-level administrative units are included.

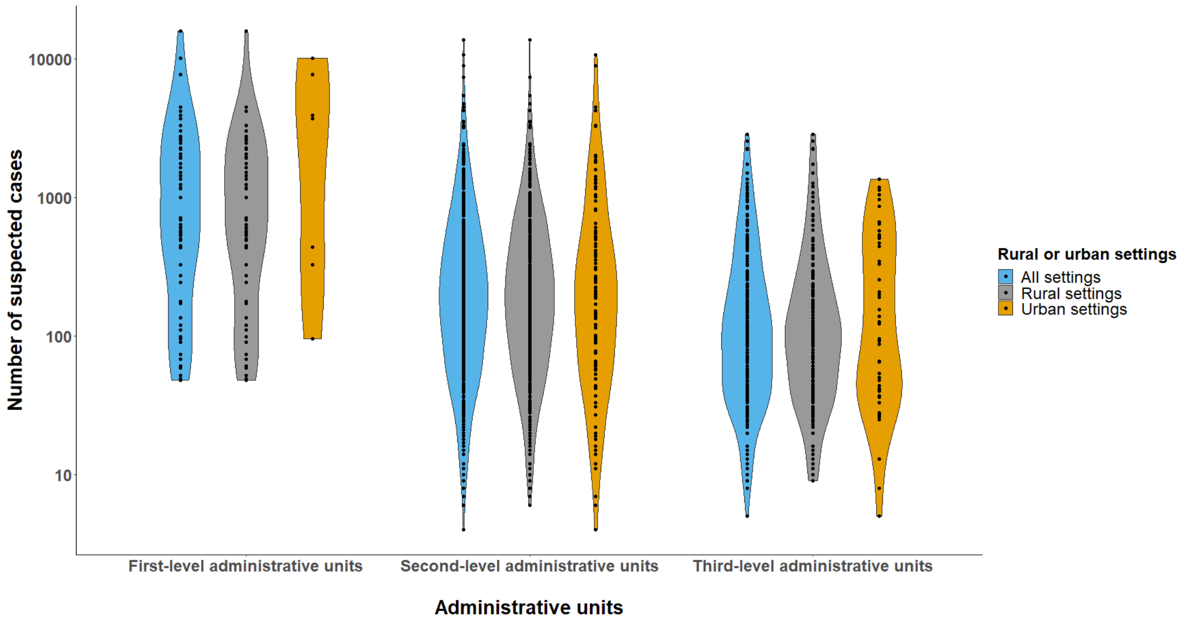


**Figure S17. Distribution of outbreak sizes stratified by rural and urban settings**

This figure shows the distributions of outbreak sizes across rural and urban settings at different spatial reporting units. Only outbreaks reported at the first-, second- and third-level administrative units are included.

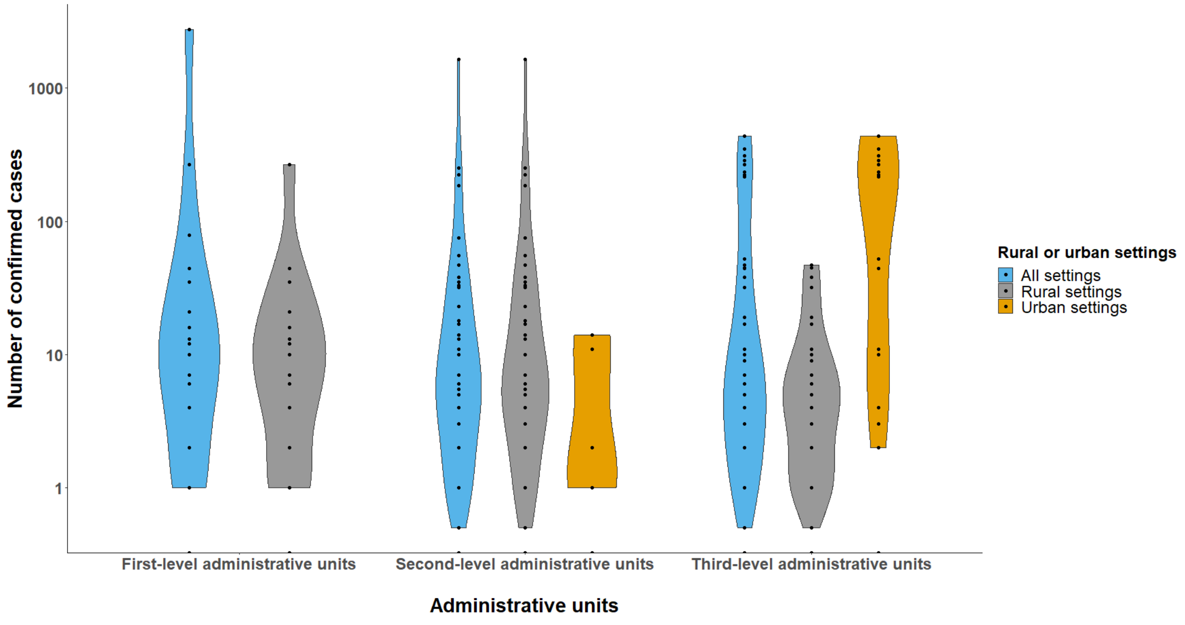


**Figure S18. Distributions of the number of confirmed cases stratified by rural and urban settings**

This figure shows the distributions of the number of confirmed cases across rural and urban settings at different spatial reporting units. Only outbreaks reported at the first-, second- and third-level administrative units are included.

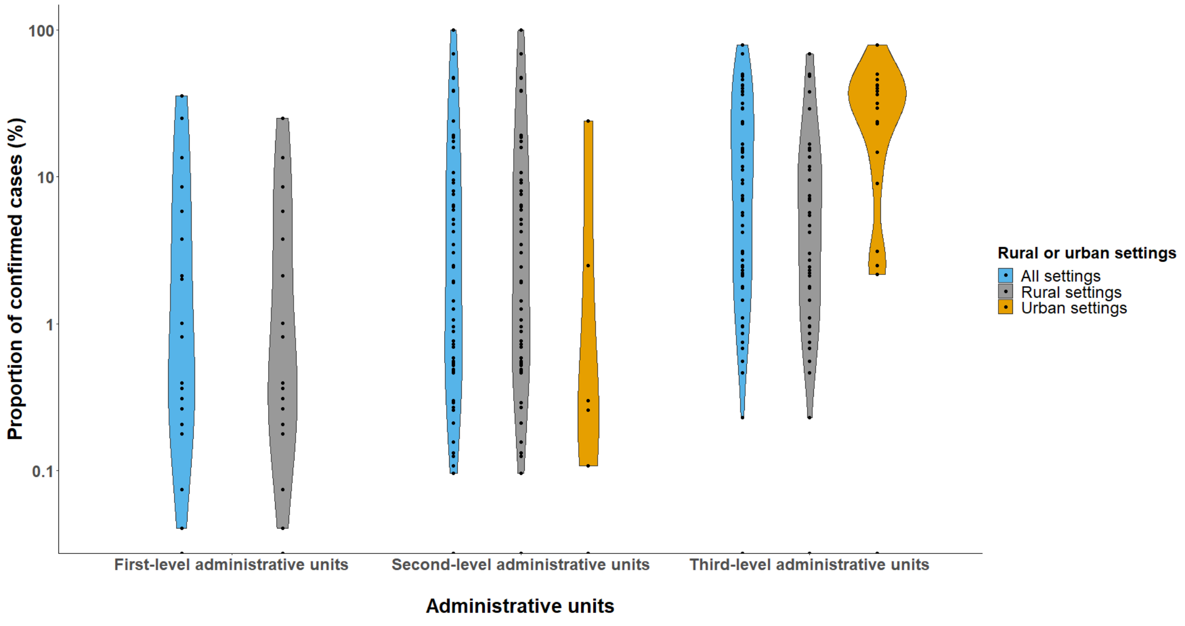


**Figure S19. Distributions of the proportion of confirmed cases stratified by rural and urban settings**

This figure shows the distributions of the proportion of confirmed cases across rural and urban settings at different spatial reporting units. Only outbreaks reported at the first-, second- and third-level administrative units are included.

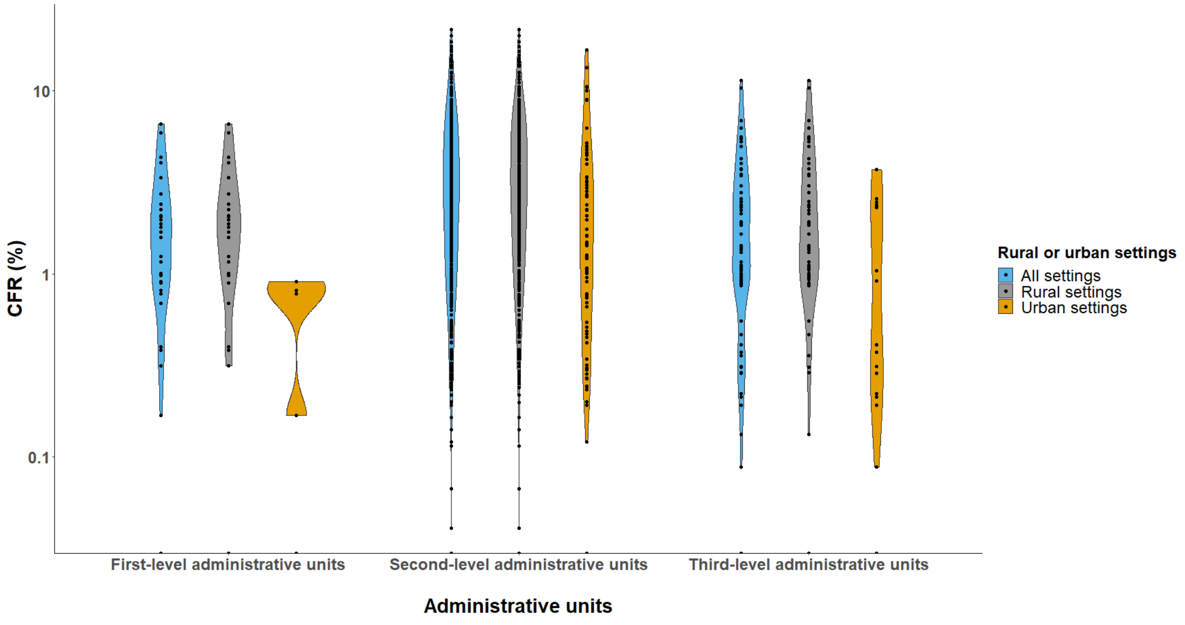


**Figure S20. Distributions of CFRs (%) stratified by rural and urban settings**

This figure shows the distributions of CFRs of outbreaks across rural and urban settings at different spatial reporting units. Only outbreaks reported at the first-, second- and third-level administrative units are included.

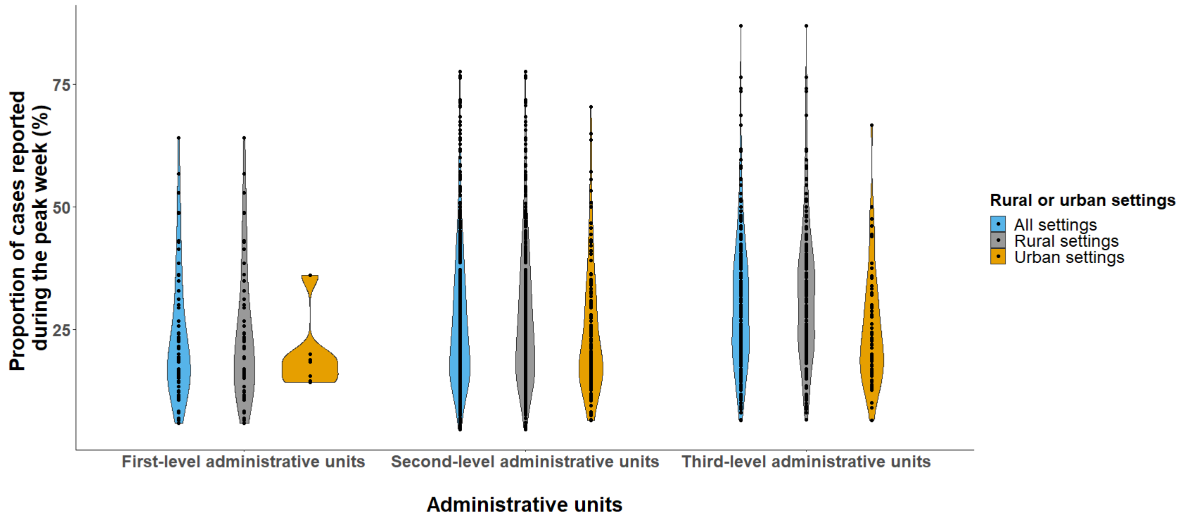


**Figure S21. Distributions of proportion of cases reported during the peak week stratified by rural and urban settings**

This figure shows the distributions of the proportion of cases reported during the peak week of outbreaks across rural and urban settings at different spatial reporting units. Only outbreaks reported at the first-, second- and third-level administrative units are included.

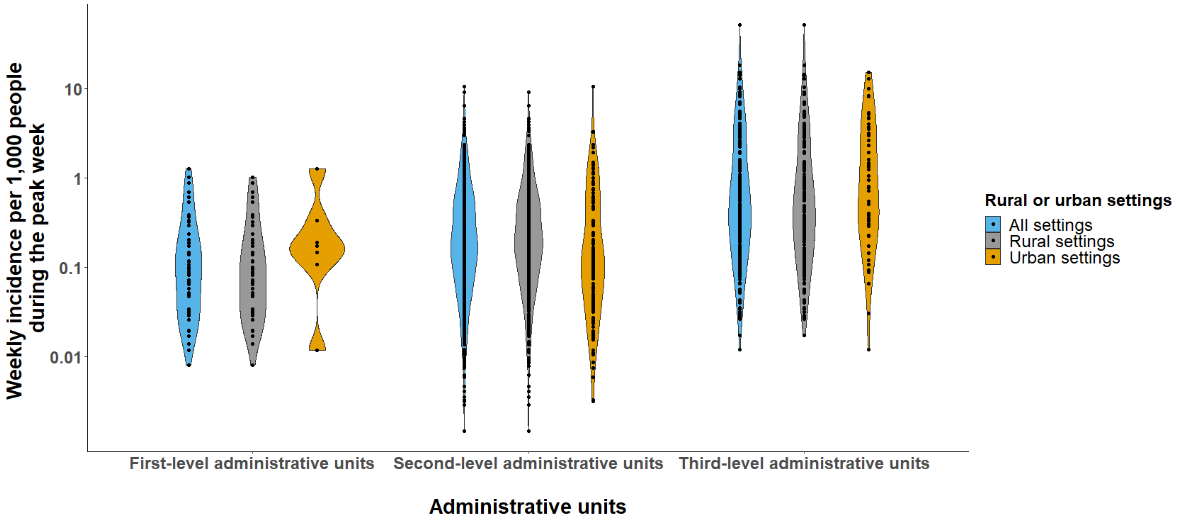


**Figure S22. Distributions of weekly incidence per 1,000 people during the peak week stratified by rural and urban settings**

This figure shows the distributions of weekly incidence per 1,000 people during the peak week of outbreaks across rural and urban settings at different spatial reporting units. Only outbreaks reported at the first-, second- and third-level administrative units are included.

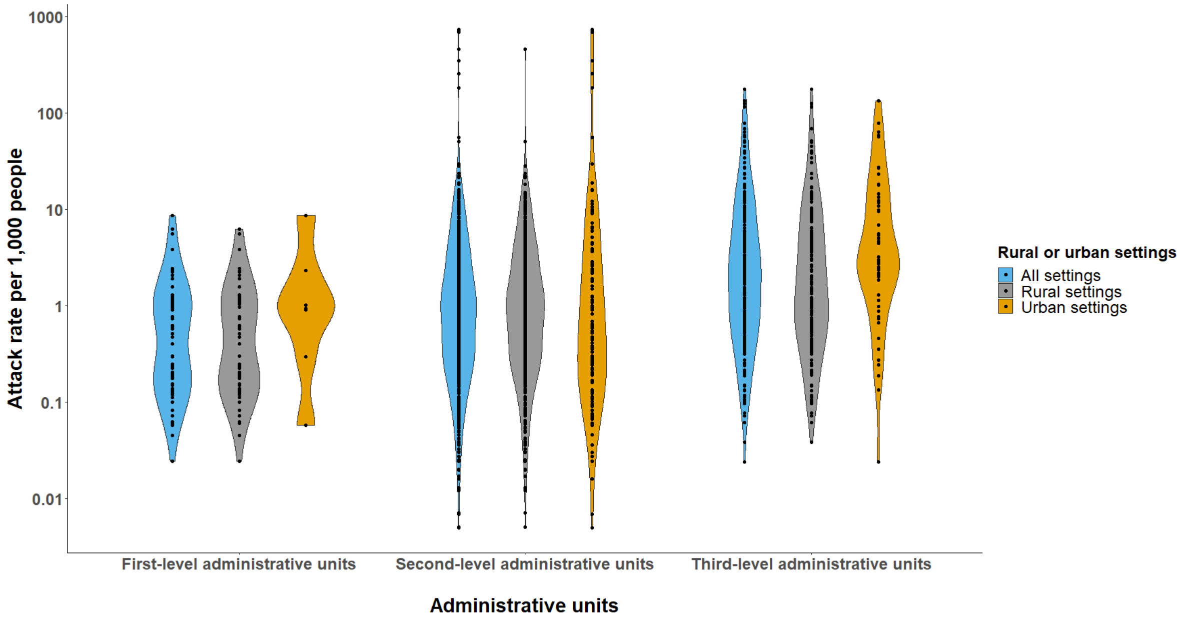


**Figure S23. Distributions of attack rates per 1,000 people stratified by rural and urban settings**

This figure shows the distributions of attack rates per 1,000 people across rural and urban settings at different spatial reporting units. Only outbreaks reported at the first-, second- and third-level administrative units are included.

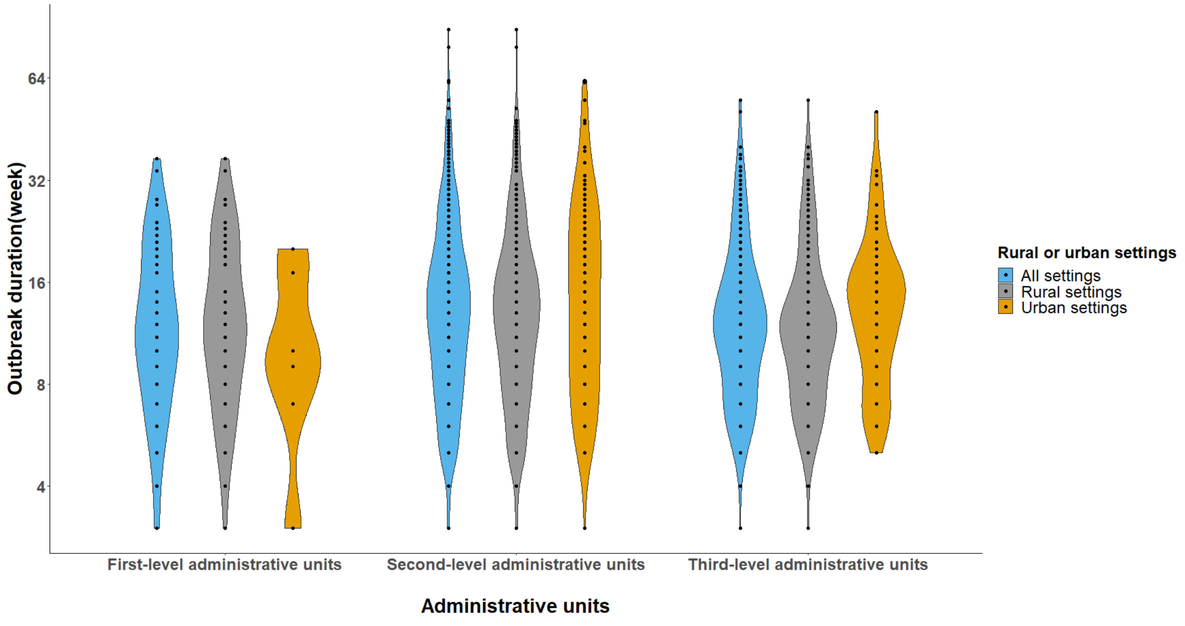


**Figure S24. Distributions of outbreak durations (weeks) stratified by rural and urban settings**

This figure shows the distributions of duration of outbreaks across rural and urban settings at different spatial reporting units. Only outbreaks reported at the first-, second- and third-level administrative units are included.

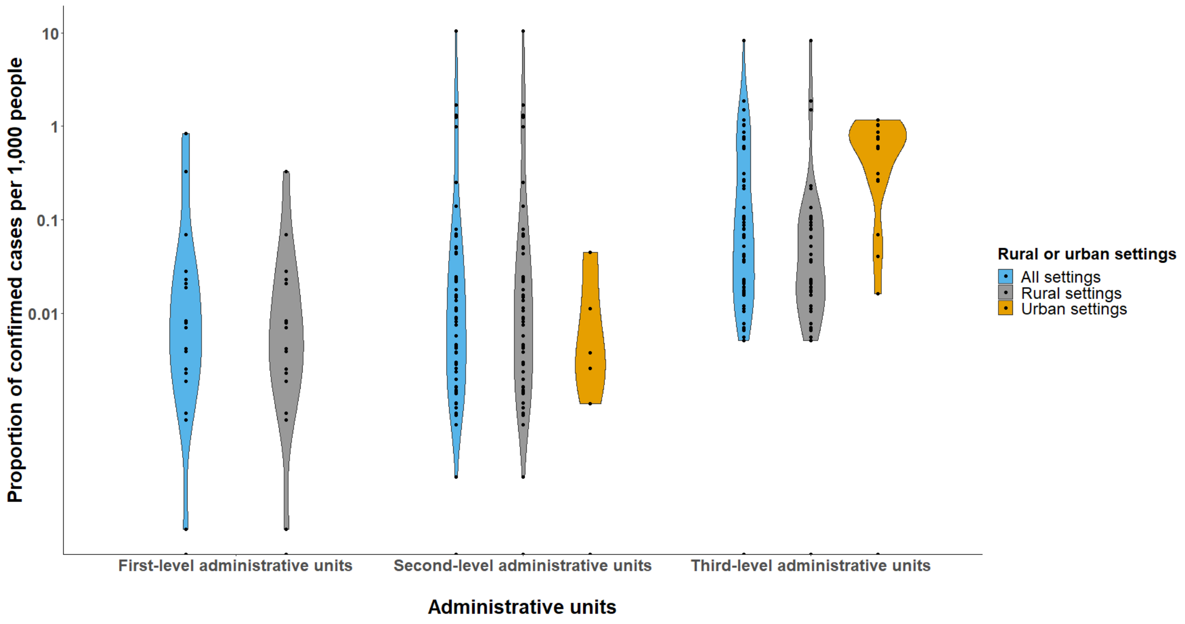


**Figure S25. Distributions of the proportion of confirmed cases per 1,000 people stratified by rural and urban settings**

This figure shows the distributions of the proportion of confirmed cases per 1,000 people across rural and urban settings at different spatial reporting units. Only outbreaks reported at the first-, second- and third-level administrative units are included.

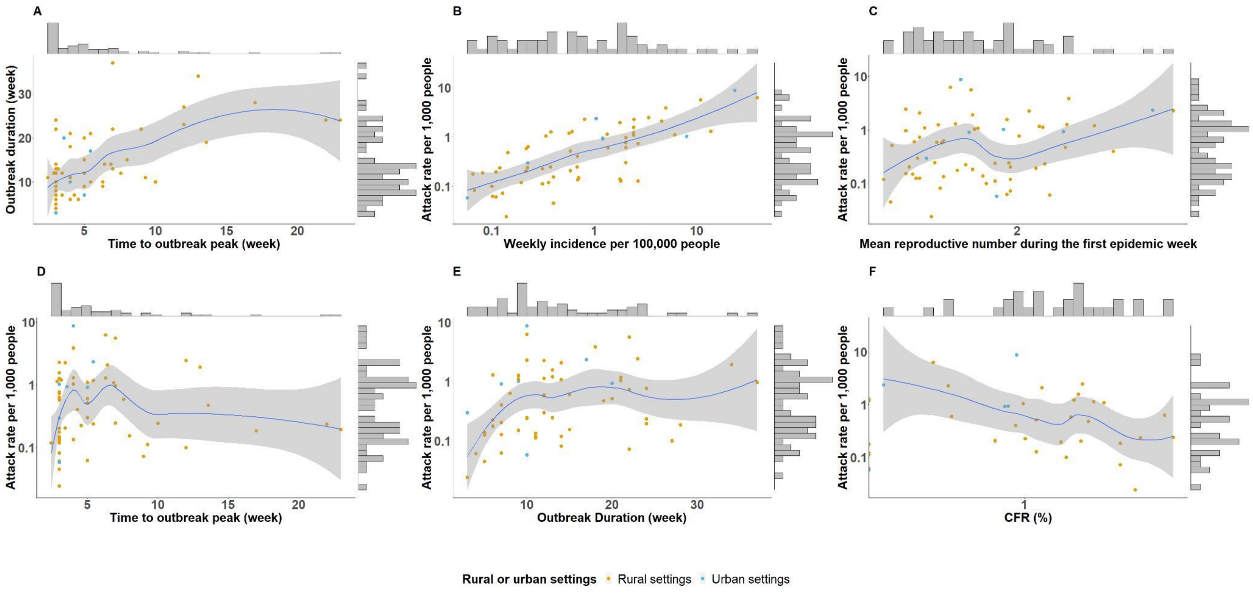

**Figure S26. Bivariate relationships between epidemic metrics among outbreaks reported at the first-level administrative units**

This figure shows the correlations between different epidemic metrics for first-level administrative unit outbreaks, including outbreak threshold, mean reproductive number during the first epidemic week, attack rate, duration, time to outbreak peak and CFR. The marginal histograms show the distributions of univariates. Panel A shows the correlation between time to outbreak peak (week) and outbreak duration (week). Panel B shows the correlation between outbreak threshold (i.e., weekly incidence per 100,000 people) and attack rate per 1,000 people. Panel C shows the correlation between mean reproductive number during the first epidemic week and attack rate per 1,000 people. Panel D shows the correlation between time to outbreak peak (week) and attack rate per 1,000 people. Panel E shows the correlation between outbreak duration (week) and attack rate per 1,000 people. Panel F shows the correlation between CFR (%) and attack rate per 1,000 people.

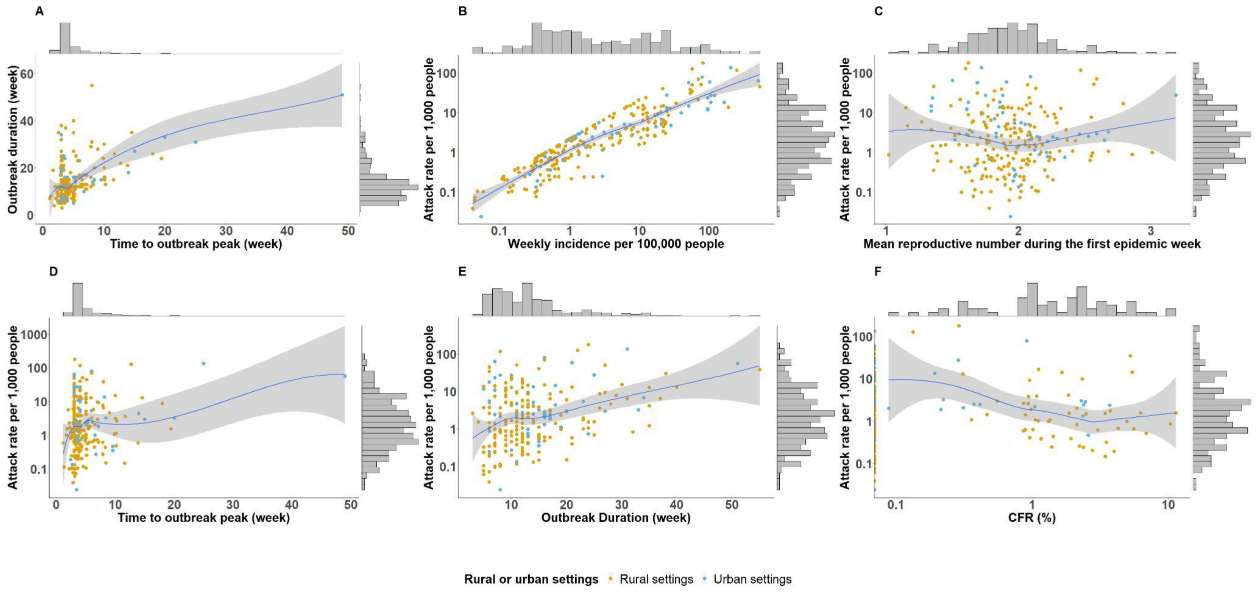


**Figure S27. Bivariate relationships between epidemic metrics among outbreaks reported at the third-level administrative units**

This figure shows the correlations between different epidemic metrics for third-level administrative unit outbreaks, including outbreak threshold, mean reproductive number during the first epidemic week, attack rate, duration, time to outbreak peak and CFR. The marginal histograms show the distributions of univariates. Panel A shows the correlation between time to outbreak peak (week) and outbreak duration (week). Panel B shows the correlation between outbreak threshold (i.e., weekly incidence per 100,000 people) and attack rate per 1,000 people. Panel C shows the correlation between mean reproductive number during the first epidemic week and attack rate per 1,000 people. Panel D shows the correlation between time to outbreak peak (week) and attack rate per 1,000 people. Panel E shows the correlation between outbreak duration (week) and attack rate per 1,000 people. Panel F shows the correlation between CFR (%) and attack rate per 1,000 people.

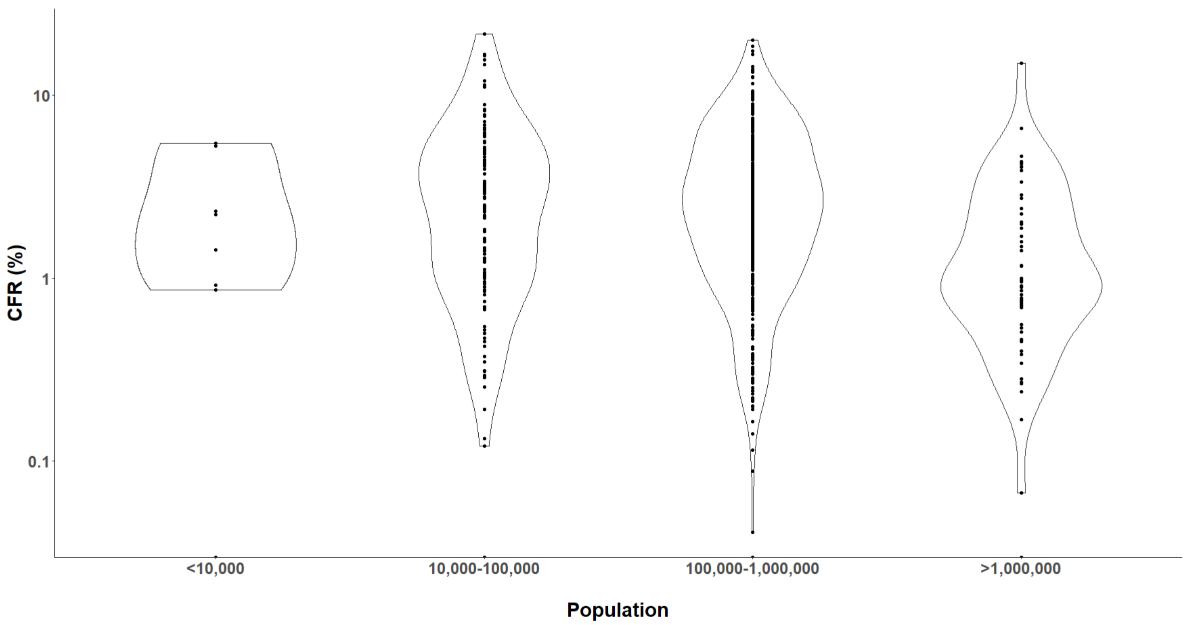


**Figure S28. Distributions of CFRs (%) stratified by population sizes**
This figure shows the distributions of CFRs (%) in regions with different population sizes.

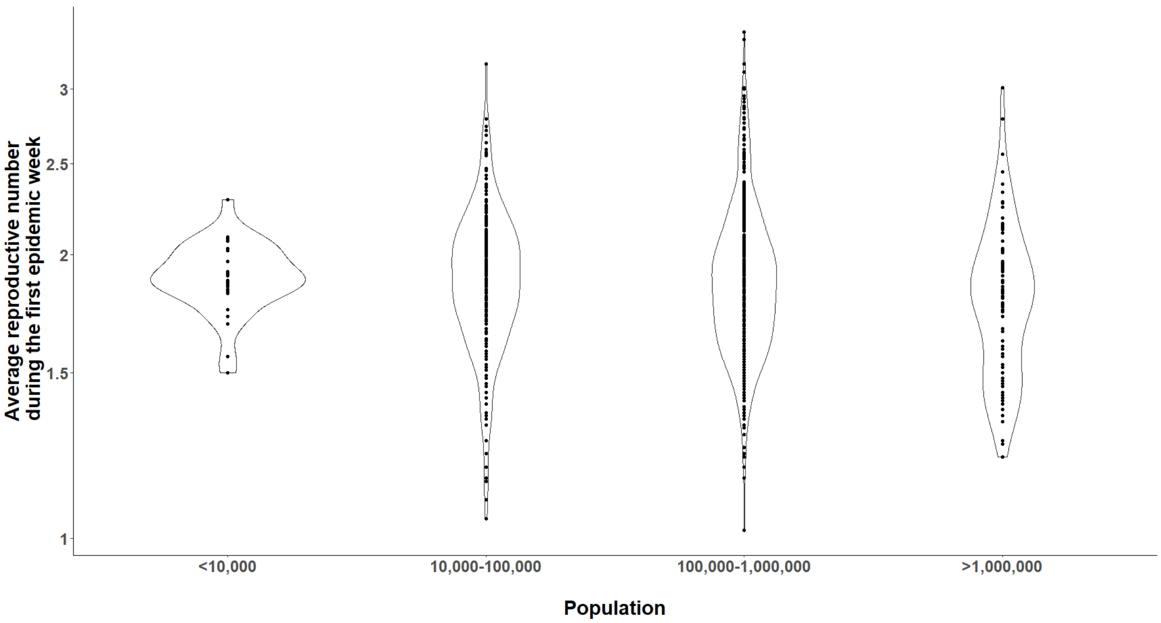


**Figure S29. Distributions of early reproductive numbers during the first epidemic week stratified by population sizes**

This figure shows the distributions of early reproductive numbers during the first epidemic week in regions with different population sizes.

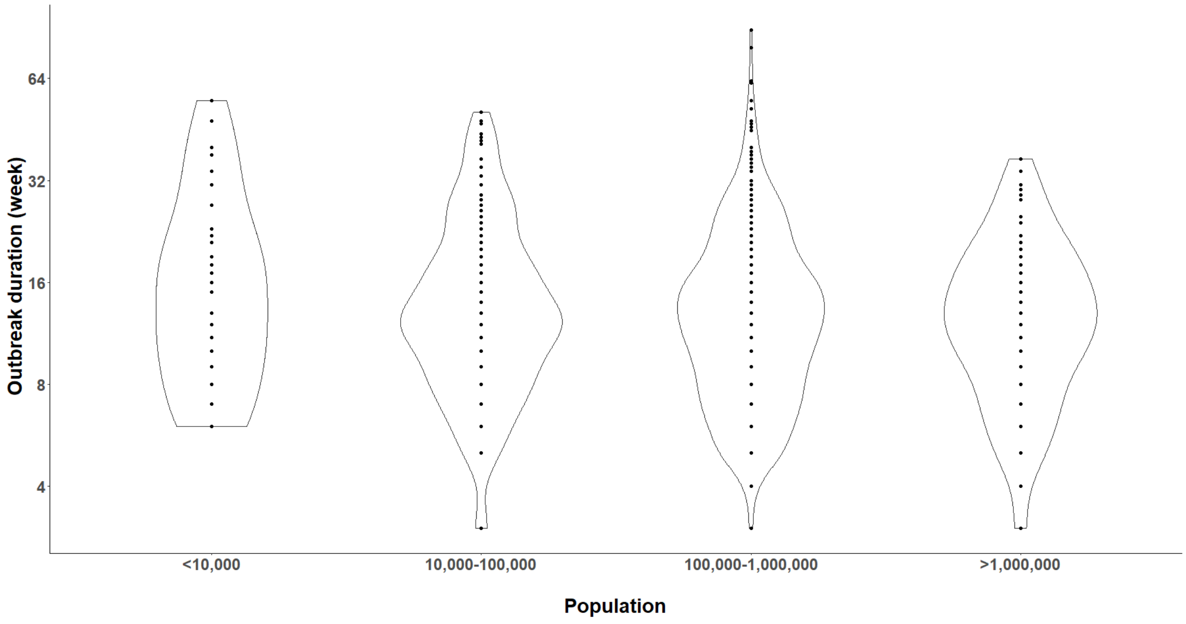


**Figure S30. Distributions of outbreak duration (week) stratified by population sizes**

This figure shows the distributions of outbreak duration (week) in regions with different population sizes.

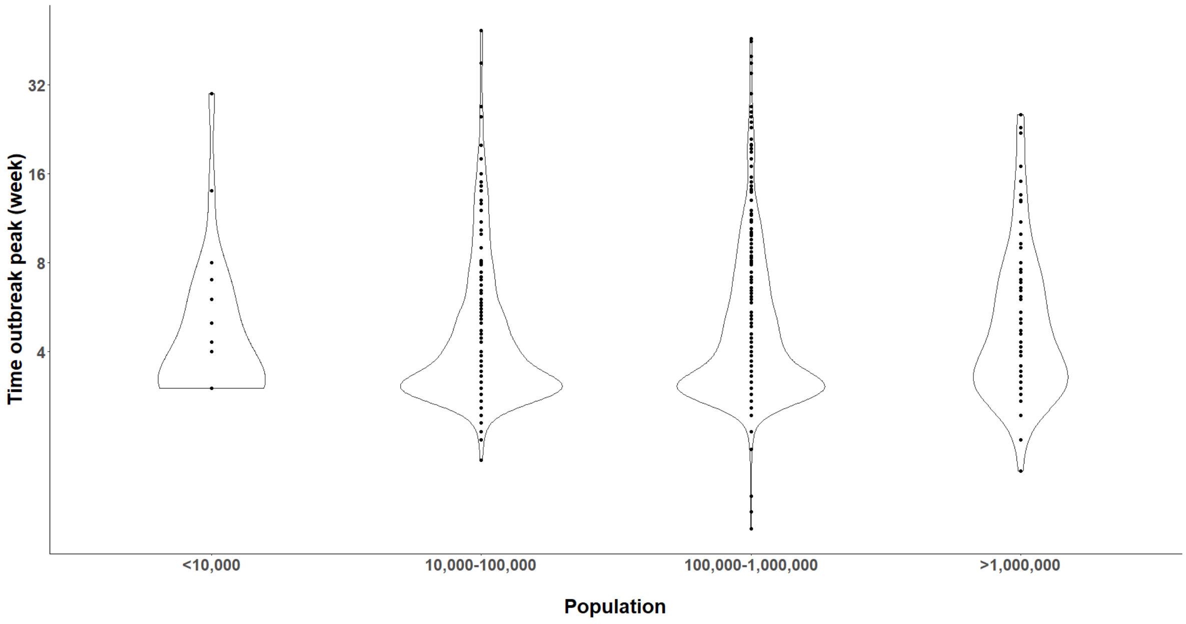

**Figure S31. Distributions of time to outbreak peak (week) stratified by population sizes**
This figure shows the distributions of time to outbreak peak (week) in regions with different population sizes.

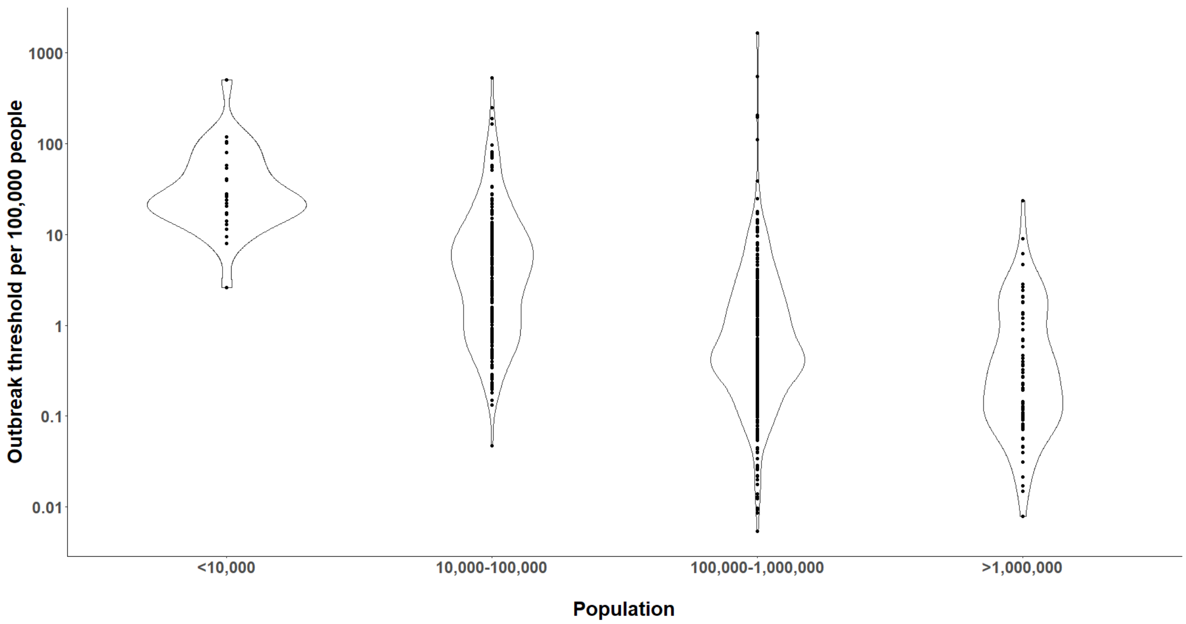


**Figure S32. Distributions of outbreak thresholds stratified by population sizes**
This figure shows the distributions of outbreak thresholds in regions with different population sizes.

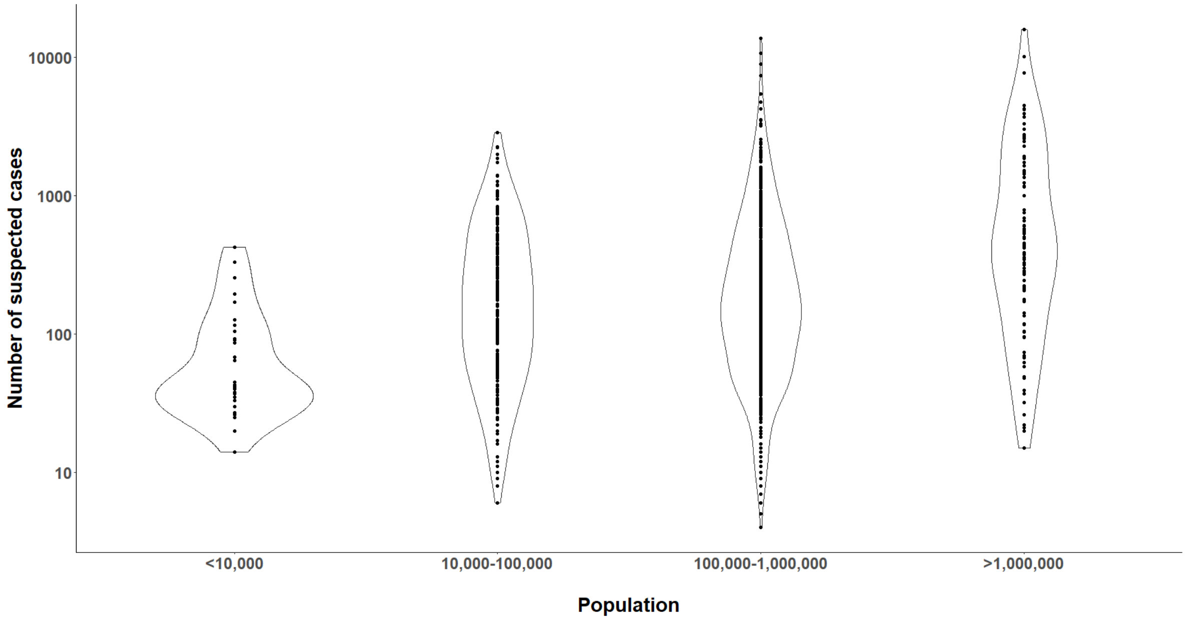


**Figure S33. Distributions of number of suspected cases stratified by population sizes**
This figure shows the distributions of number of suspected cases in regions with different population sizes.

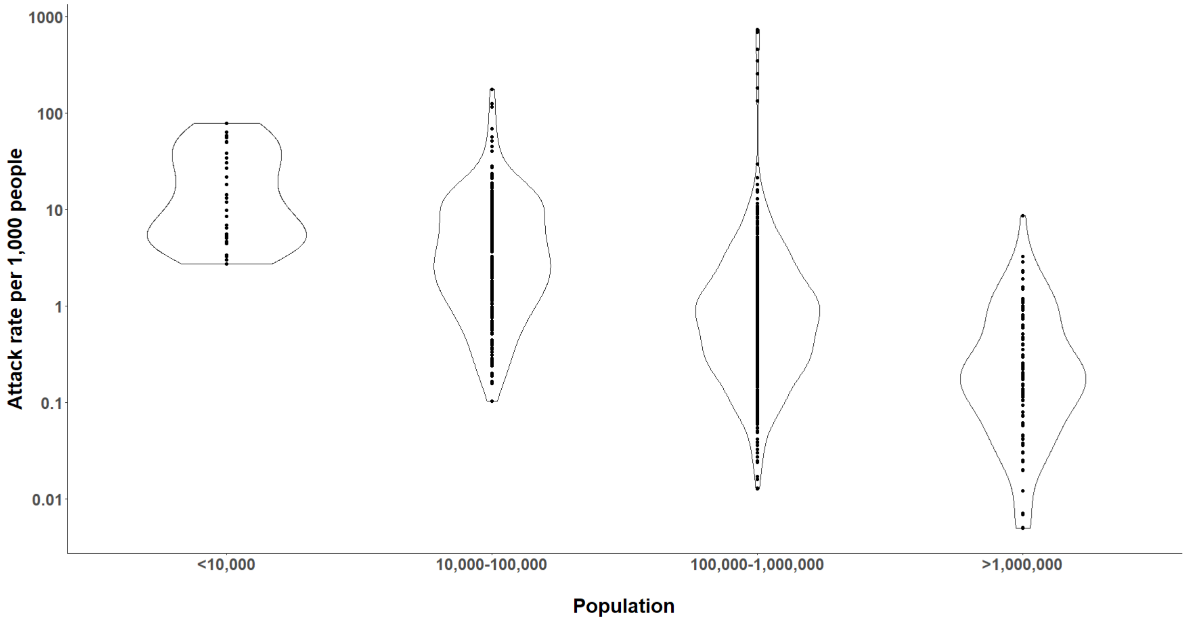


**Figure S34. Distributions of attack rates per 1,000 stratified by population sizes**
This figure shows the distributions of attack rates per 1,000 in regions with different population sizes.

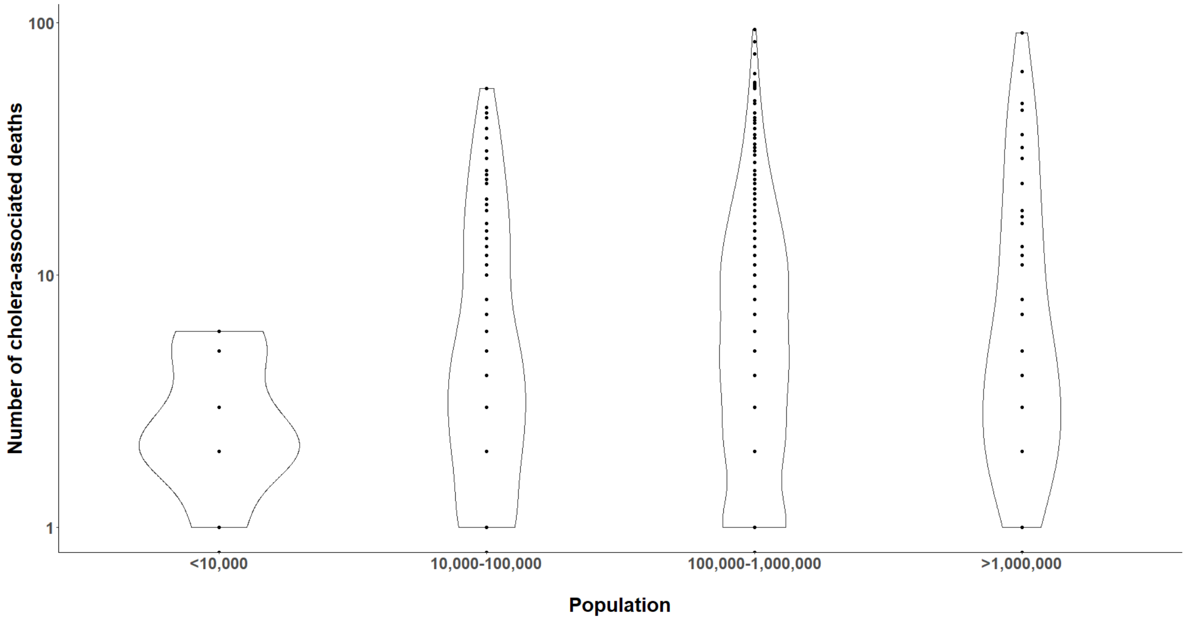


**Figure S35. Distributions of number of cholera-associated deaths stratified by population sizes**

This figure shows the distributions of number of cholera-associated deaths in regions with different population sizes.

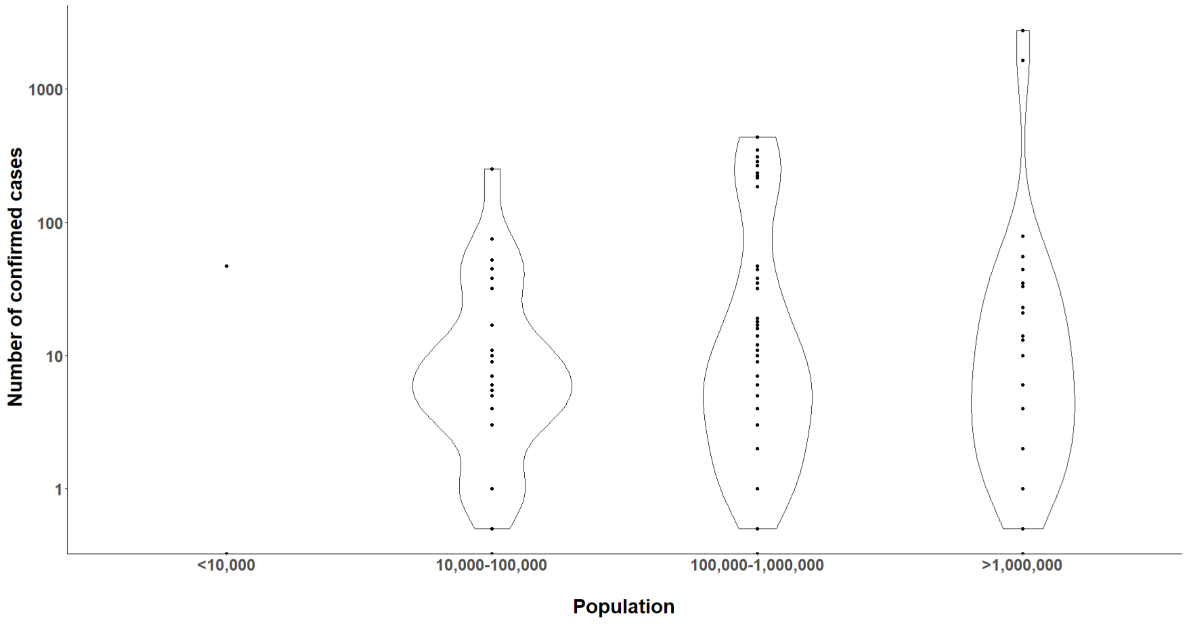


**Figure S36. Distributions of number of confirmed cases stratified by population sizes**

This figure shows the distributions of number of confirmed cases in regions with different population sizes.

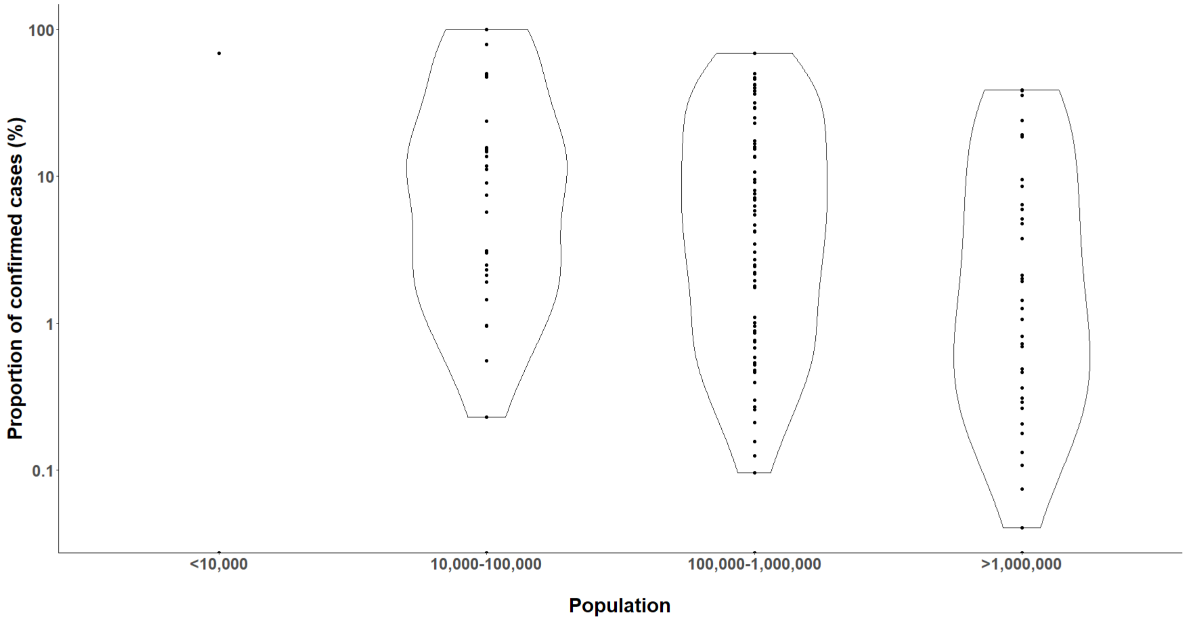


**Figure S37. Distributions of proportion of confirmed cases stratified by population sizes**

This figure shows the distributions of proportion of confirmed cases in regions with different population sizes.

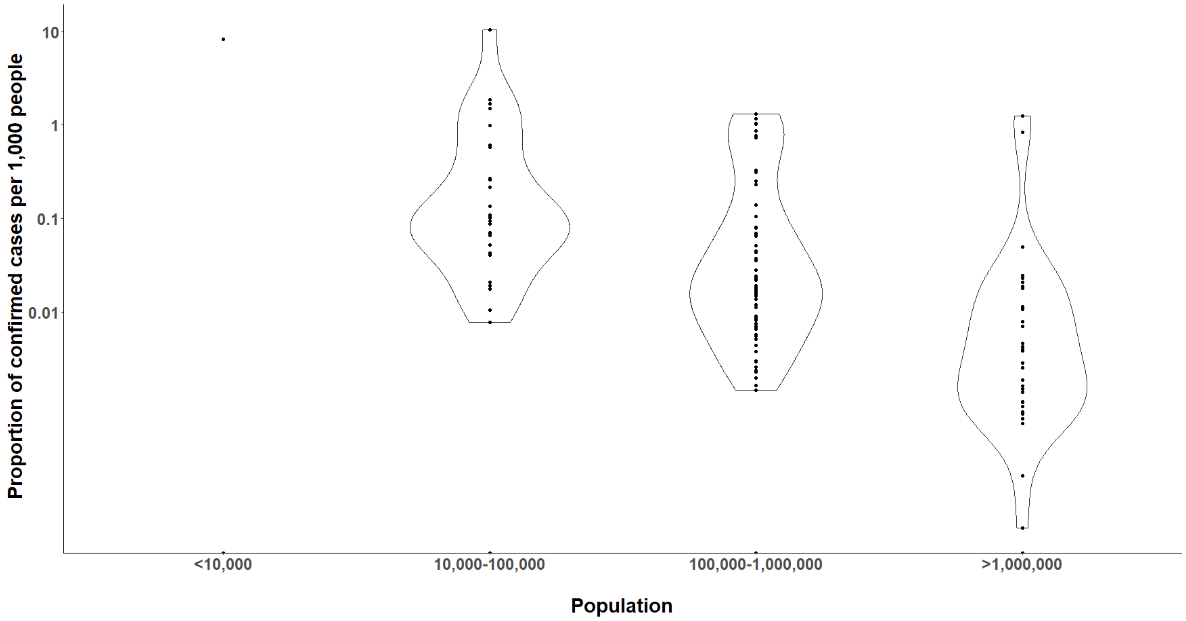


**Figure S38. Distributions of proportion of confirmed cases per 1,000 people stratified by population sizes**

This figure shows the distributions of proportion of confirmed cases per 1,000 people in regions with different population sizes.

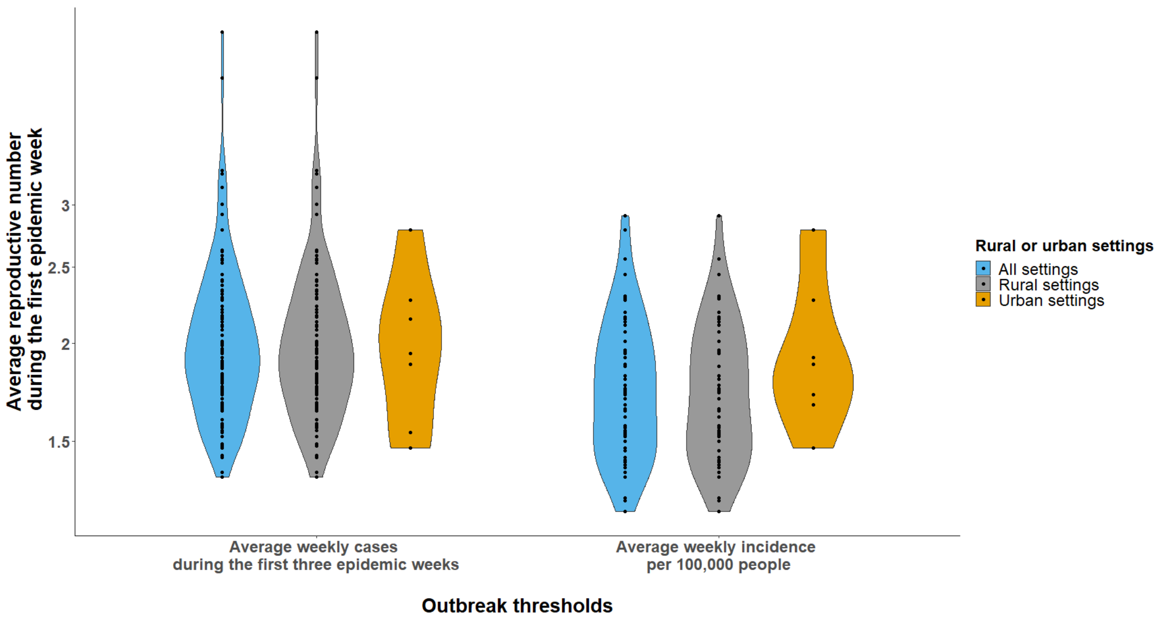


**Figure S39. Distributions of average reproductive numbers during the first epidemic week stratified by rural and urban settings using different outbreak threshold definitions among outbreaks at the first-level administrative units**
This figure shows the distributions of average reproductive numbers during the first epidemic week across rural and urban settings using different outbreak threshold definitions. Only outbreaks reported at the first-level administrative units are included.

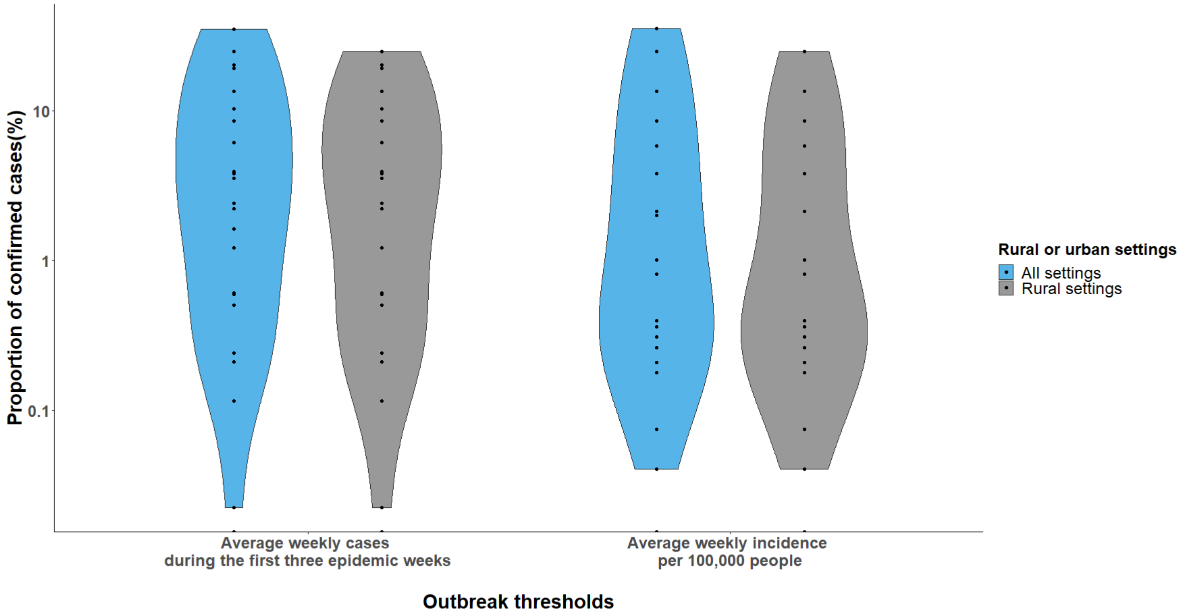


**Figure S40. Distributions of proportion of confirmed cases (%) stratified by rural and urban settings using different outbreak threshold definitions among outbreaks at the first-level administrative units**

This figure shows the distributions of proportion of confirmed cases (%) across rural and urban settings using different outbreak threshold definitions. Only outbreaks reported at the first-level administrative units are included.

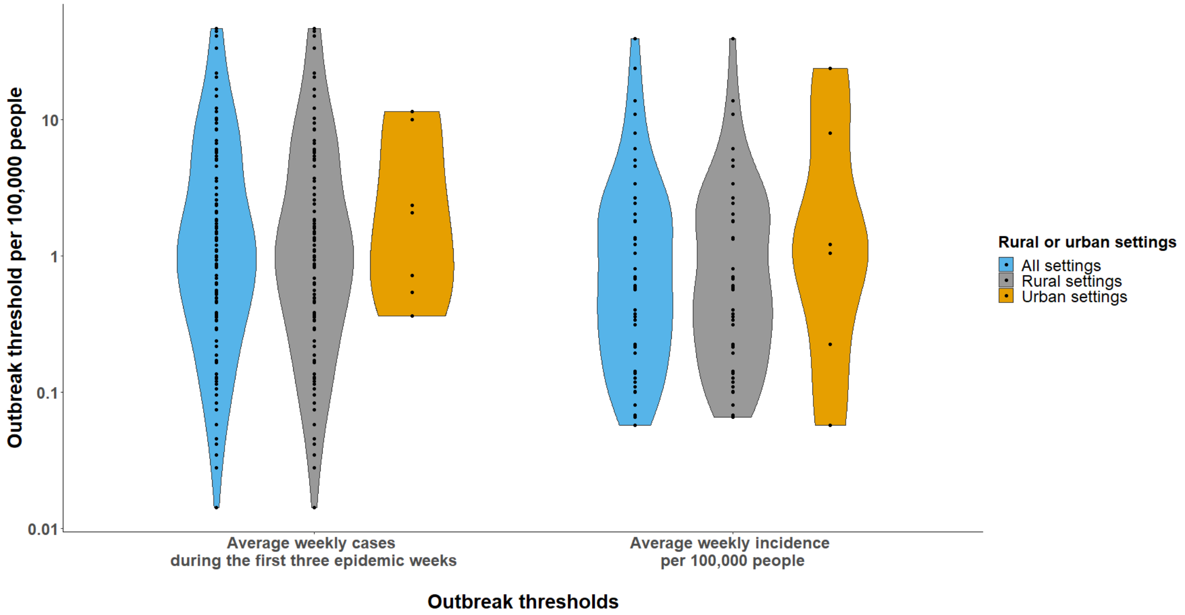


**Figure S41. Distributions of outbreak thresholds per 100,000 people stratified by rural and urban settings using different outbreak threshold definitions among outbreaks at the first-level administrative units**

This figure shows the distributions of outbreak thresholds per 100,000 people across rural and urban settings using different outbreak threshold definitions. Only outbreaks reported at the first-level administrative units are included.

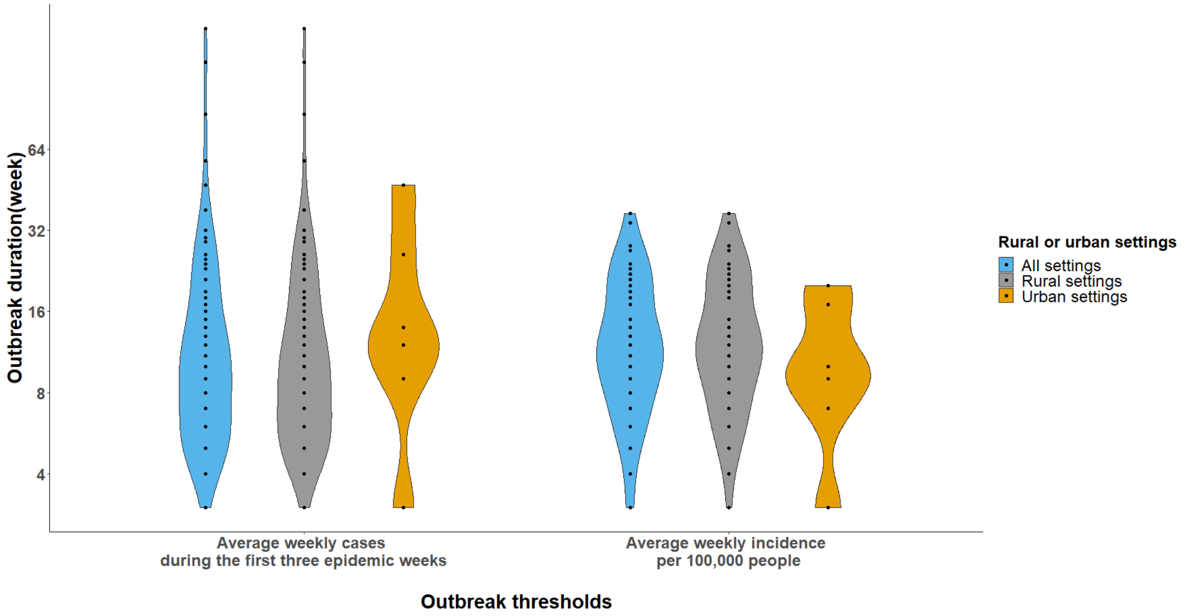


**Figure S42. Distributions of outbreak durations (week) stratified by rural and urban settings using different outbreak threshold definitions among outbreaks at the first-level administrative units**

This figure shows the distributions of outbreak durations (week) across rural and urban settings using different outbreak threshold definitions. Only outbreaks reported at the first-level administrative units are included.

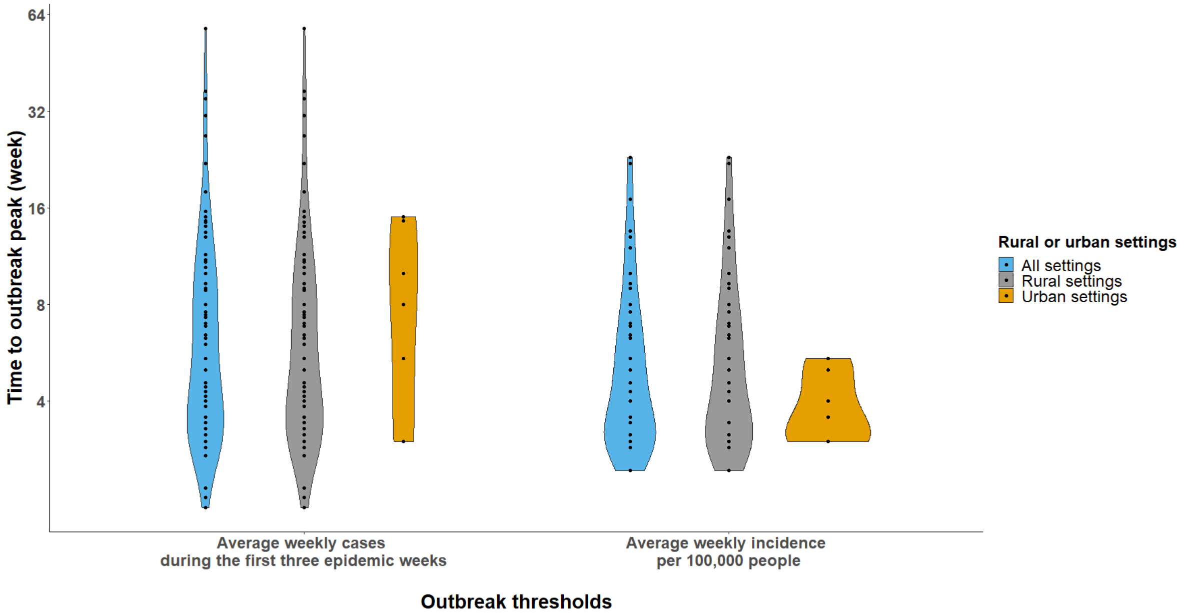


**Figure S43. Distributions of time to outbreak peak (week) stratified by rural and urban settings using different outbreak threshold definitions among outbreaks at the first-level administrative units**

This figure shows the distributions of time to outbreak peak (week) across rural and urban settings using different outbreak threshold definitions. Only outbreaks reported at the first-level administrative units are included.

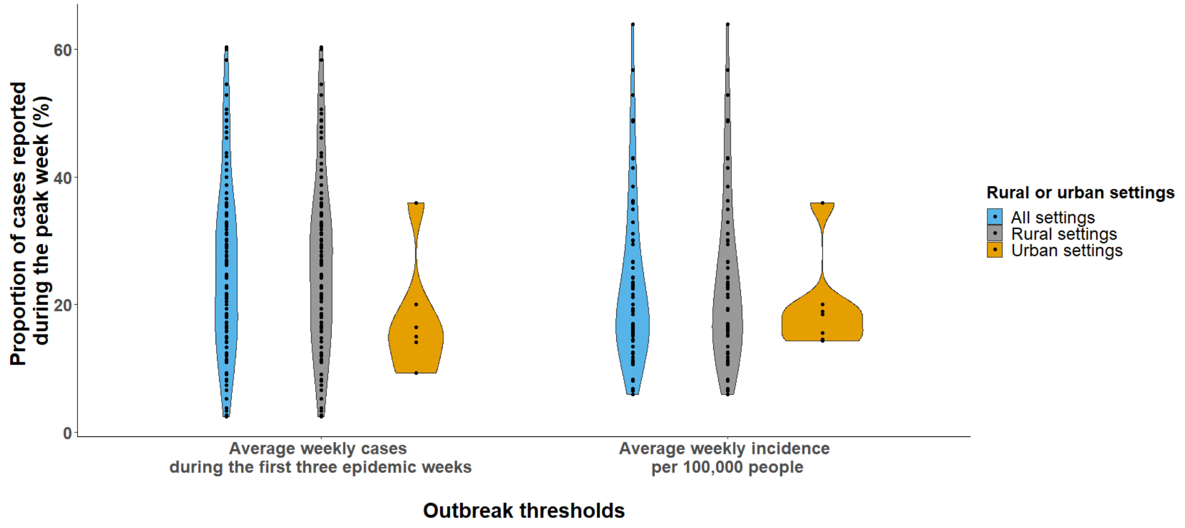


**Figure S44. Distributions of the proportion of cases reported during the peak week (%) stratified by rural and urban settings using different outbreak threshold definitions among outbreaks at the first-level administrative units**

This figure shows the distributions of the proportion of cases reported during the peak week (%) across rural and urban settings using different outbreak threshold definitions. Only outbreaks reported at the first-level administrative units are included.

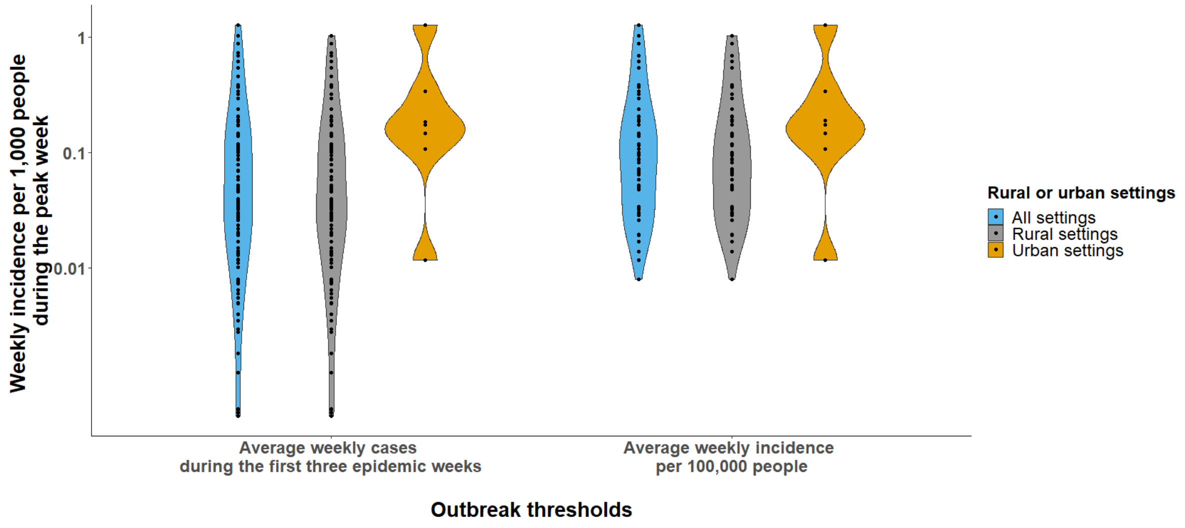


**Figure S45. Distributions of the weekly incidence per 1,000 people during the peak week stratified by rural and urban settings using different outbreak threshold definitions among outbreaks at the first-level administrative units**

This figure shows the distributions of the weekly incidence per 1,000 people during the peak week across rural and urban settings using different outbreak threshold definitions. Only outbreaks reported at the first-level administrative units are included.

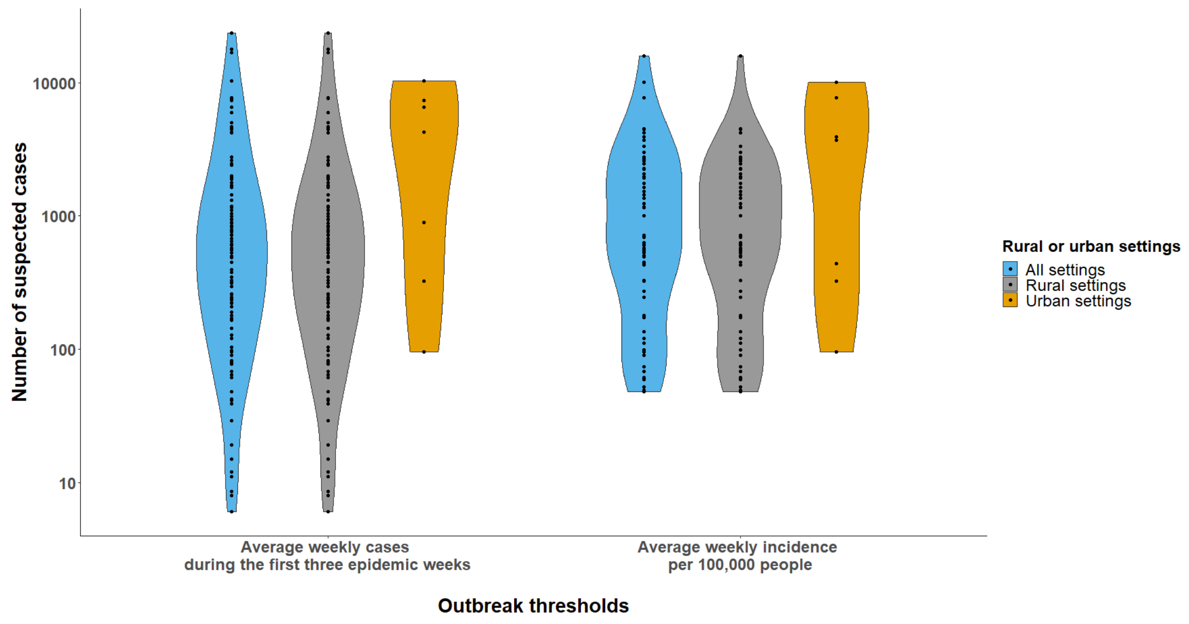


**Figure S46. Distributions of number of suspected cases stratified by rural and urban settings using different outbreak threshold definitions among outbreaks at the first-level administrative units**

This figure shows the distributions of number of suspected cases across rural and urban settings using different outbreak threshold definitions. Only outbreaks reported at the first-level administrative units are included.

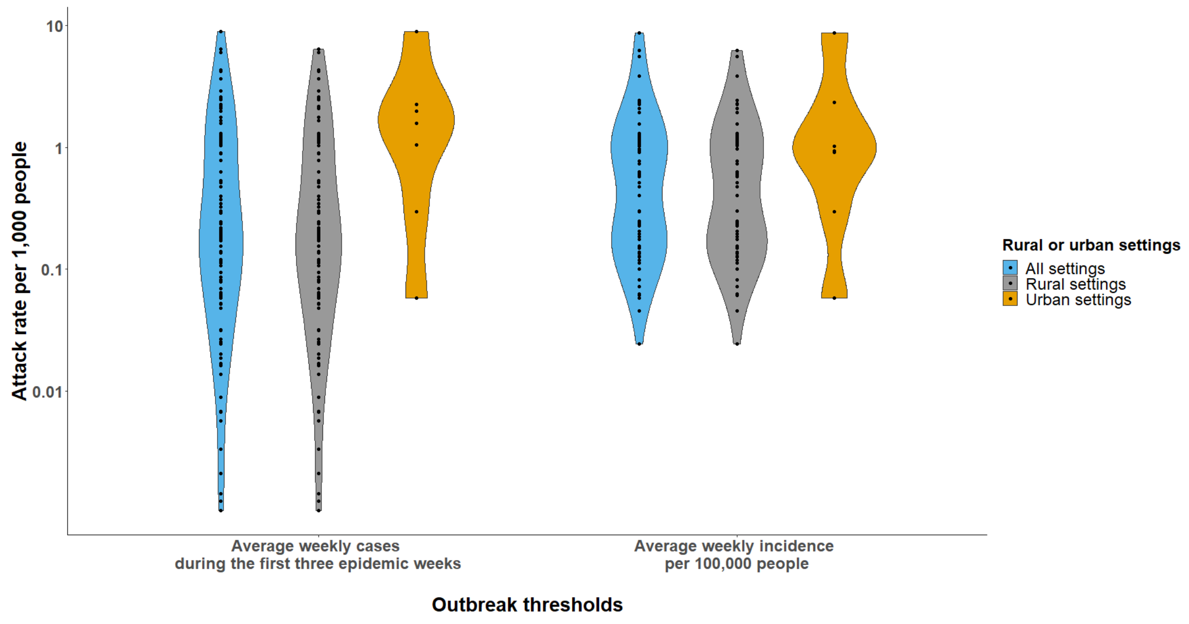

**Figure S47. Distributions of attack rates stratified by rural and urban settings using different outbreak threshold definitions among outbreaks at the first-level administrative units**

This figure shows the distributions of attack rates per 1,000 people across rural and urban settings using different outbreak threshold definitions. Only outbreaks reported at the first-level administrative units are included.

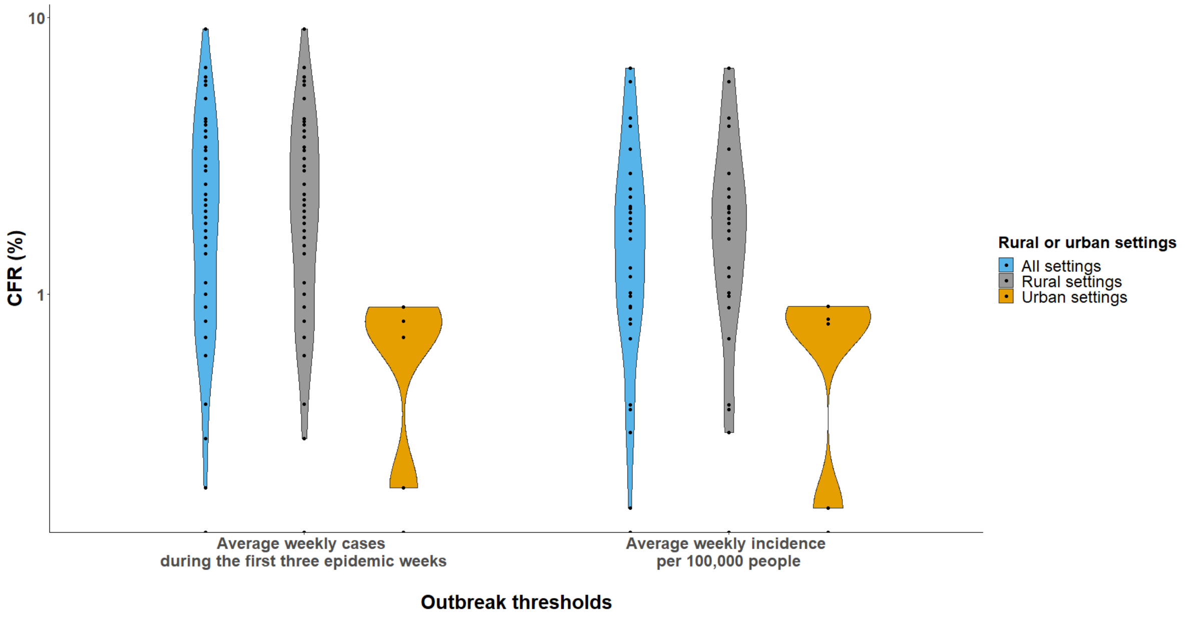


**Figure S48. Distributions of CFRs (%) stratified by rural and urban settings using different outbreak threshold definitions among outbreaks at the first-level administrative units**

This figure shows the distributions of CFR (%) across rural and urban settings using different outbreak threshold definitions. Only outbreaks reported at the first-level administrative units are included.

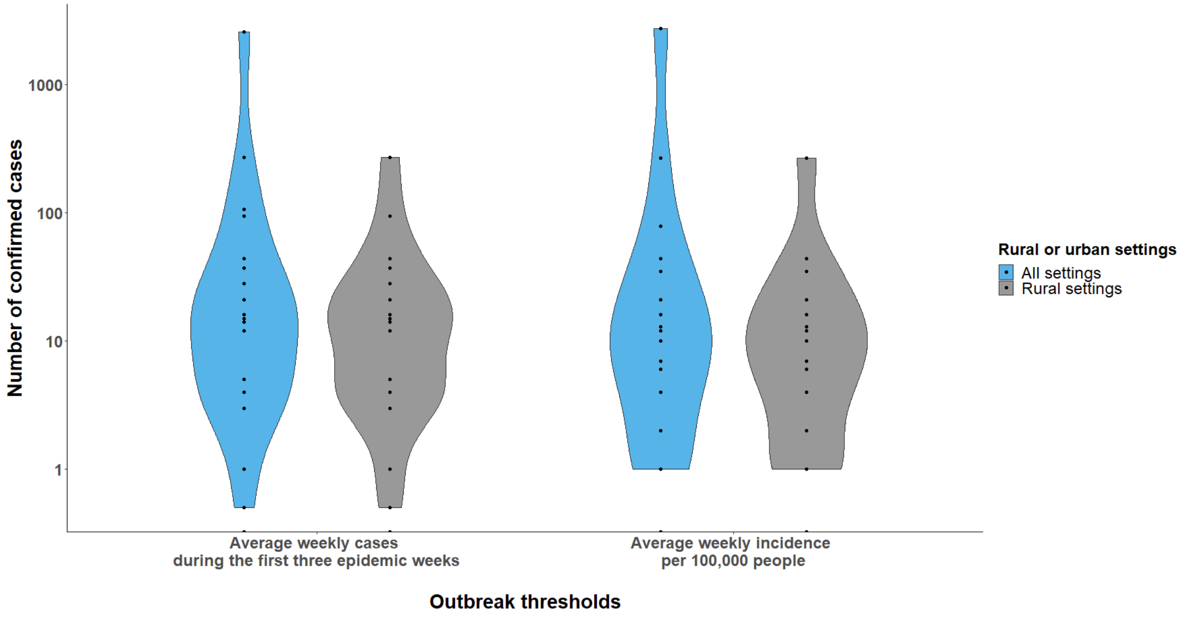


**Figure S49. Distributions of number of confirmed cases stratified by rural and urban settings using different outbreak threshold definitions among outbreaks at the first-level administrative units**

This figure shows the distributions of number of confirmed cases across rural and urban settings using different outbreak threshold definitions. Only outbreaks reported at the first-level administrative units are included.

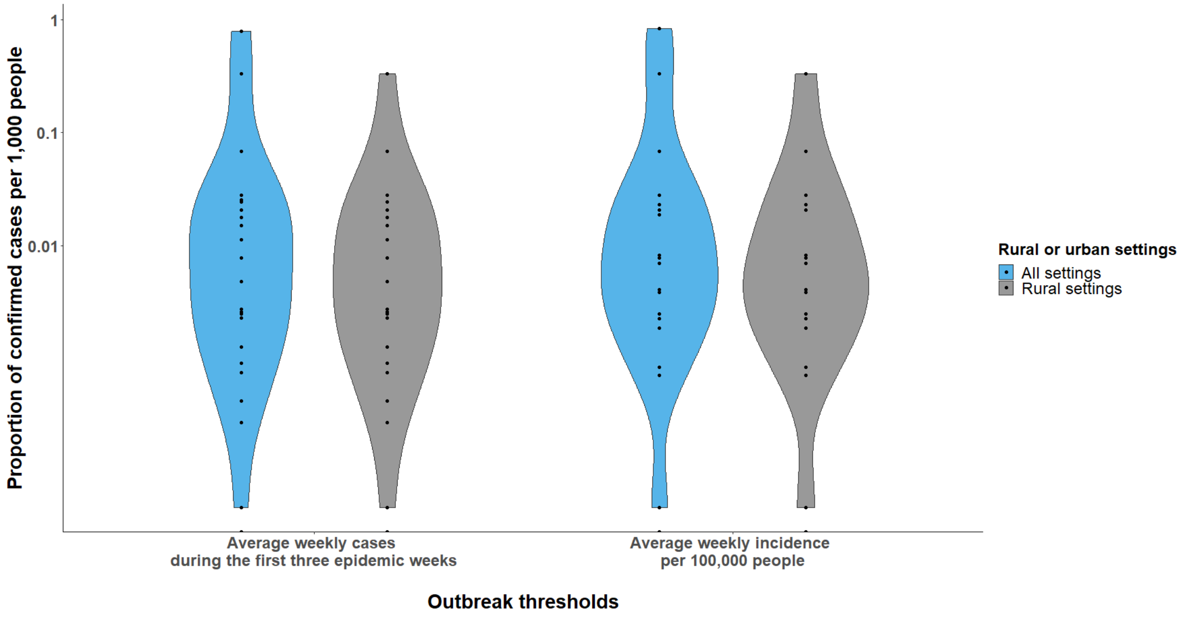

**Figure S50. Distributions of proportion of confirmed cases per 1,000 people stratified by rural and urban settings using different outbreak threshold definitions among outbreaks at the first-level administrative units**

This figure shows the distributions of proportion of confirmed cases per 1,000 people across rural and urban settings using different outbreak threshold definitions. Only outbreaks reported at the first-level administrative units are included.

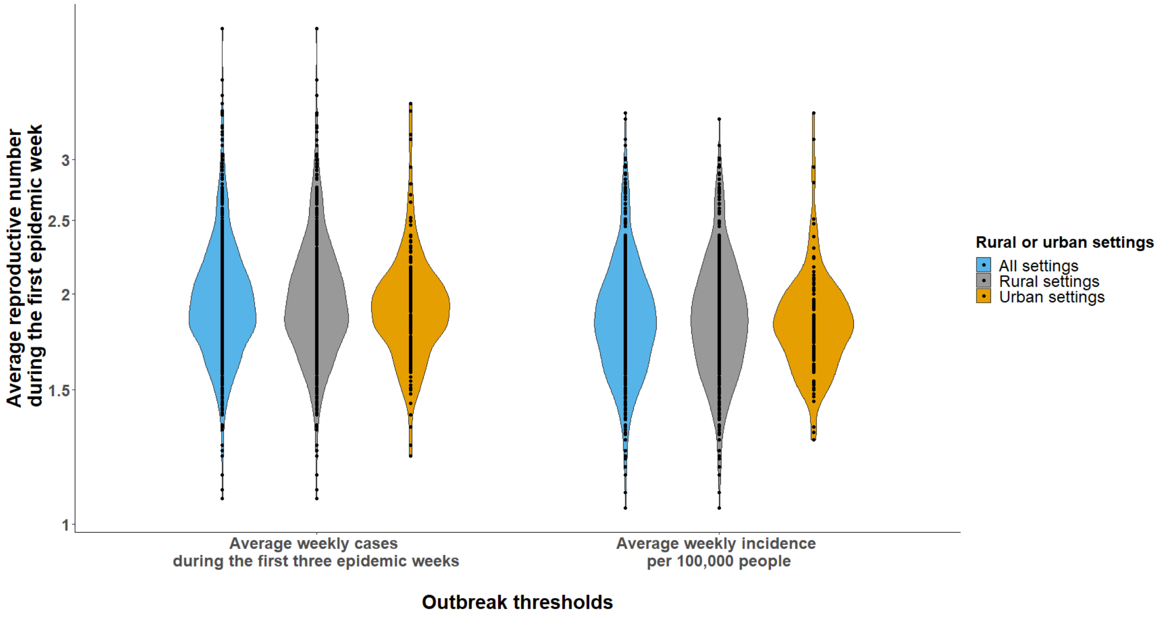


**Figure S51. Distributions of average reproductive numbers during the first epidemic week stratified by rural and urban settings using different outbreak threshold definitions among outbreaks at the second-level administrative units**

This figure shows the distributions of average reproductive numbers during the first epidemic week across rural and urban settings using different outbreak threshold definitions. Only outbreaks reported at the second-level administrative units are included.

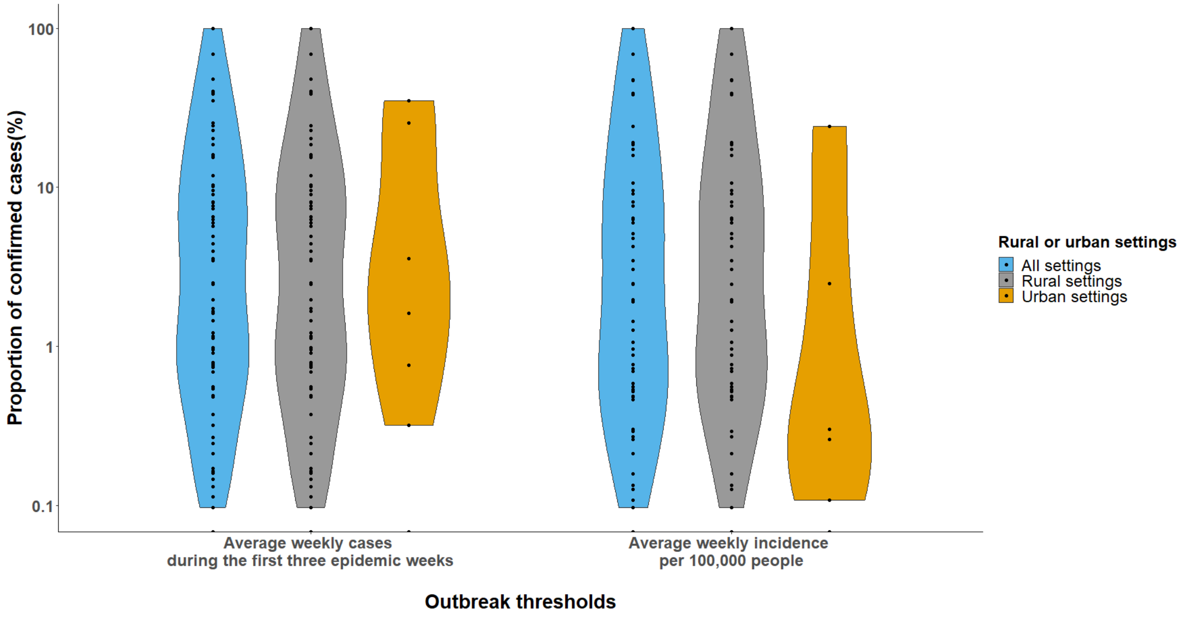


**Figure S52. Distributions of proportion of confirmed cases (%) stratified by rural and urban settings using different outbreak threshold definitions among outbreaks at the second-level administrative units**

This figure shows the distributions of proportion of confirmed cases (%) across rural and urban settings using different outbreak threshold definitions. Only outbreaks reported at the second-level administrative units are included.

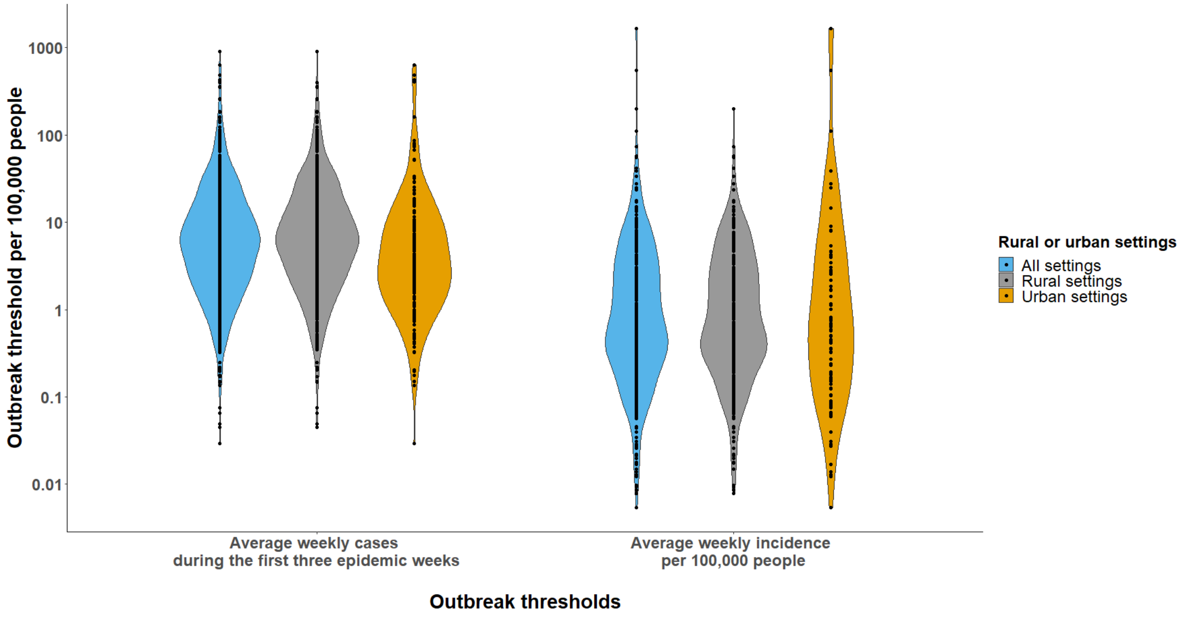

**Figure S53. Distributions of outbreak thresholds per 100,000 people stratified by rural and urban settings using different outbreak threshold definitions among outbreaks at the second-level administrative units**

This figure shows the distributions of outbreak thresholds per 100,000 people across rural and urban settings using different outbreak threshold definitions. Only outbreaks reported at the second-level administrative units are included.

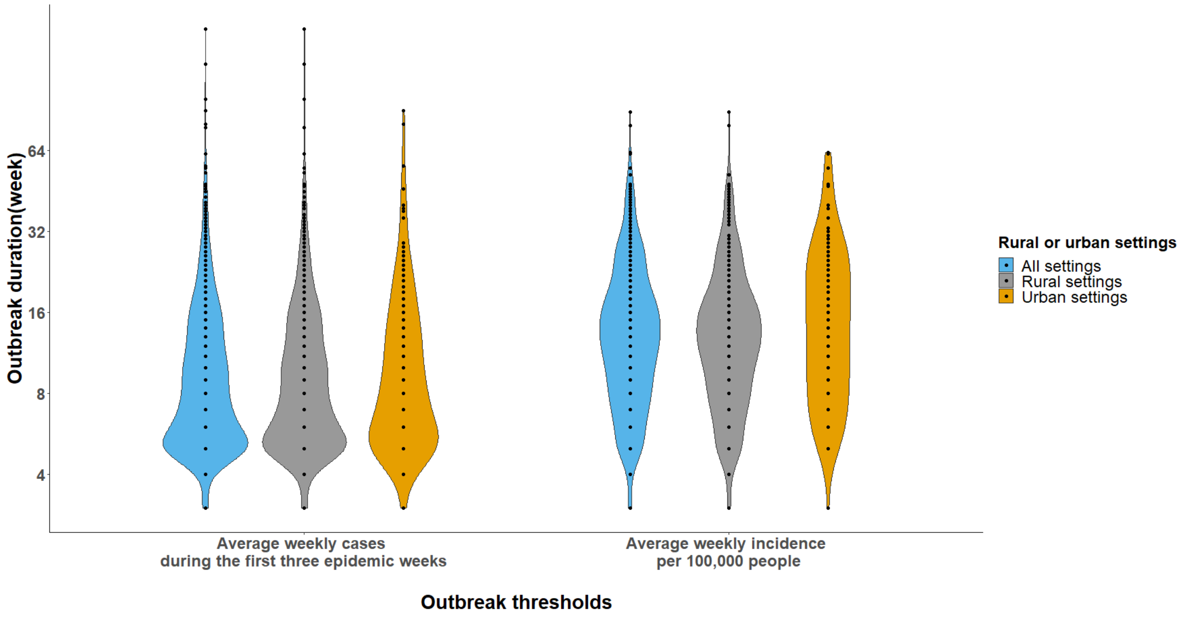


**Figure S54. Distributions of outbreak durations (week) stratified by rural and urban settings using different outbreak threshold definitions among outbreaks at the second-level administrative units**

This figure shows the distributions of outbreak durations (week) across rural and urban settings using different outbreak threshold definitions. Only outbreaks reported at the second-level administrative units are included.

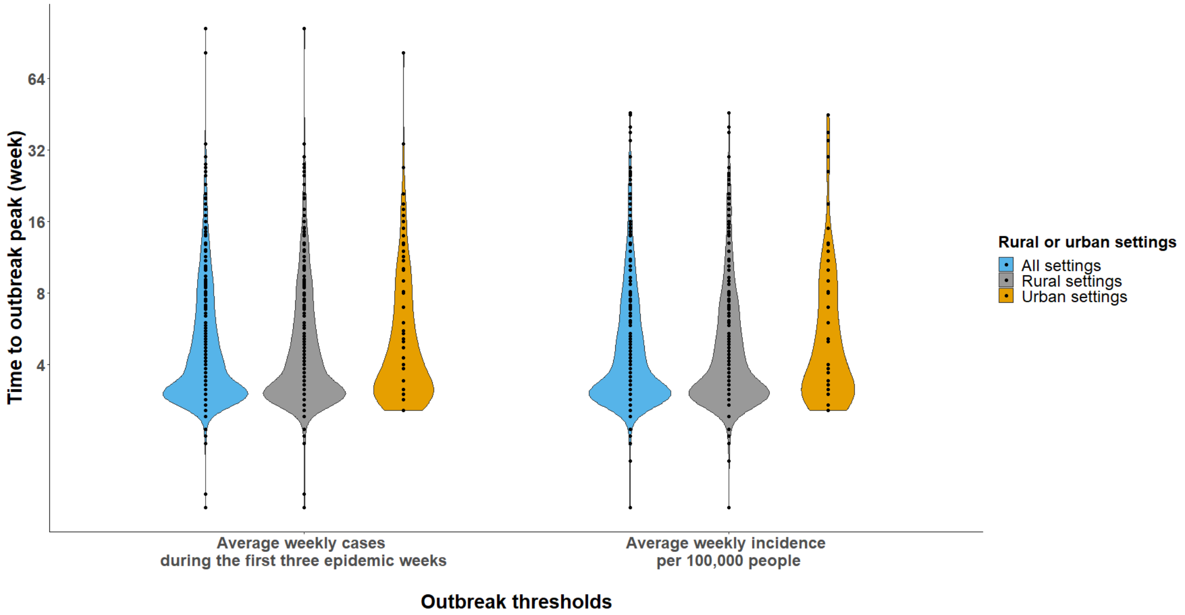


**Figure S55. Distributions of time to outbreak peak (week) stratified by rural and urban settings using different outbreak threshold definitions among outbreaks at the second-level administrative units**

This figure shows the distributions of time to outbreak peak (week) across rural and urban settings using different outbreak threshold definitions. Only outbreaks reported at the second-level administrative units are included.

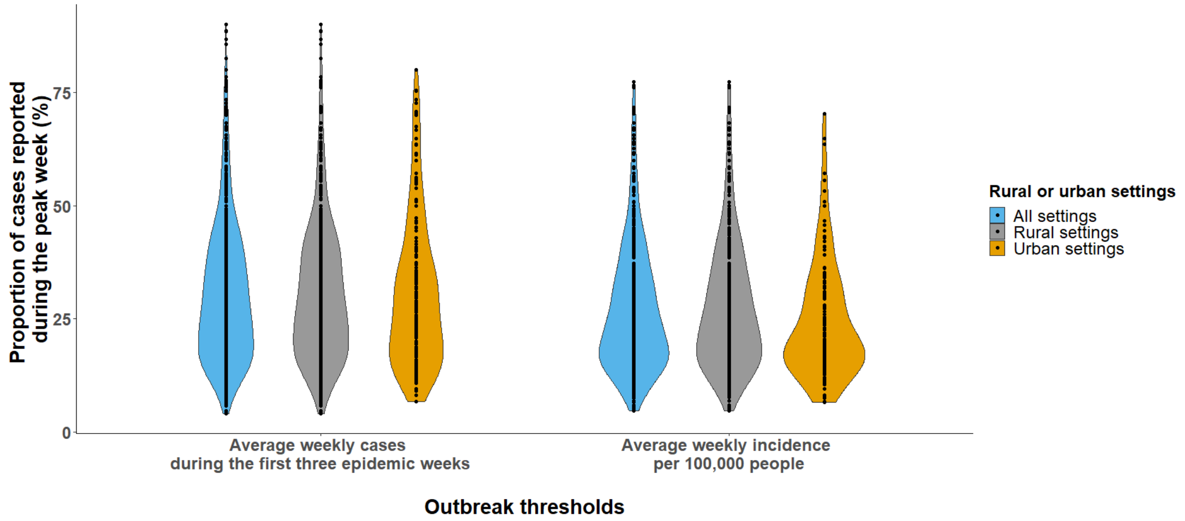


**Figure S56. Distributions of the proportion of cases reported during the peak week (%) stratified by rural and urban settings using different outbreak threshold definitions among outbreaks at the second-level administrative units**

This figure shows the distributions of the proportion of cases reported during the peak week (%) across rural and urban settings using different outbreak threshold definitions. Only outbreaks reported at the first-level administrative units are included.

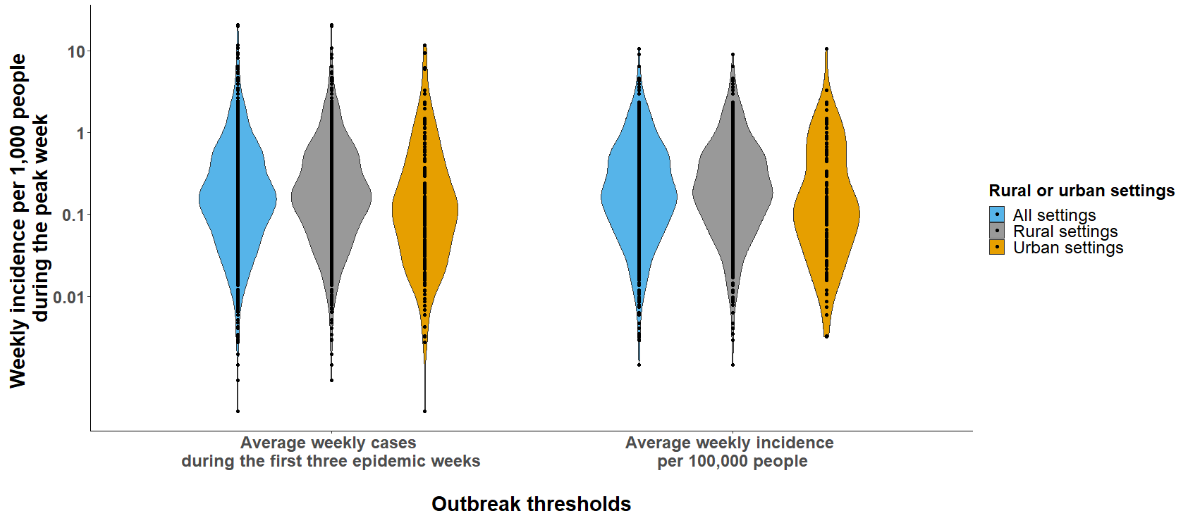


**Figure S57. Distributions of the weekly incidence per 1,000 people during the peak week stratified by rural and urban settings using different outbreak threshold definitions among outbreaks at the second-level administrative units**

This figure shows the distributions of the weekly incidence per 1,000 people during the peak week across rural and urban settings using different outbreak threshold definitions. Only outbreaks reported at the second-level administrative units are included.

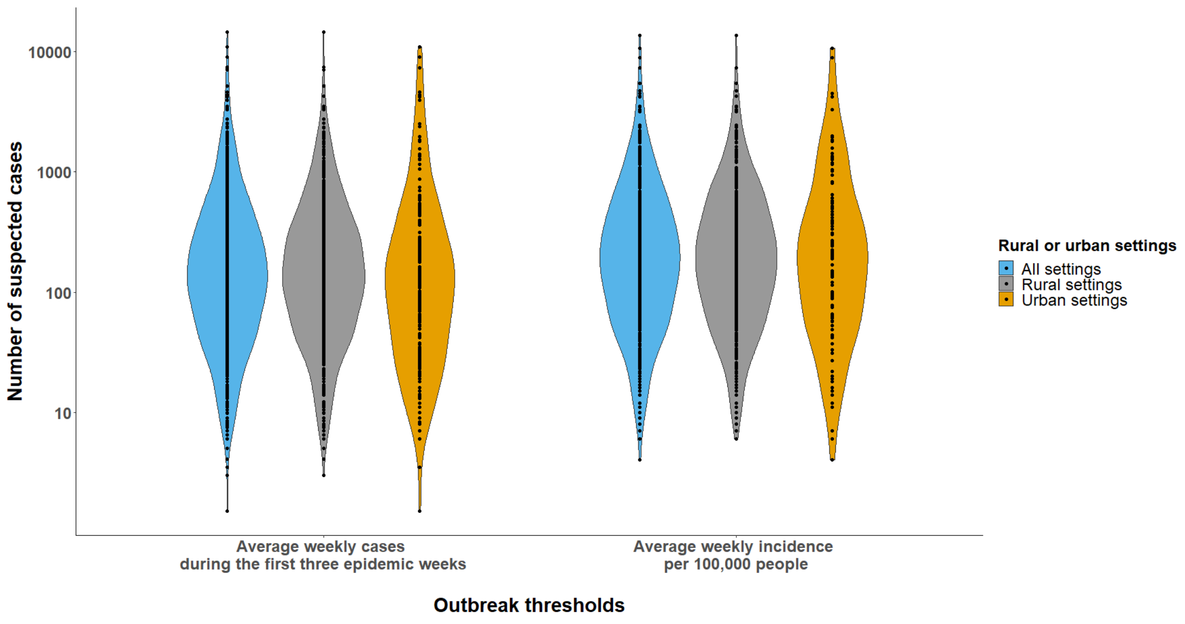

**Figure S58. Distributions of number of suspected cases stratified by rural and urban settings using different outbreak threshold definitions among outbreaks at the second-level administrative units**

This figure shows the distributions of number of suspected cases across rural and urban settings using different outbreak threshold definitions. Only outbreaks reported at the second-level administrative units are included.

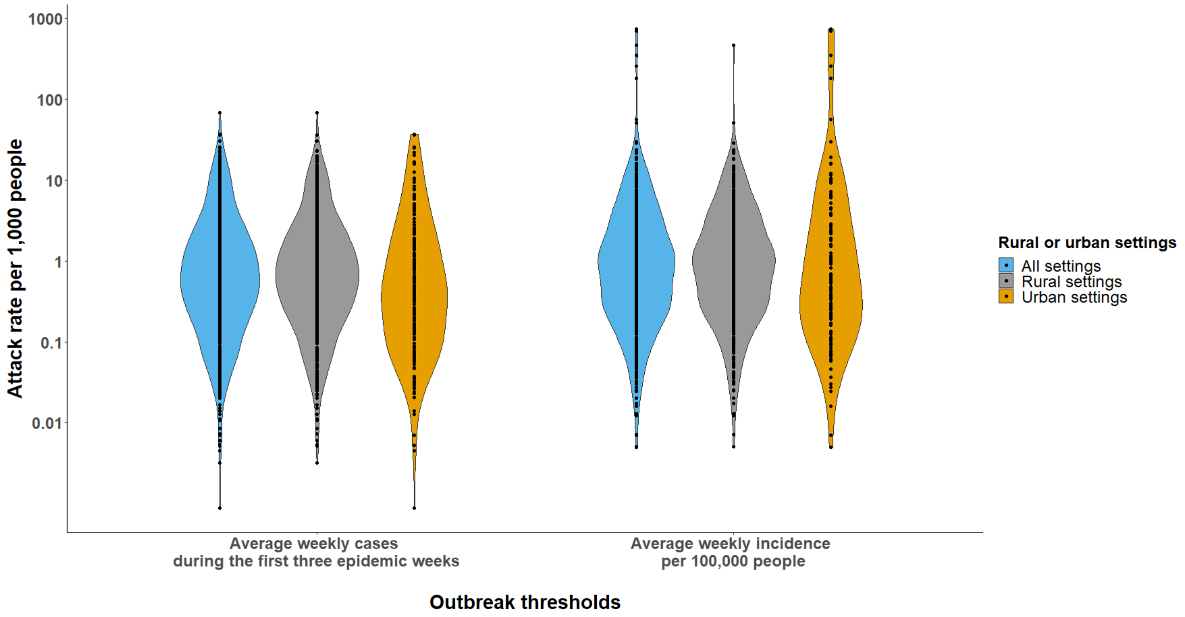


**Figure S59. Distributions of attack rates stratified by rural and urban settings using different outbreak threshold definitions among outbreaks at the second-level administrative units**

This figure shows the distributions of attack rates per 1,000 people across rural and urban settings using different outbreak threshold definitions. Only outbreaks reported at the second-level administrative units are included.

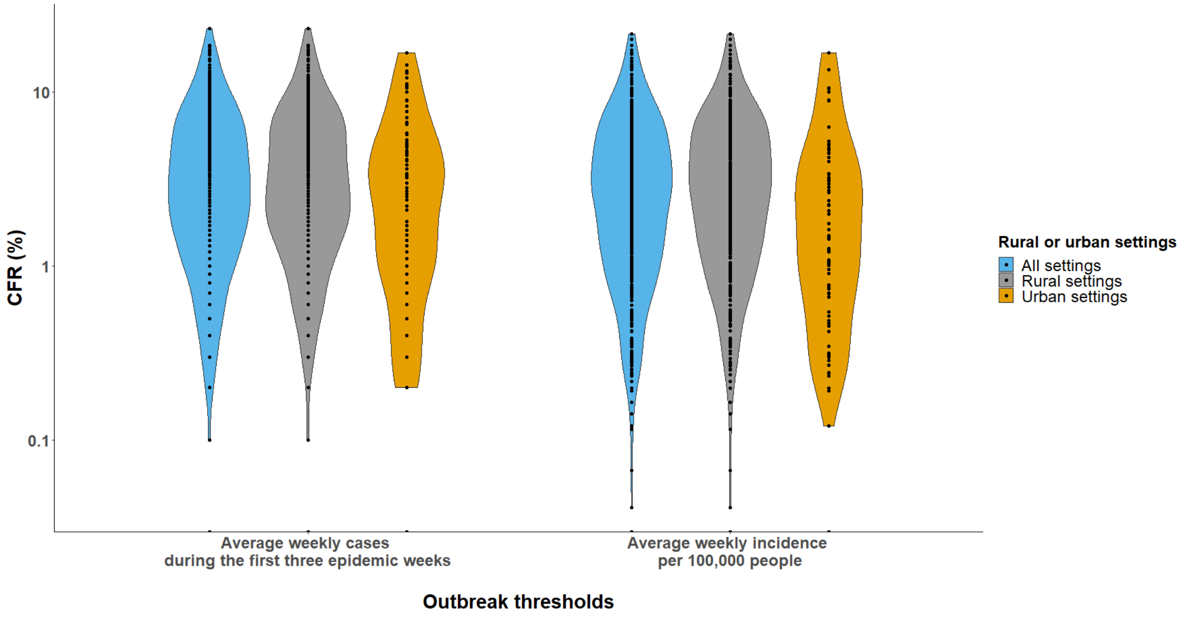


**Figure S60. Distributions of CFRs (%) stratified by rural and urban settings using different outbreak threshold definitions among outbreaks at the second-level administrative units**

This figure shows the distributions of CFR (%) across rural and urban settings using different outbreak threshold definitions. Only outbreaks reported at the second-level administrative units are included.

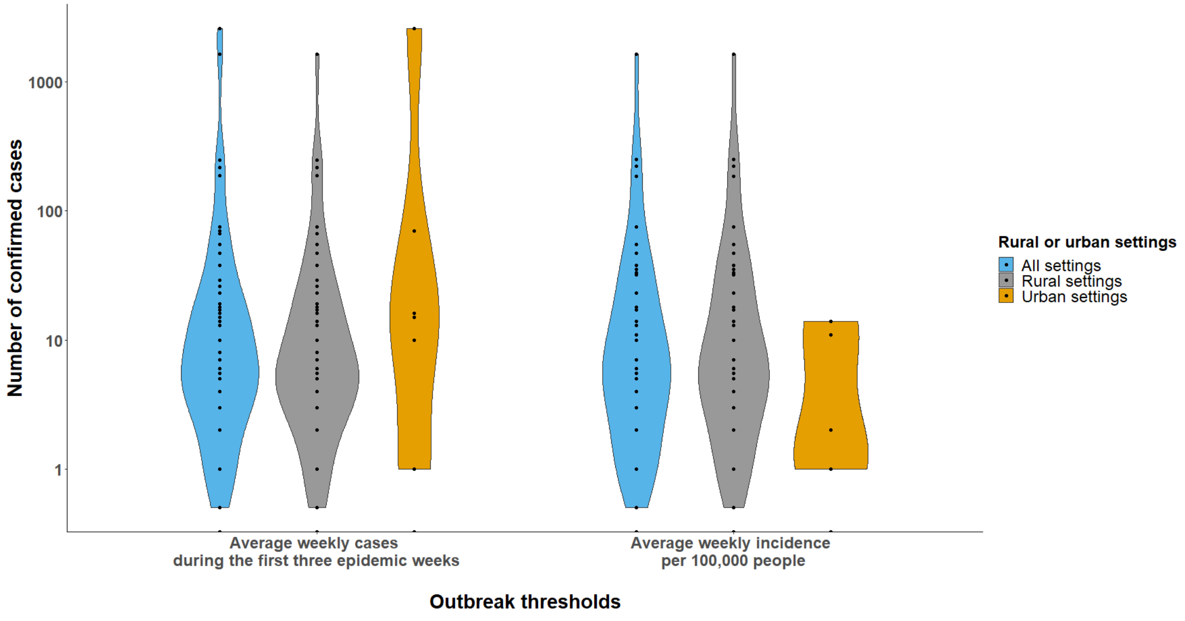

**Figure S61. Distributions of number of confirmed cases stratified by rural and urban settings using different outbreak threshold definitions among outbreaks at the second-level administrative units**

This figure shows the distributions of number of confirmed cases across rural and urban settings using different outbreak threshold definitions. Only outbreaks reported at the second-level administrative units are included.

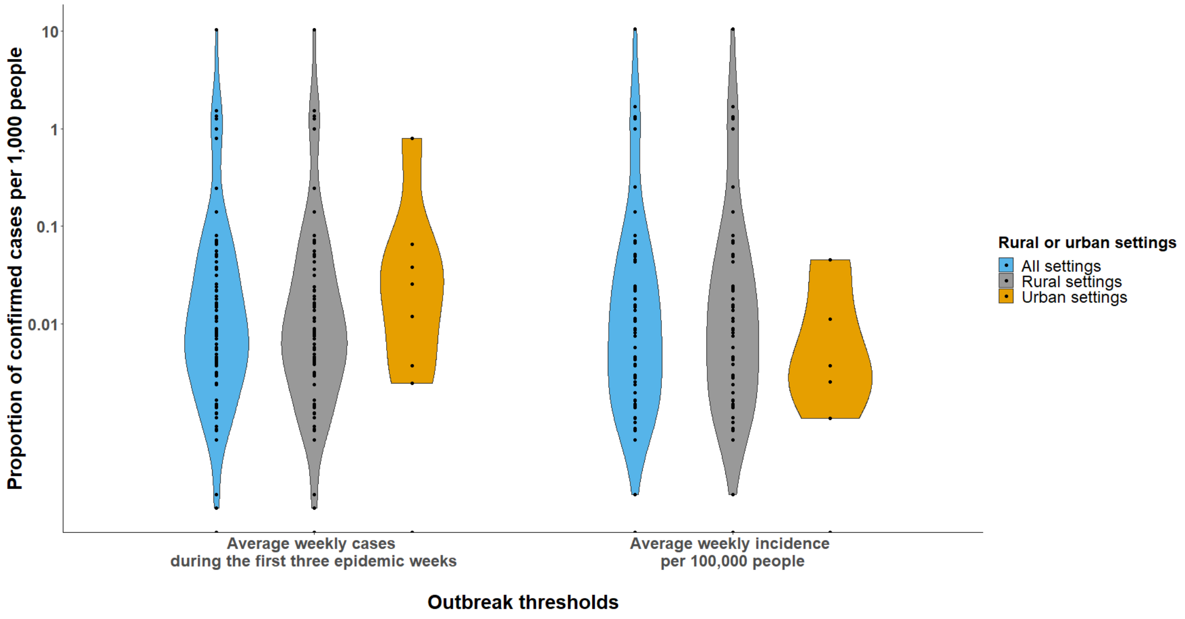


**Figure S62. Distributions of proportion of confirmed cases per 1,000 people stratified by rural and urban settings using different outbreak threshold definitions among outbreaks at the second-level administrative units**

This figure shows the distributions of proportion of confirmed cases per 1,000 people across rural and urban settings using different outbreak threshold definitions. Only outbreaks reported at the second-level administrative units are included.

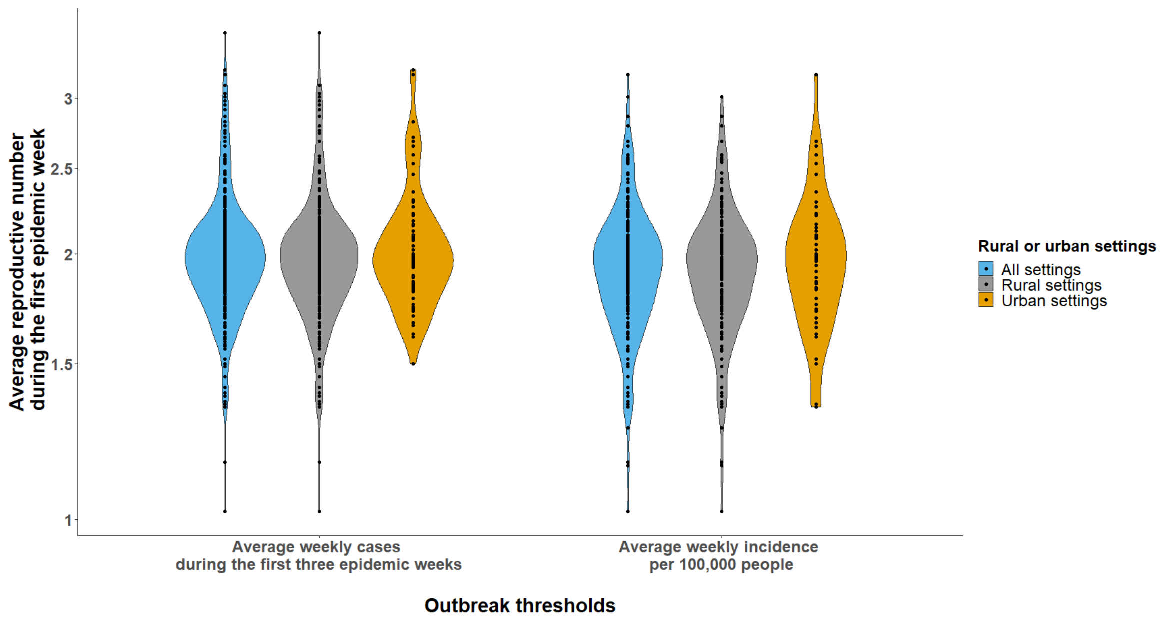


**Figure S63. Distributions of average reproductive numbers during the first epidemic week stratified by rural and urban settings using different outbreak threshold definitions among outbreaks at the third-level administrative units**
This figure shows the distributions of average reproductive numbers during the first epidemic week across rural and urban settings using different outbreak threshold definitions. Only outbreaks reported at the third-level administrative units are included.

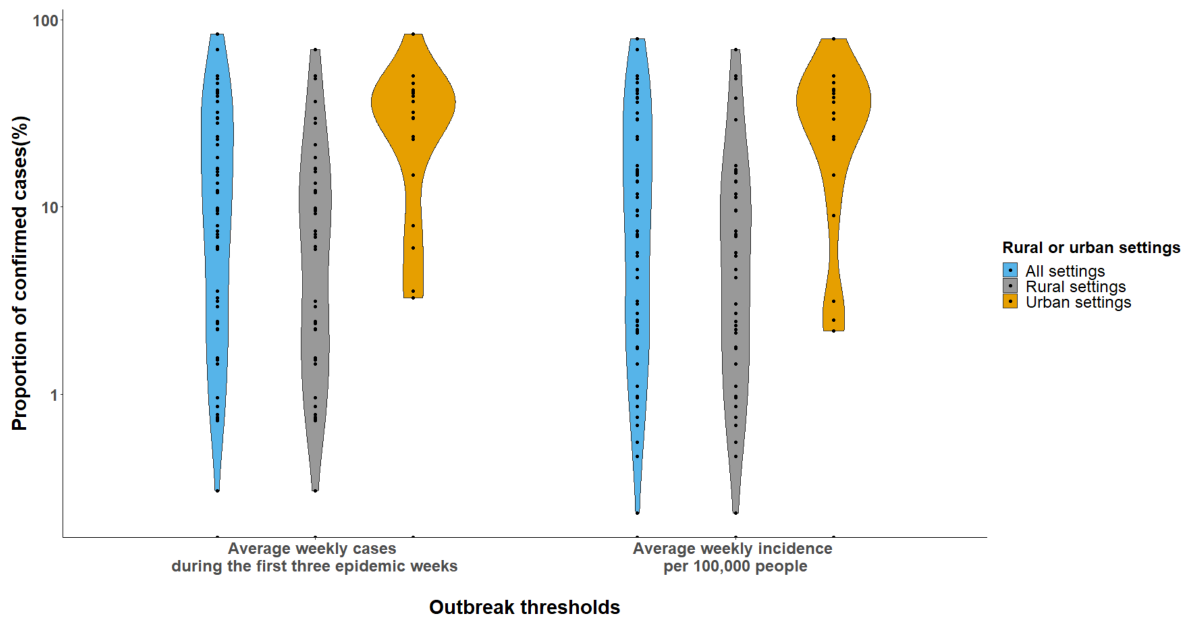

**Figure S64. Distributions of proportion of confirmed cases (%) stratified by rural and urban settings using different outbreak threshold definitions among outbreaks at the third-level administrative units**

This figure shows the distributions of proportion of confirmed cases (%) across rural and urban settings using different outbreak threshold definitions. Only outbreaks reported at the third-level administrative units are included.

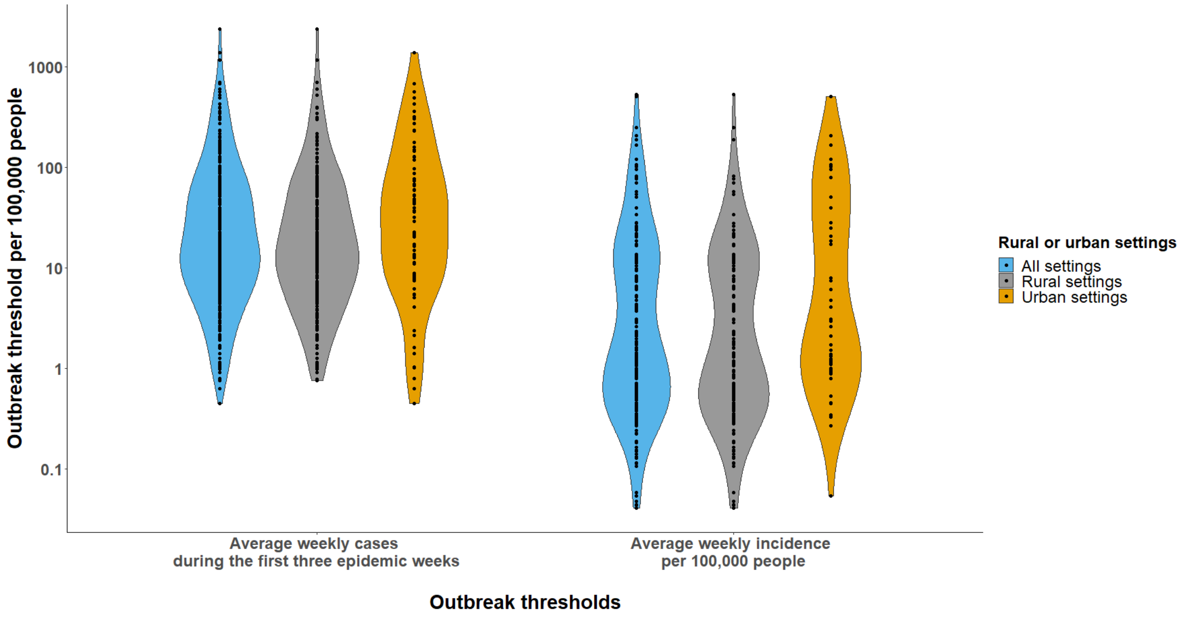


**Figure S65. Distributions of outbreak thresholds per 100,000 people stratified by rural and urban settings using different outbreak threshold definitions among outbreaks at the third-level administrative units**

This figure shows the distributions of outbreak thresholds per 100,000 people across rural and urban settings using different outbreak threshold definitions. Only outbreaks reported at the third-level administrative units are included.

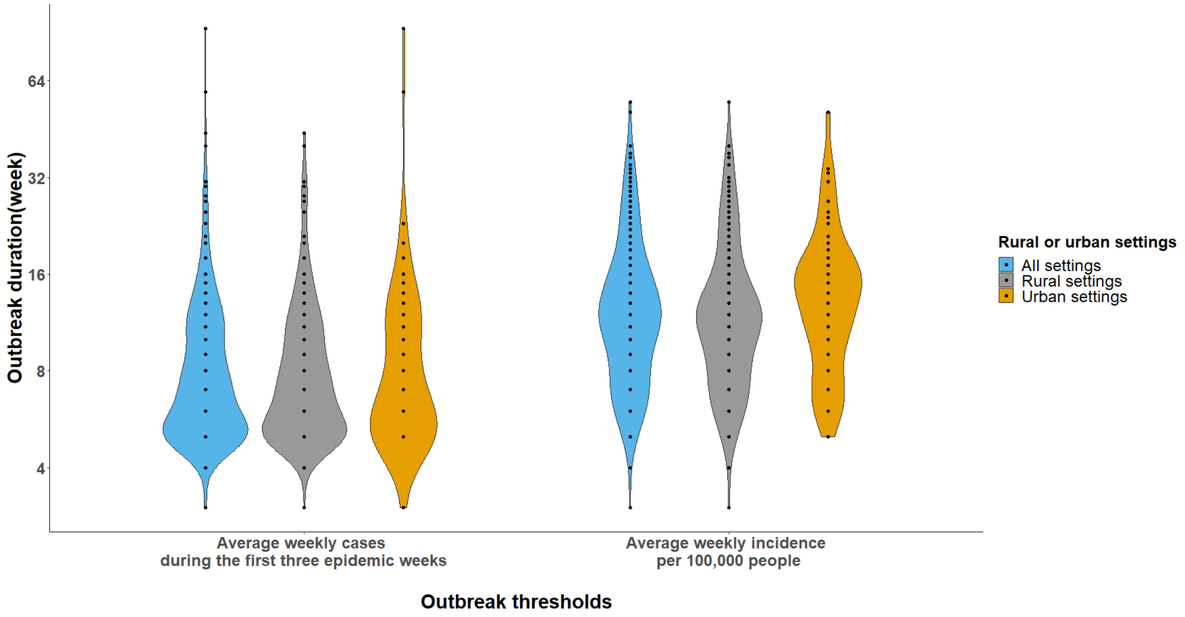


**Figure S66. Distributions of outbreak durations (week) stratified by rural and urban settings using different outbreak threshold definitions among outbreaks at the third-level administrative units**

This figure shows the distributions of outbreak durations (week) across rural and urban settings using different outbreak threshold definitions. Only outbreaks reported at the third-level administrative units are included.

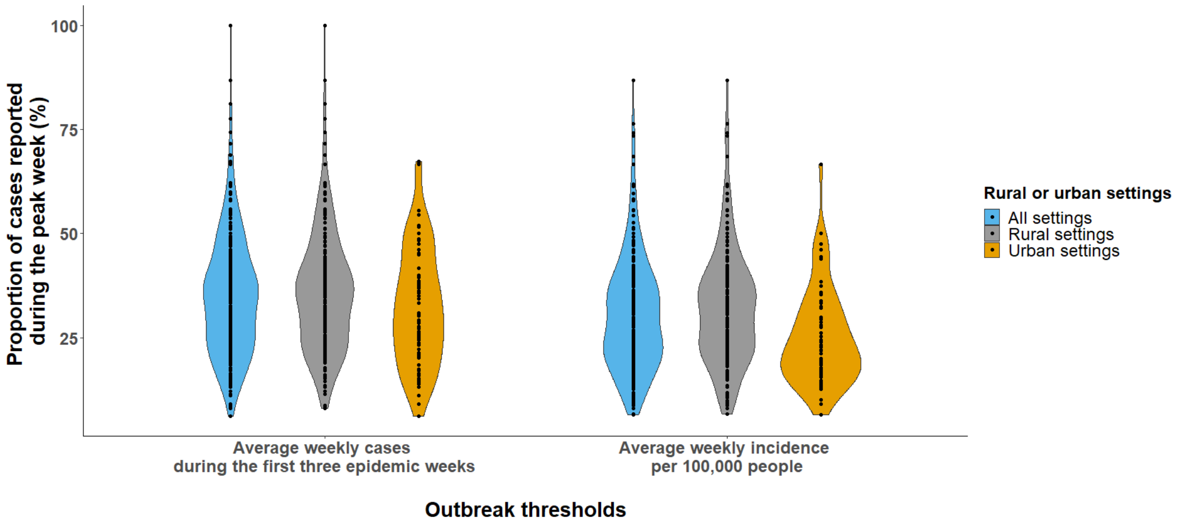


**Figure S67. Distributions of the proportion of cases reported during the peak week (%) stratified by rural and urban settings using different outbreak threshold definitions among outbreaks at the third-level administrative units**

This figure shows the distributions of the proportion of cases reported during the peak week (%) across rural and urban settings using different outbreak threshold definitions. Only outbreaks reported at the third-level administrative units are included.

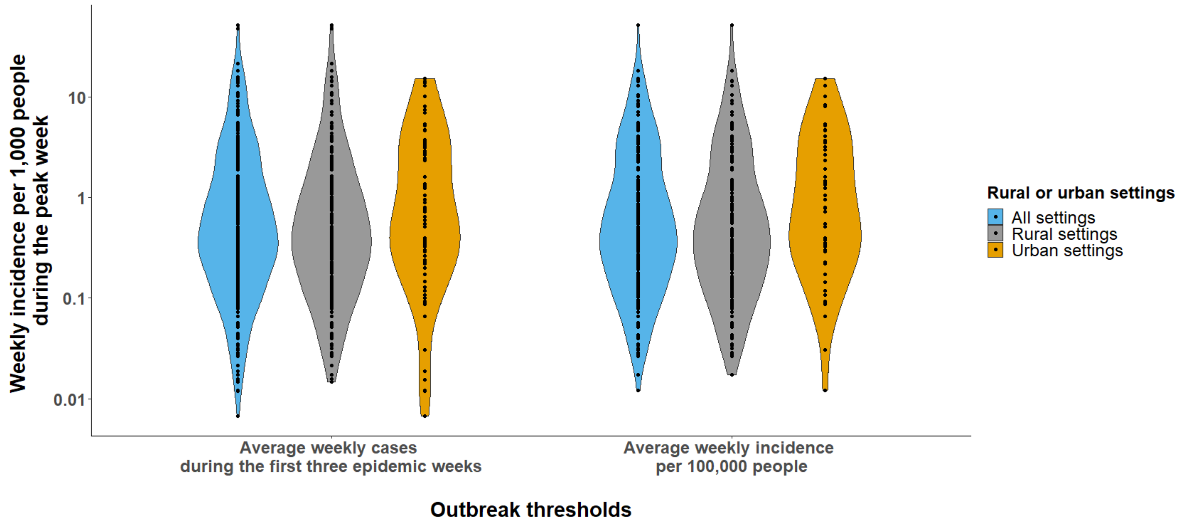


**Figure S68. Distributions of the weekly incidence per 1,000 people during the peak week stratified by rural and urban settings using different outbreak threshold definitions among outbreaks at the third-level administrative units**

This figure shows the distributions of the weekly incidence per 1,000 people during the peak week across rural and urban settings using different outbreak threshold definitions. Only outbreaks reported at the third-level administrative units are included.

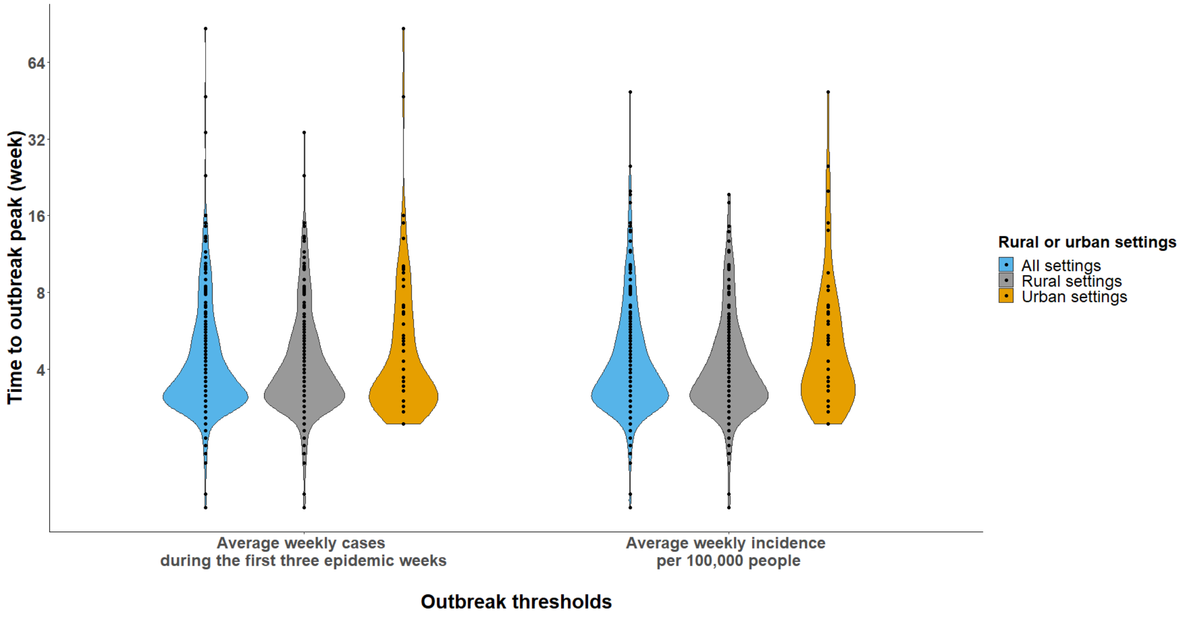

**Figure S69. Distributions of time to outbreak peak (week) stratified by rural and urban settings using different outbreak threshold definitions among outbreaks at the third-level administrative units**

This figure shows the distributions of time to outbreak peak (week) across rural and urban settings using different outbreak threshold definitions. Only outbreaks reported at the third-level administrative units are included.

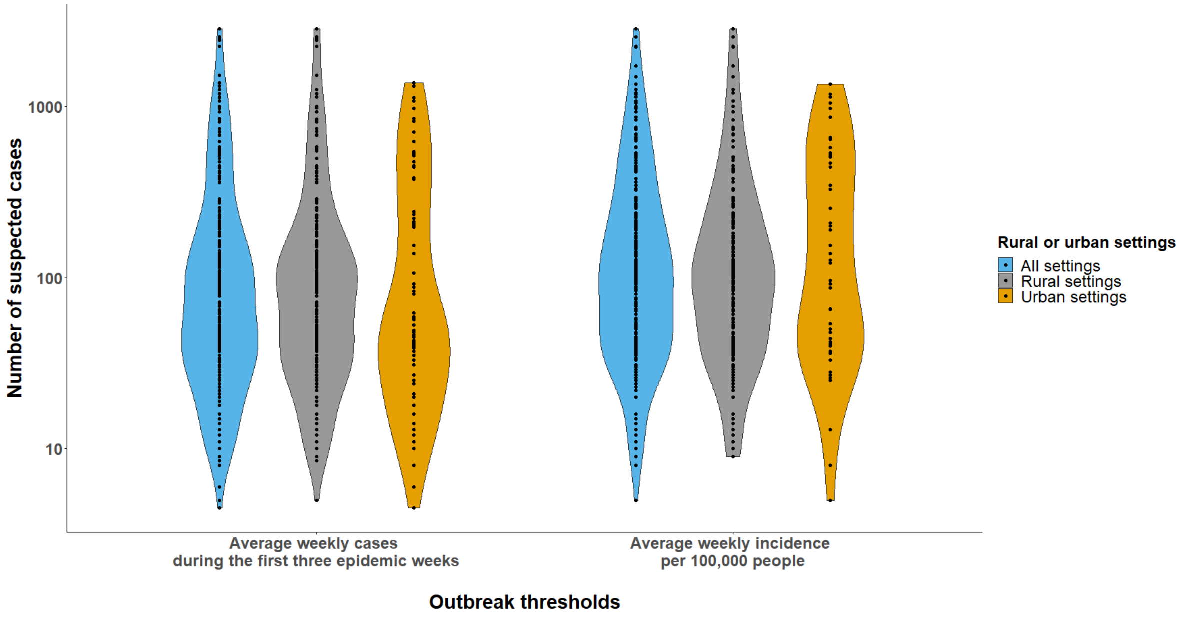


**Figure S70. Distributions of number of suspected cases stratified by rural and urban settings using different outbreak threshold definitions among outbreaks at the third-level administrative units**

This figure shows the distributions of number of suspected cases across rural and urban settings using different outbreak threshold definitions. Only outbreaks reported at the third-level administrative units are included.

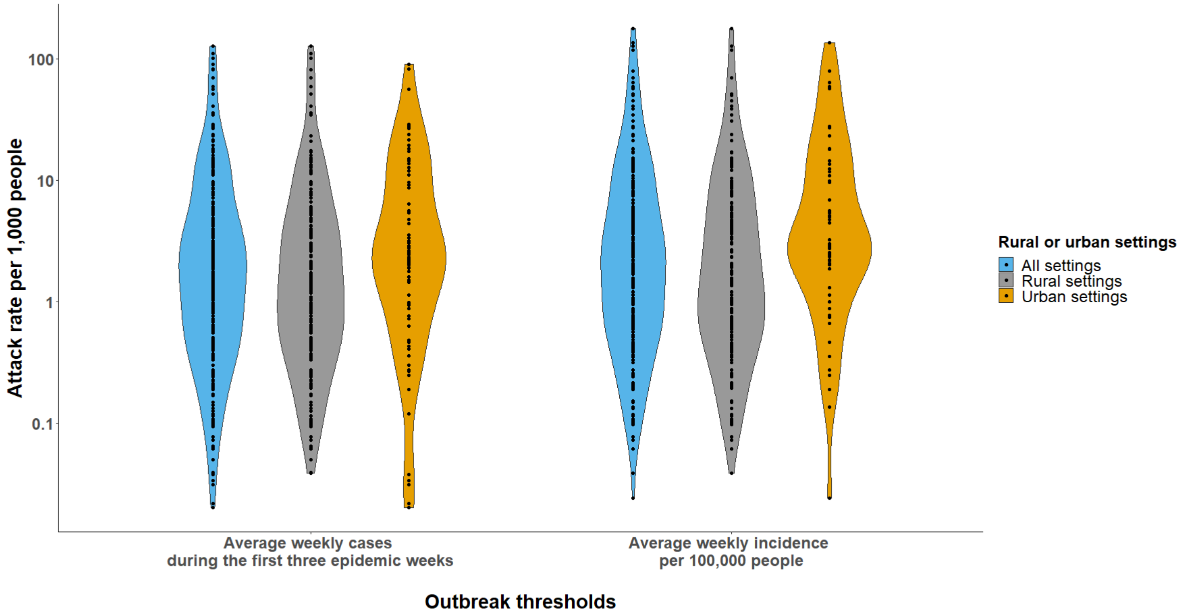


**Figure S71. Distributions of attack rates stratified by rural and urban settings using different outbreak threshold definitions among outbreaks at the third-level administrative units**

This figure shows the distributions of attack rates per 1,000 people across rural and urban settings using different outbreak threshold definitions. Only outbreaks reported at the third-level administrative units are included.

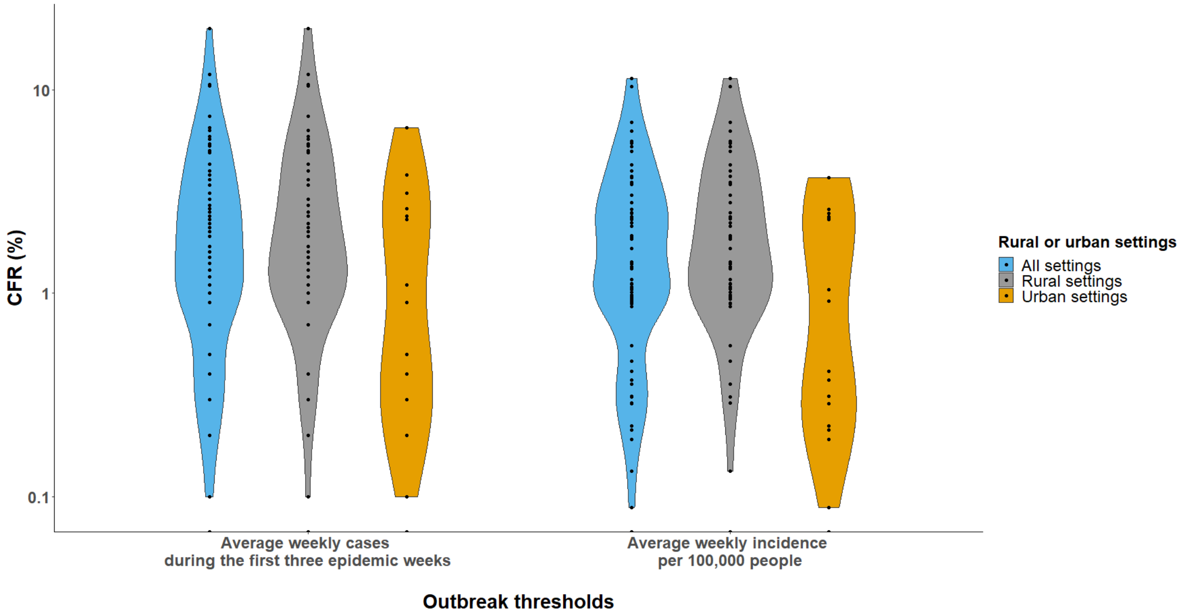

**Figure S72. Distributions of CFRs (%) stratified by rural and urban settings using different outbreak threshold definitions among outbreaks at the third-level administrative units**

This figure shows the distributions of CFR (%) across rural and urban settings using different outbreak threshold definitions. Only outbreaks reported at the third-level administrative units are included.

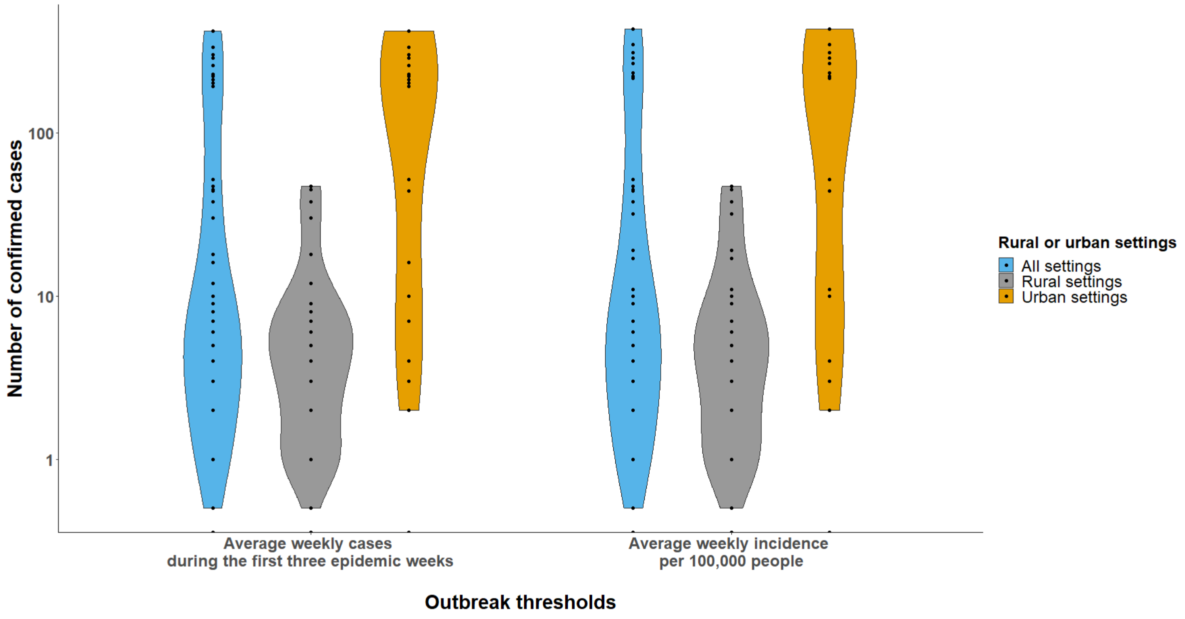


**Figure S73. Distributions of number of confirmed cases stratified by rural and urban settings using different outbreak threshold definitions among outbreaks at the third-level administrative units**

This figure shows the distributions of the number of confirmed cases across rural and urban settings using different outbreak threshold definitions. Only outbreaks reported at the third-level administrative units are included.

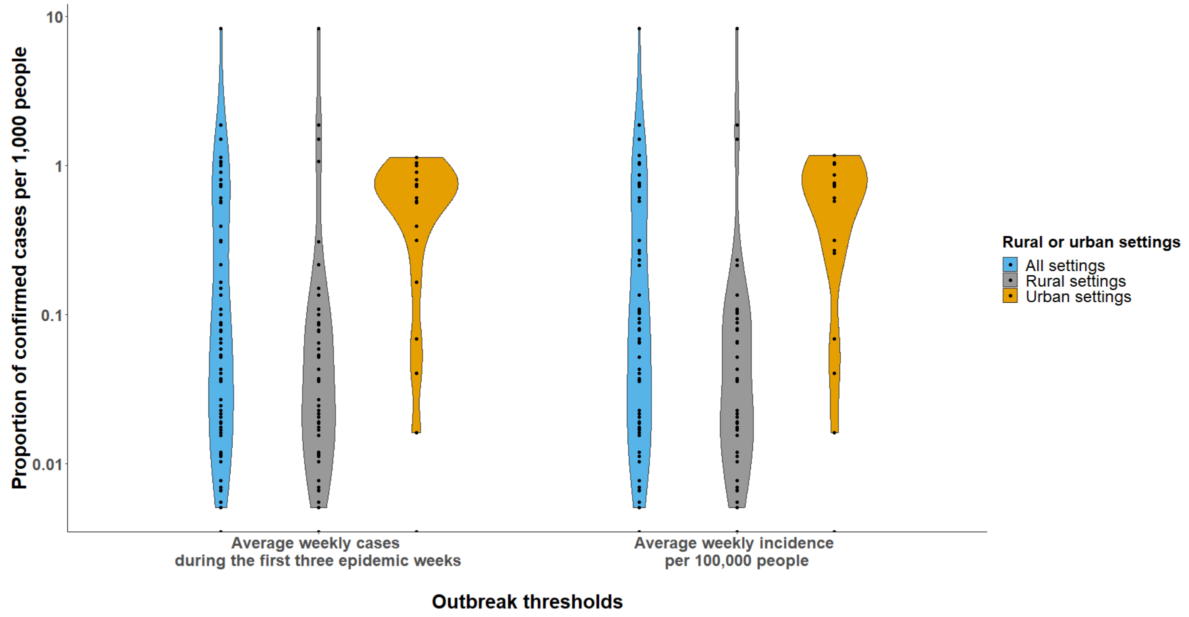


**Figure S74. Distributions of proportion of confirmed cases per 1,000 people stratified by rural and urban settings using different outbreak threshold definitions among outbreaks at the third-level administrative units**

This figure shows the distributions of proportion of confirmed cases per 1,000 people across rural and urban settings using different outbreak threshold definitions. Only outbreaks reported at the third-level administrative units are included.

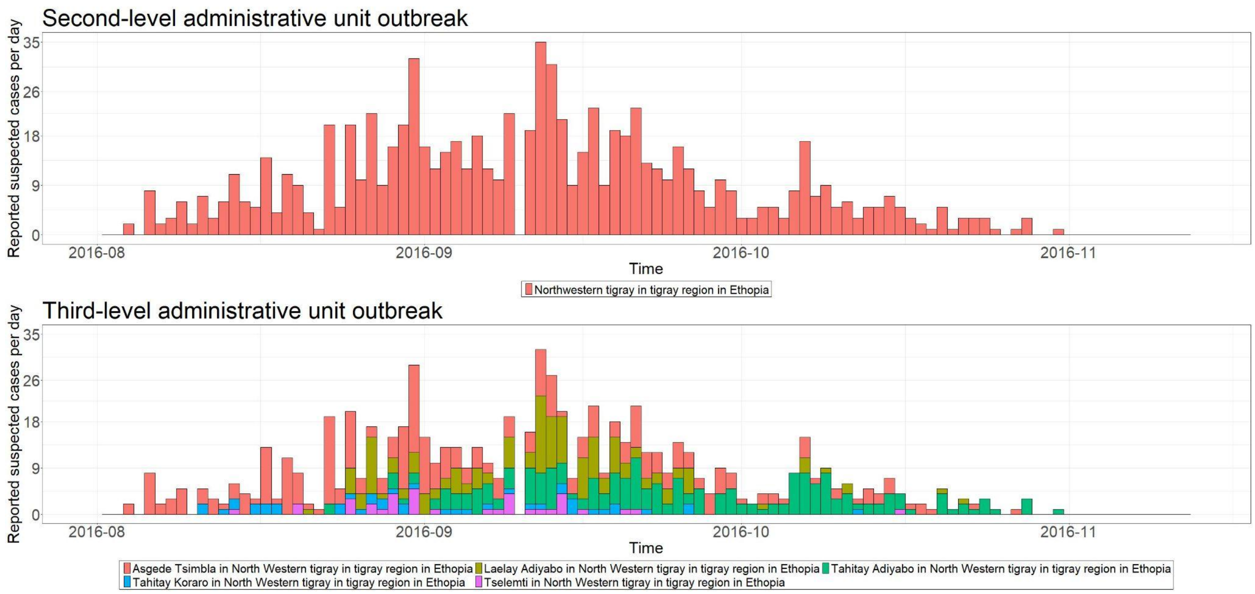


**Figure S75. Epidemic curves for the outbreak reported in North Western Tigray in Tigray region in Ethiopia.**

This figure shows the epidemic curves for the same outbreak reported at both second- and third-level administrative units. The first row represents the epidemic curve for the outbreak reported at the second-level administrative unit. The second row represents the epidemic curves for the outbreaks spanning over multiple third-level administrative units.

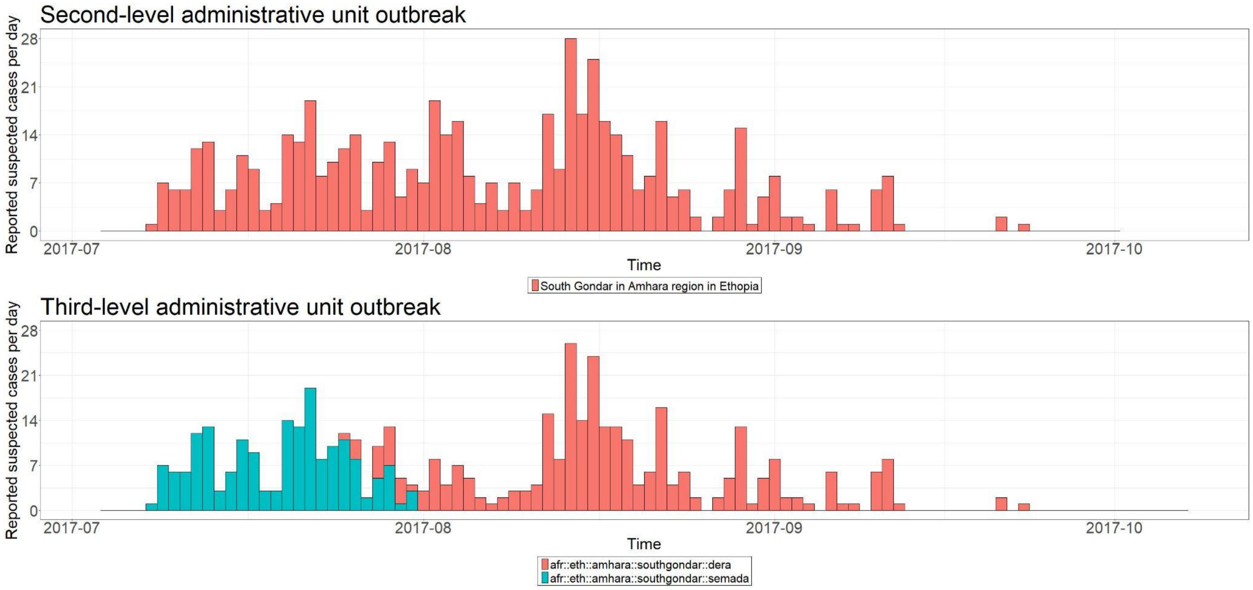


**Figure S76. Epidemic curves for the outbreak reported in South Gondar in Amhara region in Ethiopia.**

This figure shows the epidemic curves for the same outbreak reported at both second- and third-level administrative units. The first row represents the epidemic curve for the outbreak reported at the second-level administrative unit. The second row represents the epidemic curves for the outbreaks spanning over multiple third-level administrative units.

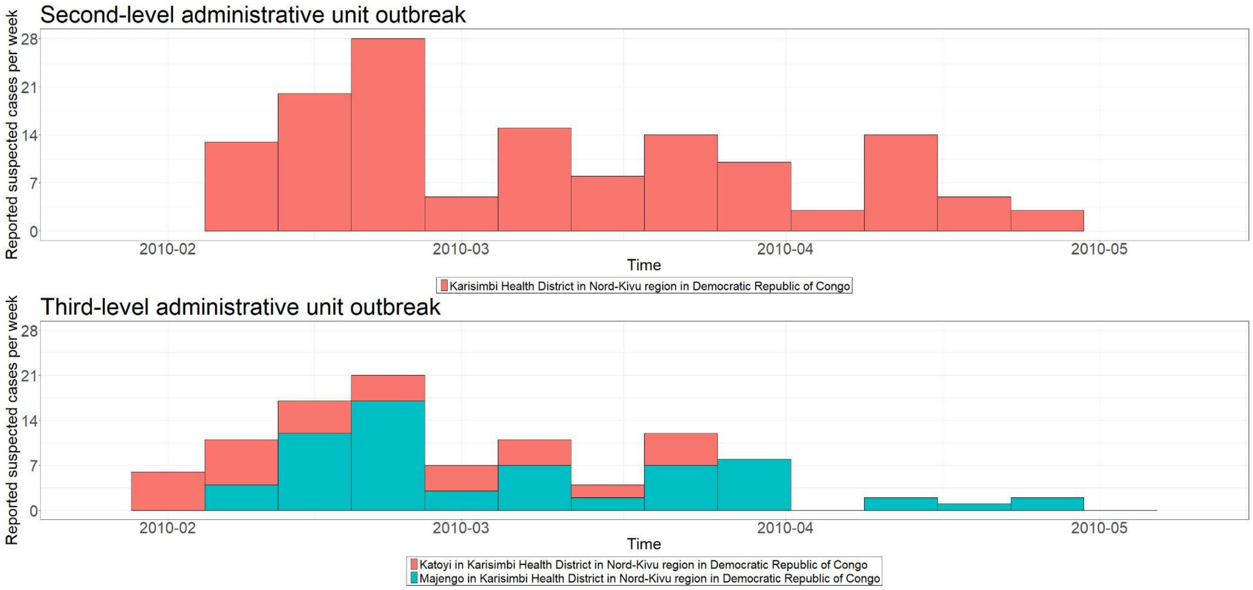


**Figure S77. Epidemic curves for the outbreak reported in Karisimbi health district in Nord-Kivu region in Democratic republic of Congo.**

This figure shows the epidemic curves for the same outbreak reported at both second- and third-level administrative units. The first row represents the epidemic curve for the outbreak reported at the second-level administrative unit. The second row represents the epidemic curves for the outbreaks spanning over multiple third-level administrative units.

### Reference

1. FAO Map Catalog [Internet]. [cited 2021 Jul 8]. Available from: http://www.fao.org/geonetwork

2. GADM [Internet]. [cited 2021 Jul 8]. Available from: https://gadm.org/

3. Welcome - humanitarian data exchange [Internet]. [cited 2021 Jul 8]. Available from: https://data.humdata.org/

4. Cori A, Ferguson NM, Fraser C, Cauchemez S. A new framework and software to estimate time-varying reproduction numbers during epidemics. Am J Epidemiol. 2013 Nov 1;178(9):1505–12.

5. Bi Q, Abdalla FM, Masauni S, Reyburn R, Msambazi M, Deglise C, et al. The Epidemiology of Cholera in Zanzibar: Implications for the Zanzibar Comprehensive Cholera Elimination Plan. J Infect Dis. 2018 Oct 15;218(suppl_3):S173–80.

6. Jones FK, Wamala JF, Rumunu J, Mawien PN, Kol MT, Wohl S, et al. Successive epidemic waves of cholera in South Sudan between 2014 and 2017: a descriptive epidemiological study. Lancet Planet Health. 2020 Dec;4(12):e577–87.

7. Becker RA, Chambers JM, Wilks AR. The New S Language: A Programming Environment for Data Analysis and Graphics. Thomson Brooks/Cole; 1988. 702 p.

8. Forsythe GE, Malcolm MA, Moler CB, Moler C. Computer Methods for Mathematical Computations. Prentice Hall; 1977. 259 p.
